# Population Genetics Analysis of *SLC3A1* and *SLC7A9* Revealed the Etiology of Cystine Stone May Be More Than What Our Current Genetic Knowledge Can Explain

**DOI:** 10.1101/2023.05.09.23289733

**Authors:** Chen-Han Wilfred Wu, Jad Badreddine, Joshua Chang, Yu-Ren Mike Huang, Fernando J. Kim, Trevor Wild, Anne Tsai, Naomi Meeks, Rodrigo Donalisio Da Silva, Wilson R. Molina, Fredrick R. Schumacher

## Abstract

**Background:** Cystine stone is a Mendelian genetic disease caused by *SLC3A1* or *SLC7A9*. In this study, we aimed to estimate the genetic prevalence of cystine stones and compare it with the clinical prevalence to better understand the disease etiology.

**Methods:** We analyzed genetic variants in the general population using the 1000 Genomes project and the Human Gene Mutation Database to extract all *SLC3A1* and *SLC7A9* pathogenic variants. All variants procured from both databases were intersected. Pathogenic allele frequency, carrier rate, and affected rate were calculated and estimated based on Hardy-Weinberg equilibrium.

**Results:** We found that 9 unique *SLC3A1* pathogenic variants were carried by 26 people and 5 unique *SLC7A9* pathogenic variants were carried by 12 people, all of whom were heterozygote carriers. No homozygote, compound heterozygote, or double heterozygote was identified in the 1000 Genome database. Based on the Hardy-Weinberg equilibrium, the calculated genetic prevalence of cystine stone disease is 1 in 30,585.

**Conclusion:** The clinical prevalence of cystine stone has been previously reported as 1 in 7,000, a notably higher figure than the genetic prevalence of 1 in 30,585 calculated in this study. This suggests that the etiology of cystine stone is more complex than what our current genetic knowledge can explain. Possible factors that may contribute to this difference include novel causal genes, undiscovered pathogenic variants, alternative inheritance models, founder effects, epigenetic modifications, environmental factors, or other modifying factors. Further investigation is needed to fully understand the etiology of cystine stone.

## Introduction

Cystine stones are the product of a metabolic defect, cystinuria, characterized by the excessive excretion of cystine in the urine [1]. This metabolic disorder is a rare heritable condition caused by a defective amino acid transport system that prevents the reabsorption of cystine and other dibasic amino acids in the kidneys and intestinal epithelium [2]. Given their low solubility at physiologic pH, continuous accumulation of cystine in the urine promotes the formation of kidney stones and increases the risk of renal damage [3]. Cystinuria is responsible for approximately 6-8% of renal calculi in pediatrics and accounts for 1-2% of all kidney stone cases. Patients with cystine stones are usually identified at a young age as the vast majority present with their first stone episode during adolescence or early adulthood [4, 5].

The prevalence of cystinuria is shown to be highly variable among different countries ranging from 1 in 2,000 in Great Britain to 1 in 100,000 in Sweden [1, 6]. Efforts to uncover the genetic etiologies of cystine stones have identified two causal genes that are involved in the pathophysiology of the disease: *SLC3A1*, which is located on chromosome 2p16, and *SLC7A9*, which is located on chromosome 19q13 [7, 8]. These genes were found to encode for the two subunit proteins that make up the amino acid transporter [9].

Cystinuria was traditionally classified into three types based on the urinary excretion patterns of obligate heterozygotes. However, this phenotypic classification was shown to be inaccurate as it poorly correlated with the levels of aminoaciduria seen in heterozygote individuals carrying the same pathogenic variant.[8, 10]. Following the identification of *SLC3A1* and *SLC7A9* genes, a new genotypic classification system was introduced dividing cystinuria into several subtypes: type A if pathogenic variants are found in both *SLC3A1* alleles, type B if pathogenic variants are found in both *SLC7A9* alleles, and putative type AB if mutations exist in both genes [11].

Genotyping patients with cystine stones is not routinely performed as it currently has no implications for their treatment and clinical course. Reports investigating the genetic epidemiology of cystine stones are relatively scarce in the literature [12]. Our study aims to determine the genetic prevalence of cystine stones caused by type A and type B cystinuria through analyzing genetic information in population databases. We believe that understanding the frequency and distribution of these genetic variants will enable future efforts to develop a more personalized approach to diagnose and manage cystine stones.

## Materials and Methods

*SLC3A1* and *SLC7A9* variants in the 1000 Genomes (1KG) database were identified (Table 1 and Supplementary Tables 1 & 2). Related individuals were excluded. The 1000 Genomes database is a comprehensive database of genomes obtained from healthy individuals in multiple populations around the world, and it identifies 98% of alleles whose frequency is more than 1% in these populations [13]. Lower frequency alleles are also identified in these populations. Known *SLC3A1* and *SLC7A9* pathogenic variants associated with cystine stones were procured from the Human Gene Mutation Database (HGMD) (HGMD® Professional, http://www.hgmd.cf.ac.uk/ac/index.php, accessed in 2016). Variants included missense variants, nonsense variants, insertions, and deletions (Table 2). Variants present in both databases were compared and intersected, resulting in 14 disease-causing variants present in both the HGMD and the 1KG database (Table 3). The references associated with these variants were manually reviewed to ensure that they were correctly associated with cystine stone disease. The presence of homozygotes, compound heterozygotes, multiple variants in cis or trans, double homozygotes, and double heterozygotes in the dataset was examined (Figure 1).

**Table 1.** Classification of SLC3A1 and SLC7A9 unique variants in the 1KG database.

| Unique Variants in 1KG |  |  |  | SLC3A1 (type A) | SLC7A9 (type B) |
| --- | --- | --- | --- | --- | --- |
| SNV | Exon | Coding | Missense | 102 (6%) | 69 (5.4%) |
|  |  |  | Nonsense | 0 | 0 |
|  |  |  | Synonymous | 7 (0.4%) | 7 (0.5%) |
|  |  | UTR |  | 47 (2.7%) | 11 (0.8%) |
|  | Intron |  |  | 1429 (84.7%) | 1124 (88.2%) |
| Deletion | Exon | Coding | Inframe | 0 | 0 |
|  |  |  | Frameshift | 0 | 0 |
|  |  | UTR |  | 0 | 0 |
|  | Intron |  |  | 63 (3.7%) | 40 (3.1%) |
| Insertion | Exon | Coding | Inframe | 0 | 0 |
|  |  |  | Frameshift | 0 | 0 |
|  |  | UTR |  | 0 | 0 |
|  | Intron |  |  | 38 (2.2%) | 23 (1.8%) |
| Complex Substitutions |  |  |  | 0 | 0 |
| Total number of Variants |  |  |  | 1686 (100%) | 1274 (100%) |

1KG: 1000 Genomes Project Phase 3; SNV: Single Nucleotide Variant; UTR: Untranslated Region

**Table 2.** Classification of SLC3A1 and SLC7A9 unique variants in the HGMD database.

| Unique Pathogenic Variants in HGMD |  |  |  | SLC3A1 (type A) | SLC7A9 (type B) |
| --- | --- | --- | --- | --- | --- |
| SNV | Exon | Coding | Missense | 81 (73.6%) | 44 (51.7%) |
|  |  |  | Nonsense | 10 (9%) | 2 (2.3%) |
|  |  |  | Synonymous | 0 | 7 (8.2%) |
|  |  | UTR |  | 0 | 0 |
|  | Intron |  |  | 0 | 0 |
| Deletion | Exon | Coding | Inframe | 1 (0.9%) | 3 (3.5%) |
|  |  |  | Frameshift | 11 (10%) | 20 (23.5%) |
|  |  | UTR |  | 0 | 0 |
|  | Intron |  |  | 0 | 2 (2.3%) |
| Insertion | Exon | Coding | Inframe | 0 | 2 (2.3%) |
|  |  |  | Frameshift | 5 (4.5%) | 4 (4.7%) |
|  |  | UTR |  | 0 | 0 |
|  | Intron |  |  | 0 | 1 (1.1%) |
| Complex Substitutions |  |  |  | 2 (1.8%) | 0 |
| Total number of Pathogenic Variants |  |  |  | 110 (100%) | 85 (100%) |

HGMD: Human Gene Mutation Database; SNV: Single Nucleotide Variant; UTR: Untranslated Region

**Table 3.** Characteristics of intersected genetic variants (pathogenic variants) of SLC3A1 and SLC7A9 in 1KG and HGMD.

|  | Chromosome | Position | Variant Identifier | Reference Allele | Variant Allele | C. nomenclature | P. nomenclature | Exon number | Type | Number of people carrying this variant (all heterogeneous) | Observed Allele Frequency (%) |
| --- | --- | --- | --- | --- | --- | --- | --- | --- | --- | --- | --- |
| SLC3A1 | chr2 | 44507990 | rs140317484 | C | T | c.566C>T | p.Thr189Met | 2/10 | missense | 6 | 6/5008 (0.12%) |
|  | chr2 | 44513202 | rs141587158 | T | C | c.797T>C | p.Phe266Ser | 4/10 | missense | 7 | 7/5008 (0.14%) |
|  | chr2 | 44528215 | rs121912697 | G | A | c.1085G>A | p.Arg362His | 6/10 | missense | 2 | 2/5008 (0.04%) |
|  | chr2 | 44528224 | rs567478582 | G | A | c.1094G>A | p.Arg365Gln | 6/10 | missense | 1 | 1/5008 (0.02%) |
|  | chr2 | 44539726 | rs187962930 | T | C | c.1334T>C | p.Ile445Thr | 8/10 | missense | 2 | 2/5008 (0.04%) |
|  | chr2 | 44539746 | rs201502095 | C | T | c.1354C>T | p.Arg452Trp | 8/10 | missense | 1 | 1/5008 (0.02%) |
|  | chr2 | 44539758 | rs139251285 | C | T | c.1366C>T | p.Arg456Cys | 8/10 | missense | 1 | 1/5008 (0.02%) |
|  | chr2 | 44539773 | rs144162964 | T | C | c.1381T>C | p.Tyr461His | 8/10 | missense | 4 | 4/5008 (0.08%) |
|  | chr2 | 44539792 | rs121912691 | T | C | c.1400T>C | p.Met467Thr | 8/10 | missense | 2 | 2/5008 (0.04%) |
|  | SLC3A1 accumulation |  |  |  |  |  |  |  |  | 26 | 26/5008 (0.52%) |
| SLC7A9 | chr19 | 33321545 | rs146815072 | G | A | c.1445C>T | p.Pro482Leu | 13/13 | missense | 1 | 1/5008 (0.02%) |
|  | chr19 | 33353409 | rs531029519 | C | T | c.562G>A | p.Val188Met | 5/13 | missense | 1 | 1/5008 (0.02%) |
|  | chr19 | 33353427 | rs79389353 | C | T | c.544G>A | p.Ala182Thr | 5/13 | missense | 5 | 5/5008 (0.1%) |
|  | chr19 | 33355112 | rs79987078 | G | A | c.368C>T | p.Thr123Met | 4/13 | missense | 2 | 2/5008 (0.04%) |
|  | chr19 | 33355167 | rs121908480 | C | T | c.313G>A | p.Gly105Arg | 4/13 | missense | 3 | 3/5008 (0.06%) |
|  | SLC7A9 accumulation |  |  |  |  |  |  |  |  | 12 | 12/5008 (0.24%) |

Chromosome position based on Genome Reference Consortium Human Build 37 (GRCh37, hg19)

**Figure 1.**
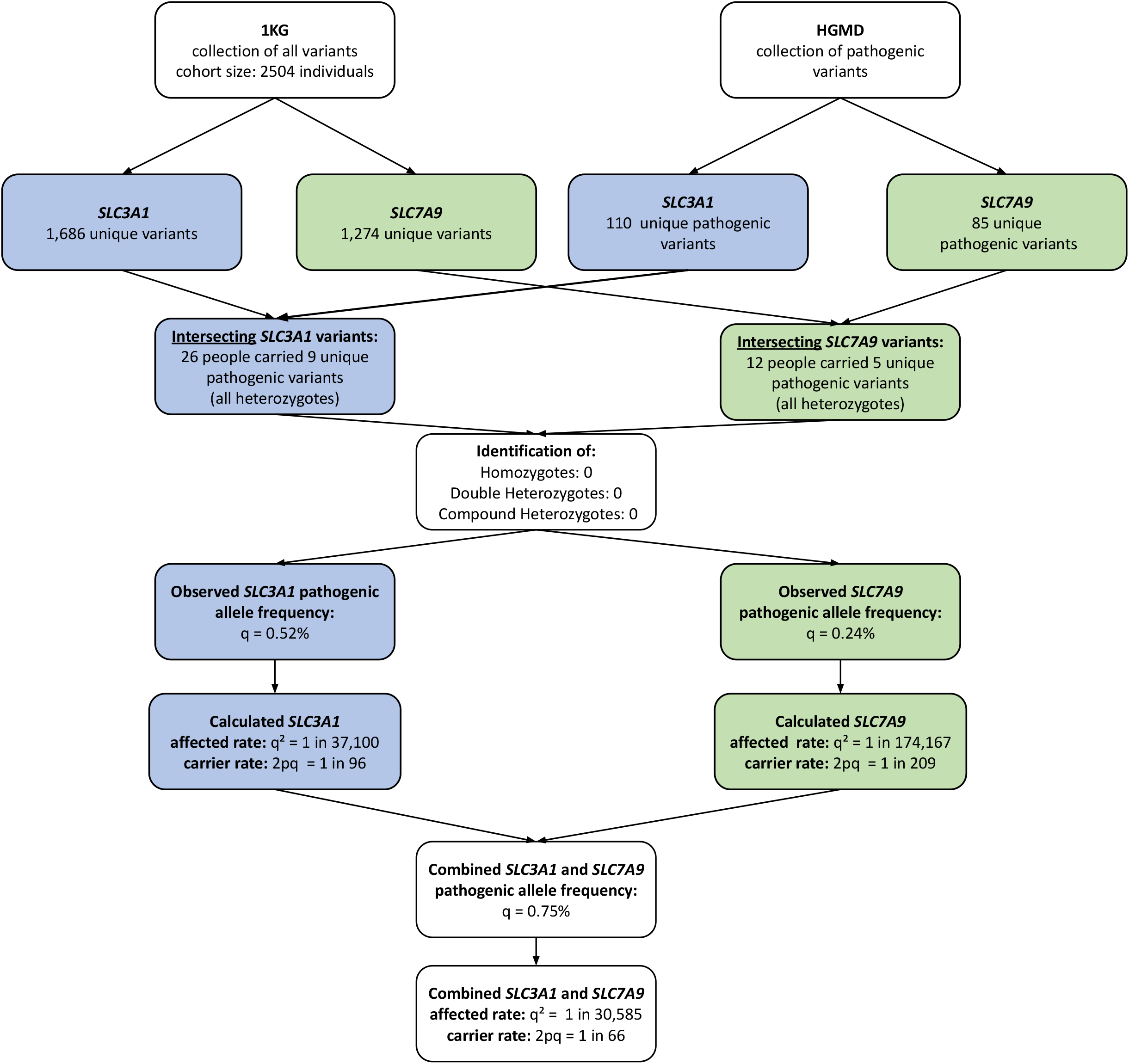
Flowchart to calculate pathogenic allele frequencies, carrier rates, and affected rates of *SLC3A1* and *SLC7A9* for cystine stone. 1KG: 1000 Genomes Project Phase 3 HGMD: Human Gene Mutation Database light blue background: specific to *SLC3A1* light green background: specific to *SLC7A9*

Allele frequencies were then calculated from the 1KG database. By summing allele frequencies over all pathogenic variants, we obtained the cumulative frequency of pathogenic variants in *SLC3A1* and *SLC7A9*. The expected carrier and affected rates were estimated using Hardy–Weinberg equilibrium p^2^ + 2pq + q^2^ = 1, where q is the frequency of pathogenic variants, Table 4 [14]. The calculated prevalence of cystine stones caused by *SLC3A1* and *SLC7A9* pathogenic variants was compared with the observed prevalence of phenotypes in several populations.

**Table 4.**
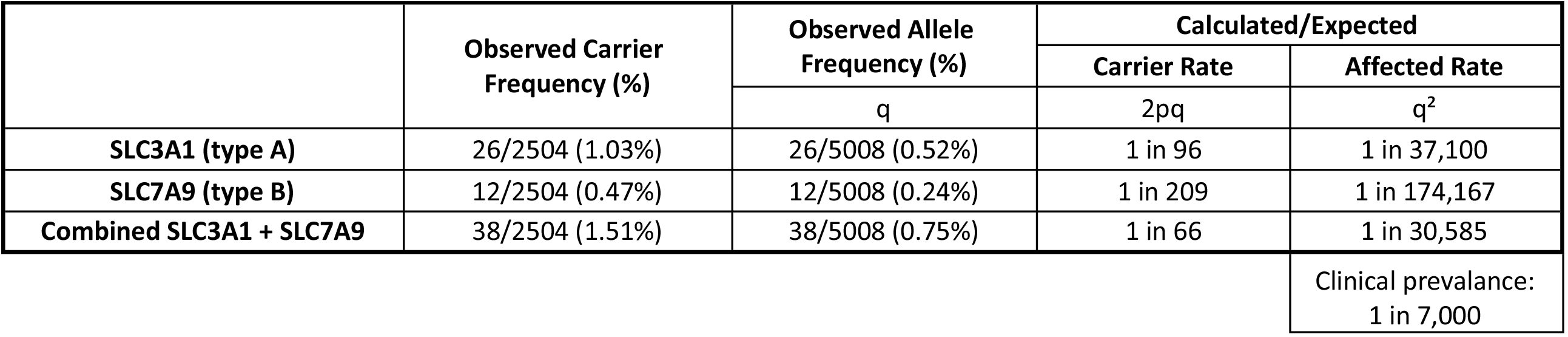
Observed and Calculated allele frequencies using Hardy-Weinberg equilibrium.

## Results

*SLC3A1* and *SLC7A9* variants in the HGMD and 1KG database were identified. The 1KG project comprehensively documents common human genetic variants by sequencing the genomes of 2,504 non-related healthy individuals from 26 populations worldwide [15]. Our results are demonstrated in Table 1 and reveal a total of 1,686 unique variants for *SLC3A1* and 1,274 unique variants for *SLC7A9* (Table 1 and Supplemental Tables 1 & 2). The vast majority of unique variants in 1KG were found to be single nucleotide variations (SNV) affecting non-coding sequences in introns (84.7% for *SLC3A1* and 88.2% for *SLC7A9*) (Table 1). Further, nucleotide insertions and deletions were exclusively observed in the intron regions and constitute 1-4 % of the total number of unique variants in both genes (Table 1). Interestingly, no nonsense variants were identified in the 1KG database for both *SLC3A1* and *SLC7A9* genes (Table 1), suggesting that nonsense variants are strictly deleterious and unnatural to have in healthy individuals.

HGMD represents a wide-ranging collection of pathogenic variants that underlie or closely correlate with human diseases [16]. A total of 110 and 85 unique pathogenic variants for *SLC3A1* and *SLC7A9*, respectively, were identified. These include missense variants, nonsense variants, insertions, deletions, and complex substitutions (Table 2). In contrast to 1KG variants, almost all reported disease-causing variants in HGMD were observed in coding regions with very little documentation of variants occurring in intron or untranslated regions (Table 2). Non-synonymous missense variants constituted the great majority of variants (73.6% for *SLC3A1* and 51.7% for *SLC7A9*). On the other hand, synonymous variants were only observed in *SLC7A9* (8.2%). Moreover, nucleotide insertions and deletions were found to contribute to more frameshift than in-frame variants in both genes (Table 2). Two complex substitutions were exclusively identified in *SLC3A1* in which multiple consecutive nucleotides were significantly altered (Table 2).

After intersecting the variants identified in the HGMD and 1KG database, only 14 unique disease-causing variants were present in both datasets (Figure1, Table 3). Of the pathogenic variants present in HGMD, only 9 unique *SLC3A1* pathogenic variants were found to be carried by 26 people and 5 unique *SLC7A9* pathogenic variants were found to be carried by 12 people from the 2,504 healthy non-related individuals in 1KG (Figure1, Table 3). Notably, all identified individuals were found to be heterozygote carriers. There were no homozygotes, compound heterozygotes, multiple pathogenic variants in cis or trans, double homozygotes, or double heterozygotes. Detailed characterization of these 14 unique variants including position, nucleotides and amino acids change, nomenclature, loci, and the number of people carrying each variant are summarized in Table 3. Remarkably, all the intersected variants were missense variants.

Based on these findings, the disease-causing alleles were found to have a frequency of 0.52% for *SLC3A1*, and 0.24% for *SLC7A9* (Table 3, Table 4). The missense variants c.797T>C (p.Phe266Ser) for *SLC3A1* and c.544G>A (p.Ala182Thr) for *SLC7A9* were found to be the most common cystine stone causing variants in 1KG database with frequencies of 0.14% and 0.1%, respectively (Table 3). Using the Hardy Weinberg Equilibrium (p^2^ + 2pq + q^2^ = 1), where q is the frequency of disease-causing variant, the expected carrier rates for cystine stones caused by *SLC3A1* and *SLC7A9* pathogenic variants were 1 in 96 and 1 in 209, respectively (Table 4). The combined expected carrier rate for both genes is 1 in 66 (Table 4). Similarly, the expected affected rates (q^2^) summarized in table 4 were shown to be 1 in 37,100 for *SLC3A1* pathogenic variants, 1 in 174,167 for *SLC7A9* pathogenic variants, and 1 in 30,585 for the cumulative rate of combining both genes (Table 4).

## Discussion

Our study estimated the prevalence of cystine stones caused by *SLC3A1* and *SLC7A9* pathogenic variants using a genetic approach that intersected *SLC3A1* and *SLC7A9* gene variants from HGMD and 1KG databases. Among 110 *SLC3A1* and 85 *SLC7A9* unique pathogenic variants in the HGMD (Table 2), a total of 14 unique variants (9 unique variants for *SLC3A1* and 5 unique variants for *SLC7A9*), all missense mutations, were found to be present in 1KG database with a cumulative frequency of 0.75% (Table 4).

### De Novo Mutations vs. Negative Selection

Only 9 out of 110 unique *SLC3A1* pathogenic variants were found in the 1KG. Similarly, only 5 out of 85 unique *SLC7A9* pathogenic variants were identified. This suggests that *SLC3A1*/*SLC7A9* pathogenic variants are not commonly carried by the general population. This phenomenon can be explained by either de novo mutations that have not yet risen to high frequencies or a negative selection against people carrying these pathogenic variants. Longitudinal studies evaluating pathogenic variants and their prevalence across time are required to better understand this phenomenon. An increase in the number of pathogenic variants or the clinical prevalence of cystine stones with time would suggest that multiple de novo mutations in *SLC3A1*/*SLC7A9* have occurred in the general population. This may point towards a possible mutation hotspot in *SLC3A1*/*SLC7A9* genes or the presence of a triggering environmental mutagen. On the contrary, if the observed clinical prevalence of cystine stones stays the same, or even trends down with time, then this would support the notion of a negative selection occurring in the general population against individuals carrying *SLC3A1*/*SLC7A9* pathogenic variants. This would require in-depth research to understand the potential disadvantages and detriments conferred by these pathogenic variants on their carriers. Additionally, defects of *SLC3A1/SLC7A9* will cause leaks of other dibasic amino acids through urine, such as lysine, arginine, and ornithine [2]. Although these compounds do not form kidney stones, the clinical significance of the loss may need to be further explored.

### Discrepancy between Clinical and Genetic Prevalence

The worldwide prevalence of cystinuria is reported to be 1 in 7,000 individuals, however, variation by geographical location happens to be very significant (e.g. 1 per 2,000 in Spain, 1 per 4,000 in Australia, 1 per 18,000 in Japan) [17, 18]. The clinically observed prevalence in the general population is considerably higher than the genetic approach used in our study which estimated the overall affected rate of cystine stone disease to be 1 in 30,585. This indicates the etiology of cystine stone is far more than what our current genetic knowledge can explain. Possible causes for this discrepancy include undiscovered novel causal genes, pathogenic variants, alternative inheritance models, founder effect, or other modification factors.

Indeed, our current knowledge of cystine stone pathogenesis is far from complete. No pathogenic variants were detected in *SLC3A1* or *SLC7A9* in at least 5-10% of patients with clinically confirmed cystine stones [19, 20]. In other words, it is possible that genes other than *SLC3A1* and *SLC7A9* are implicated in the pathogenesis of cystine stones [21, 22].

Additionally, many identified variants in patients with cystine stones were uncharacterized or of unknown clinical significance as their role in the disease process was unclear [11]. This observation has been corroborated by several reports suggesting that our unawareness of many potential cystine stone-associated variants might account for the discrepancy between the genetic and clinical prevalence [23, 24]). It has been proposed that many of these unrecognized pathogenic variants escape detection as they lie deep in the intronic regions potentially affecting splice sites, 5’ promoter regions, and 3’ polyadenylation regions without actually altering the peptide sequence [22].

Another potential explanation for the difference between the genetic and clinical prevalence of cystine stone disease is the possibility of an alternate mode of inheritance. Although cystine stone disease is considered an autosomal recessive phenotype, cystinuria (without cystine stones) may exhibit an autosomal dominant pattern of inheritance with incomplete penetrance. These cases have been notably reported in heterozygote carriers of *SLC7A9* pathogenic variants [22]. This is suggested by the higher than normal urinary levels of cystine and late-onset disease observed in obligate heterozygotes [25]. Despite a low risk of nephrolithiasis, the wide variability in clinical presentation and disease course seen in these individuals warrant further research to better understand their risk profile.

The high clinical prevalence of cystinuria observed in some populations may be due to a founder effect, a reduced genetic diversity that happens when a community is founded by a small number of inhabiting ancestors and has not recruited newcomers [26]. This is exceptionally true in Libyan Jewish populations in which the clinical prevalence of cystinuria is as high as 1 in 2,500 [27].

Another feature of cystine stone disease that has been highlighted by many studies is the wide variability of phenotypes seen in individuals carrying the same genetic variant. This has been demonstrated by the different disease severities, frequency of stone episodes, and age of onset, even in families carrying the same pathogenic variant [23, 28]. For example, in the International Cystinuria Consortium cohort, it was noted that 15-30% of individuals started forming renal calculi before the age of 3. On the other hand, around 6% of the patients with homozygous pathogenic variants in *SLC3A1* and *SLC7A9* were stone-free [29]. This broad variable expressivity suggests that other factors such as epigenetic modification, environmental factors, or modifier genes might contribute to the presentation of the disease [30].

### Strengths and Limitations

Our study has several strengths and limitations. Our biggest strength is utilizing a human-curated database, HGMD professional (https://www.hgmd.cf.ac.uk/ac/index.php), to assess the pathogenicity of each variant of two genes. Another strength is that we utilized the population database that provides individual-level genome data, 1KG (https://www.genome.gov/27528684/1000-genomes-project), to assess the carrier and allele frequencies. Compared to other databases that provide aggregate data, we were able to confirm that there are no homozygotes, no compound heterozygotes, or double heterozygotes.

One limitation we have is that the HGMD professional version is proprietary, and is not available to the general public. In this study, we used the disease-causing variants that were documented in HGMD back in 2016. Therefore, novel pathogenic variants that were discovered after 2016 were not included. However, as discussed, longitudinal studies across different time points are needed to evaluate potential de novo mutations or negative selection processes. Our findings mark a reference point that future epidemiological studies can use to test our proposed hypotheses behind the observed discrepancy between the genetic and clinical prevalence of cystine stone disease.

## Conclusion

Our study aims to estimate the genetic prevalence of cystine stones disease. Among the 110 and 85 pathogenic variants for *SLC3A1* and *SLC7A9*, respectively, only 14 unique disease-causing variants were carried in the general population in the 1KG database. The observed clinical prevalence of cystine stone disease (1 in 7,000) is much higher than the overall genetic prevalence caused by disease-causing variants in *SLC3A1* and *SLC7A9* (1 in 30,585), indicating the etiology of cystine stone is far more than what our current genetic knowledge can explain. This difference may be explained by novel causal genes, undiscovered pathogenic variants, alternative inheritance models, founder effect, epigenetic modification, environmental factors, modifier genes, or other modification factors. There could also be undiscovered de novo mutations or negative selection processes for *SLC3A1* and *SLC7A9*. Our results shed light on this knowledge gap and call for further research to identify further factors that are implicated in the pathogenesis of cystine stones.

## Data Availability

All data produced in the present study are available upon reasonable request to the authors

https://www.internationalgenome.org/

## Acknowledgements

We gratefully acknowledge the invaluable and unwavering support provided by the University of Colorado School of Medicine, Harvard Medical School, and Case Western Reserve University and University Hospitals. Their resources have been instrumental in the advancement of our research, enabling us to pursue discoveries with confidence and rigor.

## Ethics approval statement

### Disclosure of potential conflicts of interes

No funding was received to assist with the preparation of this manuscript. The authors have no competing interests to declare that are relevant to the content of this article.

### Research involving Human Participants and/or Animals

This is an observational study. The data used is de-identified and publicly available. The Case Western Reserve University/ University Hospital institutional review board has confirmed that no ethical approval is required.

### Informed consent

Given the deidentified nature of the data, the Case Western Reserve University/ University Hospital institutional review board determined that this observational study did not constitute human participants research and was thus exempted from review and the need for informed consent, in accordance with 45 CFR §46.

### Contributorship Statement

C.W W: Project development, Data Collection, Manuscript writing

J B: Data collection, Manuscript writing

J C : Data analysis, Manuscript editing

YR H: Visualization

FJ K: Manuscript editing

T W: Manuscript editing

A T: Manuscript editing

N M: Data collection

RD S: Manuscript editing

WR M: Manuscript editing

FR S: Data analysis

