## supplementary table 1 for "Population Genetics Analysis of *SLC3A1* and *SLC7A9* Revealed the Etiology of Cystine Stone May Be More Than What Our Current Genetic Knowledge Can Explain"

**Supplementary Table 1. Variants of SLC3A1 in the 1KG database**

| Chr | Start Position | End Position | Variant type | Ref Genotype \| Ref Proteotype | Variant Genotype 1\| Variant Proteotype 1 | Count | Variant Genotype 2\| Variant Proteotype 2 | Count |
|---|---|---|---|---|---|---|---|---|
| **chr2** | 44502708 | 44502708 | SNV | G\|E | A:G\|K:E | 2 | | 0 |
| **chr2** | 44502788 | 44502788 | SNV | A\|G | C:C\|G:G | 1325 | A:C\|G:G | 873 |
| **chr2** | 44502872 | 44502872 | SNV | G\|A | A:G\|A:A | 2 | | 0 |
| **chr2** | 44503179 | 44503179 | SNV | A | A:G | 2 | | 0 |
| **chr2** | 44503430 | 44503430 | SNV | C | A:C | 17 | | 0 |
| **chr2** | 44503642 | 44503642 | SNV | A | A:G | 422 | G:G | 35 |
| **chr2** | 44503751 | 44503762 | deletion | GCTACTTGGGAG | -:- | 111 | -:GCTACTTGGGAG | 668 |
| **chr2** | 44503775 | 44503775 | SNV | G | C:G | 6 | | 0 |
| **chr2** | 44503832 | 44503832 | SNV | C | C:T | 37 | T:T | 1 |
| **chr2** | 44504129 | 44504129 | SNV | G | T:T | 4 | G:T | 128 |
| **chr2** | 44504134 | 44504134 | SNV | G | C:G | 81 | C:C | 3 |
| **chr2** | 44504169 | 44504169 | SNV | C | A:C | 21 | | 0 |
| **chr2** | 44504176 | 44504176 | SNV | C | A:C | 73 | A:A | 3 |
| **chr2** | 44504240 | 44504240 | insertion | - | -:C | 1106 | C:C | 877 |
| **chr2** | 44504280 | 44504280 | deletion | C | -:C | 3 | | 0 |
| **chr2** | 44504372 | 44504372 | SNV | G | A:G | 9 | | 0 |
| **chr2** | 44504438 | 44504438 | SNV | G | A:G | 1 | | 0 |
| **chr2** | 44504465 | 44504465 | SNV | C | T:T | 4 | C:T | 129 |
| **chr2** | 44504495 | 44504495 | SNV | C | A:C | 165 | A:A | 12 |
| **chr2** | 44504536 | 44504536 | SNV | C | C:T | 1 | | 0 |
| **chr2** | 44504558 | 44504558 | SNV | G | A:A | 4 | A:G | 130 |
| **chr2** | 44504587 | 44504587 | SNV | A | A:G | 19 | | 0 |
| **chr2** | 44504748 | 44504748 | insertion | - | -:C | 7 | | 0 |
| **chr2** | 44504857 | 44504857 | SNV | C | C:T | 307 | T:T | 13 |
| **chr2** | 44504877 | 44504877 | SNV | T | C:T | 960 | C:C | 1197 |
| **chr2** | 44505149 | 44505149 | SNV | C | A:C | 17 | | 0 |
| **chr2** | 44505185 | 44505185 | SNV | T | G:T | 805 | G:G | 1410 |
| **chr2** | 44505321 | 44505321 | SNV | C | T:T | 2 | C:T | 82 |
| **chr2** | 44505329 | 44505329 | SNV | A | A:G | 26 | | 0 |
| **chr2** | 44505429 | 44505429 | insertion | - | -:A | 11 | | 0 |
| **chr2** | 44505457 | 44505457 | SNV | G | C:G | 18 | | 0 |
| **chr2** | 44505574 | 44505574 | SNV | G | A:G | 9 | | 0 |
| **chr2** | 44505588 | 44505588 | insertion | - | -:AAAT | 22 | | 0 |
| **chr2** | 44505608 | 44505608 | insertion | - | -:AAATA | 17 | | 0 |
| **chr2** | 44505879 | 44505879 | SNV | G | A:A | 1 | A:G | 79 |
| **chr2** | 44505890 | 44505890 | SNV | A | A:G | 15 | | 0 |
| **chr2** | 44506089 | 44506089 | SNV | T | C:T | 31 | C:C | 2 |
| **chr2** | 44506145 | 44506145 | SNV | T | A:T | 589 | A:A | 49 |
| **chr2** | 44506256 | 44506256 | SNV | G | A:G | 219 | A:A | 20 |
| **chr2** | 44506345 | 44506345 | SNV | G | A:G | 1193 | A:A | 643 |
| **chr2** | 44506368 | 44506368 | SNV | A | A:T | 163 | T:T | 11 |
| **chr2** | 44506740 | 44506740 | SNV | T | C:T | 238 | C:C | 21 |
| **chr2** | 44506856 | 44506856 | SNV | T | C:T | 1 | | 0 |
| **chr2** | 44506921 | 44506921 | SNV | C | C:T | 1201 | T:T | 652 |
| **chr2** | 44506971 | 44506971 | SNV | G | C:C | 641 | C:G | 1200 |
| **chr2** | 44507074 | 44507074 | SNV | G | A:G | 19 | | 0 |
| **chr2** | 44507213 | 44507213 | SNV | C | T:T | 572 | C:T | 1148 |
| **chr2** | 44507239 | 44507239 | SNV | G | C:G | 45 | | 0 |
| **chr2** | 44507598 | 44507598 | SNV | G | A:G | 32 | A:A | 1 |
| **chr2** | 44507657 | 44507657 | SNV | G | A:G | 1 | | 0 |
| **chr2** | 44507672 | 44507672 | SNV | C | C:T | 2 | | 0 |
| **chr2** | 44507673 | 44507673 | SNV | G | A:G | 1153 | A:A | 568 |
| **chr2** | 44507820 | 44507820 | SNV | T | C:T | 4 | | 0 |
| **chr2** | 44507990 | 44507990 | SNV | C\|T | C:T\|T:M | 6 | | 0 |
| **chr2** | 44508095 | 44508095 | SNV | T | C:C | 4 | C:T | 80 |
| **chr2** | 44508142 | 44508142 | insertion | - | -:TCCC | 372 | TCCC:TCCC | 91 |
| **chr2** | 44508180 | 44508180 | SNV | G | A:G | 3 | | 0 |
| **chr2** | 44508181 | 44508181 | SNV | C | C:T | 70 | T:T | 3 |
| **chr2** | 44508193 | 44508193 | SNV | T | G:T | 207 | G:G | 18 |
| **chr2** | 44508203 | 44508203 | SNV | C | G:G | 2 | C:G | 70 |
| **chr2** | 44508251 | 44508251 | SNV | C | C:G | 11 | | 0 |
| **chr2** | 44508298 | 44508298 | insertion | - | T:T | 365 | -:T | 1602 |
| **chr2** | 44508695 | 44508695 | SNV | G | A:G | 12 | | 0 |
| **chr2** | 44508835 | 44508835 | SNV | G | A:G | 1180 | A:A | 806 |
| **chr2** | 44508932 | 44508935 | deletion | CCTT | -:CCTT | 12 | | 0 |
| **chr2** | 44508951 | 44508951 | SNV | C | C:T | 8 | | 0 |
| **chr2** | 44509044 | 44509044 | SNV | G | G:T | 3 | | 0 |
| **chr2** | 44509155 | 44509155 | SNV | C | C:T | 67 | T:T | 0 |
| **chr2** | 44509160 | 44509160 | SNV | G | A:G | 54 | | 0 |
| **chr2** | 44509276 | 44509276 | SNV | G | A:G | 7 | | 0 |
| **chr2** | 44509362 | 44509362 | SNV | T | C:T | 31 | | 0 |
| **chr2** | 44509564 | 44509564 | SNV | C | A:C | 144 | A:A | 6 |
| **chr2** | 44509653 | 44509653 | SNV | C | C:G | 1 | | 0 |
| **chr2** | 44509760 | 44509760 | SNV | A | A:C | 1 | | 0 |
| **chr2** | 44509762 | 44509762 | SNV | C | C:T | 8 | | 0 |
| **chr2** | 44509924 | 44509924 | SNV | G | C:G | 1 | | 0 |
| **chr2** | 44510104 | 44510104 | SNV | T | C:T | 1 | | 0 |
| **chr2** | 44510146 | 44510146 | SNV | A | T:T | 891 | A:T | 1154 |
| **chr2** | 44510179 | 44510179 | SNV | T | C:T | 922 | C:C | 1242 |
| **chr2** | 44510196 | 44510196 | SNV | G | T:T | 35 | G:T | 269 |
| **chr2** | 44510214 | 44510214 | SNV | C | C:T | 5 | | 0 |
| **chr2** | 44510270 | 44510270 | SNV | C | C:T | 2 | | 0 |
| **chr2** | 44510557 | 44510557 | SNV | C | C:G | 2 | | 0 |
| **chr2** | 44510578 | 44510578 | SNV | C | T:T | 3 | C:T | 70 |
| **chr2** | 44510758 | 44510758 | deletion | T | -:T | 11 | | 0 |
| **chr2** | 44510861 | 44510861 | SNV | C | A:C | 24 | | 0 |
| **chr2** | 44510936 | 44510936 | SNV | C | C:T | 159 | T:T | 9 |
| **chr2** | 44511152 | 44511152 | deletion | T | -:T | 22 | | 0 |
| **chr2** | 44511155 | 44511155 | SNV | T | A:T | 4 | | 0 |
| **chr2** | 44511198 | 44511198 | SNV | A | A:G | 6 | | 0 |
| **chr2** | 44511391 | 44511391 | SNV | G | C:G | 5 | | 0 |
| **chr2** | 44511667 | 44511667 | SNV | A | T:T | 3 | A:T | 70 |
| **chr2** | 44511787 | 44511787 | SNV | G | G:T | 33 | | 0 |
| **chr2** | 44511806 | 44511806 | SNV | A | G:G | 12 | A:G | 149 |
| **chr2** | 44511861 | 44511861 | SNV | T | C:C | 1118 | C:T | 1005 |

**Supplementary Table 1. Variants of SLC3A1 in the 1KG database**

| Chr | Start Position | End Position | Variant type | Ref Genotype \| Ref Proteotype | Variant Genotype 1\| Variant Proteotype 1 | Count | Variant Genotype 2\| Variant Proteotype 2 | Count |
|---|---|---|---|---|---|---|---|---|
| **chr2** | 44511929 | 44511929 | SNV | T | C:T | 1177 | C:C | 797 |
| **chr2** | 44512162 | 44512162 | SNV | C | C:T | 4 | | 0 |
| **chr2** | 44512524 | 44512526 | deletion | ATT | -:ATT | 19 | | 0 |
| **chr2** | 44512579 | 44512579 | SNV | C | C:T | 33 | | 0 |
| **chr2** | 44512797 | 44512797 | SNV | T | C:T | 26 | | 0 |
| **chr2** | 44512952 | 44512952 | SNV | G | A:G | 72 | A:A | 3 |
| **chr2** | 44512970 | 44512970 | SNV | C | C:T | 5 | | 0 |
| **chr2** | 44513087 | 44513087 | SNV | G | A:G | 36 | | 0 |
| **chr2** | 44513135 | 44513135 | SNV | A | A:G | 48 | | 0 |
| **chr2** | 44513202 | 44513202 | SNV | T\|F | C:T\|S:F | 7 | | 0 |
| **chr2** | 44513704 | 44513704 | SNV | C | C:T | 12 | | 0 |
| **chr2** | 44513721 | 44513721 | SNV | C | C:T | 3 | | 0 |
| **chr2** | 44513872 | 44513872 | SNV | A | G:G | 9 | A:G | 177 |
| **chr2** | 44513890 | 44513890 | SNV | G | A:G | 146 | | 0 |
| **chr2** | 44514271 | 44514271 | SNV | C | C:G | 47 | | 0 |
| **chr2** | 44514438 | 44514438 | SNV | T | C:C | 11 | C:T | 144 |
| **chr2** | 44514481 | 44514481 | SNV | C | C:T | 1 | | 0 |
| **chr2** | 44514547 | 44514547 | SNV | G | A:G | 1 | | 0 |
| **chr2** | 44514844 | 44514844 | SNV | C | C:G | 10 | | 0 |
| **chr2** | 44514862 | 44514862 | SNV | A | A:G | 1 | | 0 |
| **chr2** | 44514897 | 44514897 | SNV | G | A:G | 146 | A:A | 6 |
| **chr2** | 44515068 | 44515068 | SNV | A | A:G | 97 | G:G | 3 |
| **chr2** | 44515084 | 44515084 | SNV | C | T:T | 20 | C:T | 228 |
| **chr2** | 44515085 | 44515085 | SNV | A | G:G | 1 | A:G | 49 |
| **chr2** | 44515257 | 44515257 | SNV | C | T:T | 385 | C:T | 847 |
| **chr2** | 44515538 | 44515538 | SNV | C | C:T | 32 | | 0 |
| **chr2** | 44515579 | 44515579 | SNV | C | C:T | 118 | T:T | 11 |
| **chr2** | 44515611 | 44515611 | SNV | T | C:T | 4 | | 0 |
| **chr2** | 44515656 | 44515656 | SNV | T | A:T | 33 | | 0 |
| **chr2** | 44515668 | 44515668 | SNV | C | T:T | 20 | C:T | 228 |
| **chr2** | 44515764 | 44515764 | SNV | C | C:T | 3 | | 0 |
| **chr2** | 44515851 | 44515851 | SNV | C | T:T | 3 | C:T | 115 |
| **chr2** | 44515859 | 44515859 | SNV | G | A:G | 15 | | 0 |
| **chr2** | 44516087 | 44516087 | SNV | C | A:A | 24 | A:C | 220 |
| **chr2** | 44516088 | 44516088 | SNV | C | A:C | 220 | A:A | 24 |
| **chr2** | 44516120 | 44516120 | SNV | C | C:T | 10 | | 0 |
| **chr2** | 44516161 | 44516162 | deletion | AT | -:- | 1 | -:AT | 45 |
| **chr2** | 44516282 | 44516282 | SNV | A | G:G | 1276 | A:G | 875 |
| **chr2** | 44516351 | 44516351 | SNV | T | G:T | 1 | | 0 |
| **chr2** | 44516399 | 44516399 | SNV | G | C:G | 1 | | 0 |
| **chr2** | 44516411 | 44516411 | SNV | G | A:G | 3 | | 0 |
| **chr2** | 44516631 | 44516631 | SNV | T | C:T | 210 | C:C | 17 |
| **chr2** | 44516767 | 44516770 | deletion | TATT | -:- | 3 | -:TATT | 90 |
| **chr2** | 44516892 | 44516892 | SNV | C | A:A | 10 | A:C | 147 |
| **chr2** | 44516916 | 44516916 | SNV | C | T:T | 2 | C:T | 65 |
| **chr2** | 44516961 | 44516961 | SNV | C | C:T | 2 | | 0 |
| **chr2** | 44517017 | 44517017 | SNV | G | A:G | 2 | | 0 |
| **chr2** | 44517034 | 44517034 | SNV | T | A:A | 1 | A:T | 29 |
| **chr2** | 44517036 | 44517036 | SNV | C | G:G | 1 | C:G | 29 |
| **chr2** | 44517072 | 44517072 | SNV | A | C:C | 759 | A:C | 1183 |
| **chr2** | 44517080 | 44517080 | insertion | - | -:T | 288 | T:T | 16 |
| **chr2** | 44517148 | 44517148 | SNV | A | A:G | 238 | G:G | 22 |
| **chr2** | 44517190 | 44517190 | SNV | T | A:T | 215 | A:A | 21 |
| **chr2** | 44517204 | 44517204 | SNV | T | G:T | 1 | | 0 |
| **chr2** | 44517262 | 44517262 | insertion | - | -:T | 1404 | T:T | 316 |
| **chr2** | 44517304 | 44517304 | SNV | A | A:G | 2 | | 0 |
| **chr2** | 44517476 | 44517476 | SNV | T | G:G | 4 | G:T | 97 |
| **chr2** | 44517559 | 44517559 | SNV | G | A:G | 20 | | 0 |
| **chr2** | 44517576 | 44517576 | SNV | C | C:T | 4 | | 0 |
| **chr2** | 44517662 | 44517662 | SNV | T | C:T | 1180 | C:C | 757 |
| **chr2** | 44517704 | 44517704 | SNV | G | A:G | 1 | | 0 |
| **chr2** | 44517722 | 44517722 | SNV | G | A:G | 40 | A:A | 1 |
| **chr2** | 44517752 | 44517752 | SNV | G | A:G | 1185 | A:A | 679 |
| **chr2** | 44517759 | 44517759 | SNV | T | C:T | 73 | C:C | 3 |
| **chr2** | 44517822 | 44517822 | insertion | - | -:T | 931 | T:T | 1203 |
| **chr2** | 44517839 | 44517839 | SNV | G | A:A | 1204 | A:G | 929 |
| **chr2** | 44517933 | 44517933 | SNV | T | C:T | 2 | | 0 |
| **chr2** | 44518338 | 44518338 | SNV | G | C:G | 144 | C:C | 6 |
| **chr2** | 44518559 | 44518559 | SNV | G | A:A | 10 | A:G | 156 |
| **chr2** | 44518752 | 44518752 | SNV | G | T:T | 5 | G:T | 139 |
| **chr2** | 44518786 | 44518786 | SNV | C | C:T | 20 | | 0 |
| **chr2** | 44518866 | 44518866 | SNV | G | A:G | 3 | | 0 |
| **chr2** | 44519073 | 44519073 | SNV | T | C:T | 13 | | 0 |
| **chr2** | 44519142 | 44519142 | SNV | A | G:G | 980 | A:G | 1077 |
| **chr2** | 44519290 | 44519290 | SNV | C | C:T | 89 | T:T | 6 |
| **chr2** | 44519544 | 44519544 | SNV | C | C:G | 153 | G:G | 12 |
| **chr2** | 44519619 | 44519619 | SNV | G | C:G | 1 | | 0 |
| **chr2** | 44519713 | 44519713 | SNV | C | T:T | 1 | C:T | 69 |
| **chr2** | 44519751 | 44519751 | SNV | C | C:T | 31 | | 0 |
| **chr2** | 44519769 | 44519769 | SNV | T | C:C | 11 | C:T | 122 |
| **chr2** | 44519906 | 44519909 | deletion | ATAT | -:- | 3 | -:ATAT | 78 |
| **chr2** | 44519981 | 44519981 | SNV | C | C:T | 1 | | 0 |
| **chr2** | 44520053 | 44520053 | SNV | C | C:T | 21 | | 0 |
| **chr2** | 44520285 | 44520285 | SNV | G | C:G | 10 | | 0 |
| **chr2** | 44520657 | 44520657 | deletion | G | -:G | 1106 | -:- | 859 |
| **chr2** | 44520662 | 44520662 | SNV | G | A:A | 105 | A:G | 573 |
| **chr2** | 44520716 | 44520716 | SNV | A | A:G | 932 | G:G | 1177 |
| **chr2** | 44520774 | 44520775 | deletion | GA | -:GA | 4 | | 0 |
| **chr2** | 44521098 | 44521098 | insertion | - | G:G | 1468 | -:G | 746 |
| **chr2** | 44521107 | 44521107 | SNV | C | C:G | 2 | | 0 |
| **chr2** | 44521146 | 44521146 | SNV | G | A:G | 5 | | 0 |
| **chr2** | 44521176 | 44521176 | SNV | C | C:T | 6 | | 0 |
| **chr2** | 44521246 | 44521246 | SNV | A | G:G | 1473 | A:G | 741 |
| **chr2** | 44521544 | 44521544 | SNV | T | C:C | 1407 | C:T | 785 |

**Supplementary Table 1. Variants of SLC3A1 in the 1KG database**

| Chr | Start Position | End Position | Variant type | Ref Genotype \| Ref Proteotype | Variant Genotype 1\| Variant Proteotype 1 | Count | Variant Genotype 2\| Variant Proteotype 2 | Count |
|---|---|---|---|---|---|---|---|---|
| **chr2** | 44521608 | 44521608 | SNV | C | C:T | 19 | | 0 |
| **chr2** | 44521642 | 44521646 | deletion | AAAAA | -:AAAAA | 5 | | 0 |
| **chr2** | 44521764 | 44521764 | SNV | A | G:G | 1 | A:G | 19 |
| **chr2** | 44521978 | 44521978 | SNV | C | A:C | 1 | | 0 |
| **chr2** | 44522280 | 44522280 | SNV | G | A:G | 28 | | 0 |
| **chr2** | 44522449 | 44522449 | SNV | G | A:G | 1 | | 0 |
| **chr2** | 44522522 | 44522522 | SNV | C | C:G | 1 | | 0 |
| **chr2** | 44522560 | 44522560 | SNV | G | C:G | 15 | C:C | 2 |
| **chr2** | 44522624 | 44522624 | SNV | T | G:T | 1 | | 0 |
| **chr2** | 44522777 | 44522777 | SNV | A | A:T | 1 | | 0 |
| **chr2** | 44522805 | 44522805 | SNV | C | C:T | 19 | T:T | 1 |
| **chr2** | 44522822 | 44522822 | SNV | G | A:G | 32 | | 0 |
| **chr2** | 44522965 | 44522965 | SNV | G | G:T | 2 | | 0 |
| **chr2** | 44523059 | 44523059 | SNV | G | A:G | 2 | | 0 |
| **chr2** | 44523787 | 44523787 | SNV | T | C:T | 3 | | 0 |
| **chr2** | 44523908 | 44523908 | SNV | A | A:T | 29 | T:T | 2 |
| **chr2** | 44523966 | 44523966 | SNV | C | C:G | 3 | | 0 |
| **chr2** | 44523968 | 44523968 | SNV | T | G:T | 21 | | 0 |
| **chr2** | 44524064 | 44524064 | SNV | G | A:G | 1045 | A:A | 581 |
| **chr2** | 44524575 | 44524575 | SNV | T | C:C | 757 | C:T | 1179 |
| **chr2** | 44524603 | 44524603 | SNV | A | A:G | 5 | | 0 |
| **chr2** | 44524630 | 44524630 | SNV | T | A:T | 85 | A:A | 6 |
| **chr2** | 44524735 | 44524735 | SNV | G | A:G | 1 | | 0 |
| **chr2** | 44524765 | 44524765 | SNV | C | C:T | 34 | | 0 |
| **chr2** | 44524899 | 44524899 | SNV | C | A:C | 27 | | 0 |
| **chr2** | 44524936 | 44524936 | SNV | T | A:A | 1 | A:T | 73 |
| **chr2** | 44524945 | 44524945 | SNV | C | T:T | 16 | C:T | 205 |
| **chr2** | 44525023 | 44525023 | SNV | C | C:T | 4 | | 0 |
| **chr2** | 44525239 | 44525239 | SNV | G | C:G | 59 | | 0 |
| **chr2** | 44525299 | 44525299 | deletion | C | -:- | 1 | -:C | 72 |
| **chr2** | 44525327 | 44525327 | deletion | A | -:- | 1169 | -:A | 946 |
| **chr2** | 44525353 | 44525353 | SNV | C | C:G | 4 | | 0 |
| **chr2** | 44525355 | 44525355 | SNV | T | C:C | 1 | C:T | 20 |
| **chr2** | 44525475 | 44525475 | SNV | C | C:T | 1 | | 0 |
| **chr2** | 44525484 | 44525484 | SNV | C | C:T | 128 | T:T | 5 |
| **chr2** | 44525509 | 44525509 | SNV | A | A:G | 120 | G:G | 11 |
| **chr2** | 44525523 | 44525523 | insertion | - | T:T | 91 | -:T | 387 |
| **chr2** | 44525640 | 44525640 | SNV | C | C:T | 10 | | 0 |
| **chr2** | 44525665 | 44525665 | SNV | T | C:C | 25 | C:T | 305 |
| **chr2** | 44525804 | 44525804 | SNV | G | A:A | 67 | A:G | 363 |
| **chr2** | 44525992 | 44525994 | deletion | GAG | -:GAG | 31 | | 0 |
| **chr2** | 44526010 | 44526010 | SNV | G | A:G | 1184 | A:A | 762 |
| **chr2** | 44526073 | 44526073 | SNV | A | A:G | 11 | | 0 |
| **chr2** | 44526302 | 44526302 | SNV | C | C:T | 5 | | 0 |
| **chr2** | 44526336 | 44526336 | SNV | C | C:T | 288 | T:T | 22 |
| **chr2** | 44526405 | 44526405 | SNV | T | C:T | 9 | | 0 |
| **chr2** | 44526429 | 44526429 | SNV | C | C:T | 1 | | 0 |
| **chr2** | 44526531 | 44526531 | SNV | T | C:C | 70 | C:T | 356 |
| **chr2** | 44527069 | 44527069 | SNV | C | C:T | 1 | | 0 |
| **chr2** | 44527104 | 44527104 | SNV | C | C:G | 21 | | 0 |
| **chr2** | 44527274 | 44527274 | SNV | T | C:T | 1 | | 0 |
| **chr2** | 44527421 | 44527421 | SNV | T | A:T | 87 | A:A | 6 |
| **chr2** | 44527432 | 44527432 | SNV | C | T:T | 18 | C:T | 254 |
| **chr2** | 44527664 | 44527664 | SNV | C | A:A | 5 | A:C | 144 |
| **chr2** | 44528033 | 44528033 | SNV | G | A:G | 2 | | 0 |
| **chr2** | 44528078 | 44528078 | SNV | T | G:T | 11 | | 0 |
| **chr2** | 44528083 | 44528083 | SNV | C | T:T | 1 | C:T | 11 |
| **chr2** | 44528111 | 44528111 | SNV | G | A:A | 1 | A:G | 57 |
| **chr2** | 44528165 | 44528165 | SNV | G\|E | A:G\|E:E | 9 | | 0 |
| **chr2** | 44528256 | 44528256 | SNV | G\|G | A:G\|S:G | 7 | | 0 |
| **chr2** | 44528268 | 44528268 | deletion | T | -:- | 66 | -:T | 428 |
| **chr2** | 44528492 | 44528492 | SNV | C | C:T | 11 | T:T | 1 |
| **chr2** | 44528571 | 44528571 | SNV | T | A:T | 181 | A:A | 16 |
| **chr2** | 44528602 | 44528602 | SNV | G | A:A | 5 | A:G | 93 |
| **chr2** | 44528662 | 44528662 | SNV | G | A:G | 2 | | 0 |
| **chr2** | 44528663 | 44528663 | SNV | A | A:G | 10 | | 0 |
| **chr2** | 44528681 | 44528681 | SNV | C | G:G | 17 | C:G | 278 |
| **chr2** | 44528697 | 44528697 | SNV | G | A:A | 16 | A:G | 204 |
| **chr2** | 44528877 | 44528877 | insertion | - | A:A | 544 | -:A | 1198 |
| **chr2** | 44528922 | 44528922 | SNV | C | C:G | 506 | G:G | 1937 |
| **chr2** | 44529034 | 44529034 | insertion | - | -:CT | 34 | | 0 |
| **chr2** | 44529117 | 44529117 | SNV | C | T:T | 1 | C:T | 18 |
| **chr2** | 44529152 | 44529152 | SNV | C | G:G | 1 | C:G | 46 |
| **chr2** | 44529244 | 44529245 | deletion | TT | -:- | 17 | -:TT | 277 |
| **chr2** | 44529254 | 44529254 | SNV | A | A:G | 16 | | 0 |
| **chr2** | 44529428 | 44529428 | SNV | C | C:T | 46 | T:T | 1 |
| **chr2** | 44529449 | 44529453 | deletion | ATTAA | -:- | 5 | -:ATTAA | 145 |
| **chr2** | 44529549 | 44529549 | SNV | T | C:T | 1 | | 0 |
| **chr2** | 44529589 | 44529589 | SNV | A | A:C | 6 | | 0 |
| **chr2** | 44529594 | 44529594 | SNV | G | A:G | 1 | | 0 |
| **chr2** | 44529628 | 44529628 | SNV | T | G:T | 3 | | 0 |
| **chr2** | 44529705 | 44529705 | SNV | A | G:G | 6 | A:G | 92 |
| **chr2** | 44529715 | 44529715 | SNV | C | C:T | 72 | T:T | 1 |
| **chr2** | 44529722 | 44529722 | SNV | C | T:T | 2 | C:T | 62 |
| **chr2** | 44529743 | 44529743 | SNV | T | G:T | 1 | | 0 |
| **chr2** | 44529789 | 44529789 | SNV | T | C:C | 5 | C:T | 144 |
| **chr2** | 44529855 | 44529855 | SNV | C | C:G | 1 | | 0 |
| **chr2** | 44529870 | 44529870 | SNV | G | A:G | 11 | | 0 |
| **chr2** | 44529914 | 44529914 | SNV | A | A:C | 18 | C:C | 1 |
| **chr2** | 44529973 | 44529973 | SNV | G | A:A | 15 | A:G | 276 |
| **chr2** | 44530084 | 44530084 | SNV | T | A:A | 22 | A:T | 287 |
| **chr2** | 44530156 | 44530156 | SNV | G | A:G | 1 | | 0 |
| **chr2** | 44530224 | 44530224 | SNV | T | C:T | 8 | | 0 |
| **chr2** | 44530321 | 44530326 | deletion | TAAGAG | -:TAAGAG | 288 | -:- | 22 |

**Supplementary Table 1. Variants of SLC3A1 in the 1KG database**

| Chr | Start Position | End Position | Variant type | Ref Genotype \| Ref Proteotype | Variant Genotype 1\| Variant Proteotype 1 | Count | Variant Genotype 2\| Variant Proteotype 2 | Count |
|---|---|---|---|---|---|---|---|---|
| **chr2** | 44530384 | 44530384 | insertion | - | -:TT | 1149 | TT:TT | 855 |
| **chr2** | 44530408 | 44530408 | SNV | A | A:C | 4 | | 0 |
| **chr2** | 44530423 | 44530423 | SNV | C | T:T | 993 | C:T | 1071 |
| **chr2** | 44530605 | 44530605 | SNV | A | A:T | 356 | T:T | 39 |
| **chr2** | 44530623 | 44530623 | SNV | G | A:A | 17 | A:G | 277 |
| **chr2** | 44530682 | 44530683 | deletion | TG | -:- | 5 | -:TG | 92 |
| **chr2** | 44530761 | 44530761 | SNV | G | A:G | 3 | | 0 |
| **chr2** | 44530827 | 44530827 | SNV | G | A:A | 1 | A:G | 56 |
| **chr2** | 44530894 | 44530894 | SNV | G | A:G | 70 | A:A | 1 |
| **chr2** | 44530929 | 44530929 | SNV | C | C:G | 51 | G:G | 1 |
| **chr2** | 44530931 | 44530931 | SNV | C | C:T | 3 | | 0 |
| **chr2** | 44530940 | 44530940 | SNV | A | T:T | 15 | A:T | 168 |
| **chr2** | 44530945 | 44530945 | SNV | C | C:T | 1 | | 0 |
| **chr2** | 44530948 | 44530948 | SNV | C | T:T | 5 | C:T | 144 |
| **chr2** | 44530965 | 44530965 | SNV | C | C:T | 280 | T:T | 20 |
| **chr2** | 44530985 | 44530985 | SNV | T | C:T | 6 | | 0 |
| **chr2** | 44531071 | 44531071 | SNV | A | A:G | 5 | G:G | 1 |
| **chr2** | 44531081 | 44531081 | SNV | G | T:T | 578 | G:T | 1028 |
| **chr2** | 44531484 | 44531484 | SNV | C | T:T | 764 | C:T | 1186 |
| **chr2** | 44531499 | 44531499 | SNV | G | T:T | 5 | G:T | 145 |
| **chr2** | 44531633 | 44531633 | SNV | G | C:G | 28 | | 0 |
| **chr2** | 44531635 | 44531635 | SNV | T | C:C | 74 | C:T | 383 |
| **chr2** | 44531703 | 44531703 | SNV | C | C:T | 35 | | 0 |
| **chr2** | 44531842 | 44531843 | deletion | TG | -:TG | 22 | | 0 |
| **chr2** | 44531849 | 44531849 | SNV | A | A:T | 1 | | 0 |
| **chr2** | 44531852 | 44531852 | SNV | C | C:T | 6 | | 0 |
| **chr2** | 44531859 | 44531859 | SNV | T | G:T | 12 | G:G | 2 |
| **chr2** | 44531952 | 44531952 | insertion | - | -:T | 108 | T:T | 1 |
| **chr2** | 44532326 | 44532326 | SNV | C | T:T | 31 | C:T | 271 |
| **chr2** | 44532550 | 44532550 | SNV | A | A:C | 10 | | 0 |
| **chr2** | 44532553 | 44532556 | deletion | CTTT | -:- | 5 | -:CTTT | 145 |
| **chr2** | 44532567 | 44532568 | deletion | TT | -:- | 1214 | -:TT | 924 |
| **chr2** | 44532627 | 44532627 | SNV | C | T:T | 2 | C:T | 79 |
| **chr2** | 44532641 | 44532641 | SNV | A | A:C | 3 | | 0 |
| **chr2** | 44532838 | 44532838 | SNV | G | A:G | 178 | A:A | 16 |
| **chr2** | 44532850 | 44532850 | SNV | G | C:G | 17 | | 0 |
| **chr2** | 44532858 | 44532858 | SNV | A | A:C | 12 | | 0 |
| **chr2** | 44532947 | 44532947 | SNV | C | C:G | 10 | | 0 |
| **chr2** | 44532996 | 44532996 | SNV | G | A:G | 5 | | 0 |
| **chr2** | 44533055 | 44533055 | SNV | T | C:T | 1 | | 0 |
| **chr2** | 44533067 | 44533067 | SNV | C | C:T | 2 | | 0 |
| **chr2** | 44533104 | 44533104 | SNV | G | G:T | 9 | | 0 |
| **chr2** | 44533124 | 44533124 | SNV | G | C:G | 50 | C:C | 1 |
| **chr2** | 44533210 | 44533210 | SNV | T | G:T | 10 | | 0 |
| **chr2** | 44533279 | 44533279 | SNV | G | T:T | 4 | G:T | 129 |
| **chr2** | 44533397 | 44533397 | SNV | G | A:G | 144 | A:A | 5 |
| **chr2** | 44533663 | 44533663 | SNV | C | C:T | 95 | T:T | 4 |
| **chr2** | 44533690 | 44533690 | deletion | T | -:- | 526 | -:T | 1234 |
| **chr2** | 44533725 | 44533725 | SNV | G | C:G | 72 | C:C | 1 |
| **chr2** | 44533733 | 44533733 | SNV | C | C:T | 93 | T:T | 4 |
| **chr2** | 44533754 | 44533754 | SNV | G | A:G | 3 | | 0 |
| **chr2** | 44533800 | 44533802 | deletion | CTC | -:CTC | 3 | | 0 |
| **chr2** | 44533946 | 44533946 | SNV | T | C:C | 18 | C:T | 245 |
| **chr2** | 44533965 | 44533965 | SNV | G | A:G | 1 | | 0 |
| **chr2** | 44534012 | 44534012 | SNV | A | A:C | 1059 | C:C | 601 |
| **chr2** | 44534042 | 44534042 | SNV | A | A:T | 2 | | 0 |
| **chr2** | 44534132 | 44534132 | SNV | G | A:G | 1 | | 0 |
| **chr2** | 44534160 | 44534160 | SNV | G | C:G | 5 | | 0 |
| **chr2** | 44534186 | 44534186 | SNV | C | A:C | 1 | | 0 |
| **chr2** | 44534187 | 44534187 | SNV | C | T:T | 1 | C:T | 72 |
| **chr2** | 44534321 | 44534321 | SNV | C | A:C | 882 | A:A | 390 |
| **chr2** | 44534359 | 44534359 | SNV | C | A:C | 275 | A:A | 32 |
| **chr2** | 44534377 | 44534377 | SNV | C | C:T | 1 | | 0 |
| **chr2** | 44534391 | 44534391 | SNV | G | C:G | 1 | | 0 |
| **chr2** | 44534414 | 44534414 | SNV | T | C:T | 108 | C:C | 2 |
| **chr2** | 44534417 | 44534417 | SNV | C | C:T | 697 | T:T | 1516 |
| **chr2** | 44534502 | 44534502 | SNV | T | C:T | 38 | | 0 |
| **chr2** | 44534704 | 44534704 | deletion | T | -:- | 66 | -:T | 1664 |
| **chr2** | 44534852 | 44534852 | SNV | G | T:T | 1 | G:T | 47 |
| **chr2** | 44535034 | 44535034 | SNV | T | C:T | 128 | C:C | 4 |
| **chr2** | 44535069 | 44535069 | SNV | A | A:G | 1 | | 0 |
| **chr2** | 44535288 | 44535288 | SNV | C | C:G | 26 | | 0 |
| **chr2** | 44535347 | 44535347 | SNV | A | G:G | 2 | A:G | 49 |
| **chr2** | 44535365 | 44535365 | SNV | C | C:G | 7 | | 0 |
| **chr2** | 44535413 | 44535413 | SNV | G | A:G | 4 | | 0 |
| **chr2** | 44535454 | 44535454 | SNV | T | A:T | 13 | | 0 |
| **chr2** | 44535467 | 44535467 | SNV | G | C:G | 21 | | 0 |
| **chr2** | 44535519 | 44535519 | SNV | G | C:C | 392 | C:G | 840 |
| **chr2** | 44535643 | 44535645 | deletion | TGT | -:TGT | 7 | | 0 |
| **chr2** | 44535686 | 44535686 | SNV | G | A:A | 1268 | A:G | 877 |
| **chr2** | 44535694 | 44535694 | SNV | C | C:T | 210 | T:T | 16 |
| **chr2** | 44535715 | 44535715 | SNV | T | C:T | 1 | | 0 |
| **chr2** | 44535738 | 44535738 | SNV | G | G:T | 270 | T:T | 27 |
| **chr2** | 44535866 | 44535866 | SNV | G | A:G | 10 | | 0 |
| **chr2** | 44535896 | 44535896 | SNV | G | C:G | 10 | | 0 |
| **chr2** | 44535938 | 44535942 | deletion | CTCAG | -:- | 5 | -:CTCAG | 144 |
| **chr2** | 44535957 | 44535957 | SNV | C | C:T | 5 | | 0 |
| **chr2** | 44535958 | 44535958 | SNV | G | A:G | 265 | A:A | 25 |
| **chr2** | 44535964 | 44535964 | SNV | C | C:T | 5 | | 0 |
| **chr2** | 44535975 | 44535975 | SNV | C | T:T | 1 | C:T | 31 |
| **chr2** | 44535984 | 44535984 | SNV | G | C:G | 22 | | 0 |
| **chr2** | 44536223 | 44536223 | SNV | T | G:T | 1 | | 0 |
| **chr2** | 44536249 | 44536249 | SNV | A | G:G | 627 | A:G | 1104 |
| **chr2** | 44536362 | 44536362 | SNV | C | A:A | 13 | A:C | 217 |

**Supplementary Table 1. Variants of SLC3A1 in the 1KG database**

| Chr | Start Position | End Position | Variant type | Ref Genotype \| Ref Proteotype | Variant Genotype 1\| Variant Proteotype 1 | Count | Variant Genotype 2\| Variant Proteotype 2 | Count |
|---|---|---|---|---|---|---|---|---|
| **chr2** | 44536372 | 44536372 | SNV | C | C:G | 35 | | 0 |
| **chr2** | 44536546 | 44536546 | SNV | T | C:C | 845 | C:T | 1169 |
| **chr2** | 44536634 | 44536634 | deletion | G | -:- | 17 | -:G | 235 |
| **chr2** | 44536677 | 44536677 | SNV | C | T:T | 1 | C:T | 51 |
| **chr2** | 44536704 | 44536704 | SNV | T | C:T | 6 | | 0 |
| **chr2** | 44536706 | 44536706 | SNV | C | C:T | 45 | | 0 |
| **chr2** | 44536716 | 44536716 | SNV | G | G:T | 63 | T:T | 1 |
| **chr2** | 44536757 | 44536757 | SNV | G | A:G | 1 | | 0 |
| **chr2** | 44536807 | 44536807 | SNV | C | T:T | 1 | C:T | 20 |
| **chr2** | 44536821 | 44536821 | SNV | C | G:G | 1 | C:G | 61 |
| **chr2** | 44536893 | 44536893 | insertion | - | -:TGAC | 16 | | 0 |
| **chr2** | 44536928 | 44536928 | SNV | G | G:T | 263 | T:T | 15 |
| **chr2** | 44537028 | 44537028 | SNV | T | G:T | 7 | | 0 |
| **chr2** | 44537036 | 44537036 | insertion | - | T:T | 14 | -:T | 204 |
| **chr2** | 44537145 | 44537145 | SNV | A | A:C | 10 | | 0 |
| **chr2** | 44537157 | 44537157 | SNV | A | A:C | 9 | | 0 |
| **chr2** | 44537222 | 44537222 | deletion | G | -:G | 3 | | 0 |
| **chr2** | 44537288 | 44537288 | SNV | C | C:T | 1 | | 0 |
| **chr2** | 44537296 | 44537296 | SNV | T | A:T | 9 | | 0 |
| **chr2** | 44537397 | 44537397 | SNV | G | A:G | 30 | A:A | 2 |
| **chr2** | 44537422 | 44537422 | SNV | C | C:T | 10 | | 0 |
| **chr2** | 44537568 | 44537571 | deletion | TTCT | -:- | 5 | -:TTCT | 144 |
| **chr2** | 44537586 | 44537586 | SNV | T | C:T | 9 | | 0 |
| **chr2** | 44537702 | 44537702 | SNV | T | C:T | 1 | | 0 |
| **chr2** | 44537742 | 44537742 | SNV | G | A:A | 1 | A:G | 63 |
| **chr2** | 44537821 | 44537821 | SNV | G | A:G | 27 | | 0 |
| **chr2** | 44537828 | 44537828 | SNV | G | G:T | 2 | | 0 |
| **chr2** | 44537829 | 44537829 | SNV | C | A:A | 1 | A:C | 13 |
| **chr2** | 44537831 | 44537831 | insertion | - | T:T | 4 | -:T | 175 |
| **chr2** | 44537916 | 44537916 | SNV | T | C:T | 1 | | 0 |
| **chr2** | 44537978 | 44537978 | SNV | C | C:T | 24 | T:T | 2 |
| **chr2** | 44538080 | 44538080 | SNV | C | C:T | 285 | T:T | 20 |
| **chr2** | 44538237 | 44538239 | deletion | TTC | -:TTC | 44 | | 0 |
| **chr2** | 44538326 | 44538326 | SNV | T | A:T | 1 | | 0 |
| **chr2** | 44538484 | 44538484 | SNV | A | A:G | 864 | G:G | 1291 |
| **chr2** | 44538534 | 44538534 | SNV | T | C:T | 5 | | 0 |
| **chr2** | 44538634 | 44538634 | SNV | G | A:G | 2 | | 0 |
| **chr2** | 44538733 | 44538733 | SNV | G | A:G | 53 | A:A | 1 |
| **chr2** | 44538862 | 44538862 | insertion | - | A:A | 810 | -:A | 1229 |
| **chr2** | 44538876 | 44538876 | SNV | C | A:C | 413 | A:A | 19 |
| **chr2** | 44538929 | 44538929 | SNV | G | G:T | 16 | T:T | 1 |
| **chr2** | 44538932 | 44538932 | SNV | C | C:T | 84 | T:T | 4 |
| **chr2** | 44538936 | 44538936 | SNV | C | T:T | 1 | C:T | 73 |
| **chr2** | 44538963 | 44538963 | SNV | G | A:A | 18 | A:G | 262 |
| **chr2** | 44539009 | 44539009 | SNV | C | T:T | 1270 | C:T | 875 |
| **chr2** | 44539041 | 44539041 | SNV | A | G:G | 939 | A:G | 1098 |
| **chr2** | 44539207 | 44539207 | SNV | C | C:T | 9 | | 0 |
| **chr2** | 44539287 | 44539287 | SNV | T | G:T | 150 | G:G | 5 |
| **chr2** | 44539306 | 44539306 | SNV | T | C:T | 270 | C:C | 52 |
| **chr2** | 44539418 | 44539418 | SNV | T | G:T | 16 | | 0 |
| **chr2** | 44539423 | 44539423 | SNV | C | A:C | 1 | | 0 |
| **chr2** | 44539514 | 44539514 | SNV | C | C:T | 15 | | 0 |
| **chr2** | 44539758 | 44539758 | SNV | C\|R | C:T\|R:C | 1 | | 0 |
| **chr2** | 44539865 | 44539865 | SNV | C\|A | C:T\|A:A | 1 | | 0 |
| **chr2** | 44539913 | 44539913 | insertion | - | -:AGCTATAAATACAAG | 147 | AGCTATAAATACAAG:AGCTATAAATACAAG | 5 |
| **chr2** | 44540028 | 44540028 | SNV | A | A:T | 1 | | 0 |
| **chr2** | 44540072 | 44540072 | SNV | A | A:G | 1 | | 0 |
| **chr2** | 44540112 | 44540112 | SNV | T | A:T | 4 | | 0 |
| **chr2** | 44540180 | 44540180 | SNV | G | C:C | 1 | C:G | 77 |
| **chr2** | 44540219 | 44540219 | SNV | G | A:A | 2 | A:G | 87 |
| **chr2** | 44540241 | 44540241 | SNV | C | T:T | 1196 | C:T | 930 |
| **chr2** | 44540341 | 44540341 | SNV | T | A:T | 1 | | 0 |
| **chr2** | 44540361 | 44540361 | SNV | A | A:C | 1 | | 0 |
| **chr2** | 44540405 | 44540405 | SNV | G | C:G | 201 | C:C | 15 |
| **chr2** | 44540455 | 44540455 | SNV | C | A:C | 6 | | 0 |
| **chr2** | 44540519 | 44540519 | insertion | - | -:C | 10 | | 0 |
| **chr2** | 44540548 | 44540548 | SNV | G | A:G | 76 | A:A | 4 |
| **chr2** | 44540581 | 44540581 | SNV | C | C:T | 49 | T:T | 1 |
| **chr2** | 44540677 | 44540677 | SNV | C | C:T | 1 | | 0 |
| **chr2** | 44540766 | 44540766 | SNV | A | A:C | 1 | | 0 |
| **chr2** | 44540807 | 44540807 | SNV | A | A:G | 1 | | 0 |
| **chr2** | 44540874 | 44540874 | SNV | G | C:G | 3 | | 0 |
| **chr2** | 44541045 | 44541045 | SNV | C\|N | C:T\|N:N | 9 | | 0 |
| **chr2** | 44541170 | 44541170 | SNV | T | A:T | 1 | | 0 |
| **chr2** | 44541257 | 44541257 | SNV | T | C:T | 18 | | 0 |
| **chr2** | 44541302 | 44541302 | SNV | A | A:G | 9 | | 0 |
| **chr2** | 44541342 | 44541342 | SNV | C | C:G | 144 | G:G | 5 |
| **chr2** | 44541512 | 44541512 | SNV | C | C:T | 86 | T:T | 2 |
| **chr2** | 44541688 | 44541688 | SNV | A | A:G | 56 | G:G | 1 |
| **chr2** | 44541706 | 44541706 | SNV | G | C:G | 873 | C:C | 1276 |
| **chr2** | 44541725 | 44541725 | SNV | C | C:T | 1191 | T:T | 778 |
| **chr2** | 44541804 | 44541804 | SNV | A | A:G | 27 | | 0 |
| **chr2** | 44541844 | 44541844 | SNV | G | G:T | 4 | | 0 |
| **chr2** | 44541857 | 44541857 | SNV | C | G:G | 16 | C:G | 282 |
| **chr2** | 44541896 | 44541896 | SNV | T | C:T | 74 | C:C | 1 |
| **chr2** | 44541918 | 44541919 | deletion | AT | -:- | 783 | -:AT | 1188 |
| **chr2** | 44541997 | 44541997 | SNV | T | G:G | 1 | G:T | 75 |
| **chr2** | 44542039 | 44542039 | SNV | G | A:A | 1 | A:G | 35 |
| **chr2** | 44542062 | 44542073 | deletion | TTTTTTTTTTTT | -:TTTTTTTTTTTT | 1112 | -:- | 840 |
| **chr2** | 44542147 | 44542147 | SNV | C | C:T | 172 | T:T | 13 |
| **chr2** | 44542161 | 44542161 | SNV | C | C:G | 104 | G:G | 1 |
| **chr2** | 44542219 | 44542219 | SNV | C | T:T | 2 | C:T | 109 |
| **chr2** | 44542284 | 44542284 | SNV | G | A:G | 29 | | 0 |
| **chr2** | 44542640 | 44542640 | SNV | G | A:A | 5 | A:G | 144 |

**Supplementary Table 1. Variants of SLC3A1 in the 1KG database**

| Chr | Start Position | End Position | Variant type | Ref Genotype \| Ref Proteotype | Variant Genotype 1\| Variant Proteotype 1 | Count | Variant Genotype 2\| Variant Proteotype 2 | Count |
|---|---|---|---|---|---|---|---|---|
| **chr2** | 44542643 | 44542643 | deletion | A | -:A | 961 | -:- | 1212 |
| **chr2** | 44542712 | 44542712 | SNV | C | C:T | 1 | | 0 |
| **chr2** | 44542734 | 44542734 | SNV | G | C:G | 9 | | 0 |
| **chr2** | 44542737 | 44542737 | SNV | G | C:G | 19 | | 0 |
| **chr2** | 44542817 | 44542817 | SNV | A | A:G | 1 | | 0 |
| **chr2** | 44542863 | 44542863 | SNV | T | G:T | 35 | | 0 |
| **chr2** | 44542951 | 44542953 | deletion | TGG | -:- | 1 | -:TGG | 63 |
| **chr2** | 44543023 | 44543023 | SNV | G | C:G | 221 | C:C | 16 |
| **chr2** | 44543097 | 44543097 | SNV | C | A:C | 34 | | 0 |
| **chr2** | 44543125 | 44543125 | SNV | C | C:G | 859 | G:G | 1290 |
| **chr2** | 44543243 | 44543243 | SNV | G | A:G | 70 | A:A | 1 |
| **chr2** | 44543303 | 44543303 | SNV | C | C:G | 93 | G:G | 5 |
| **chr2** | 44543317 | 44543317 | SNV | C | C:T | 886 | T:T | 1256 |
| **chr2** | 44543389 | 44543389 | SNV | T | A:T | 1090 | A:A | 593 |
| **chr2** | 44543434 | 44543434 | SNV | G | C:G | 22 | | 0 |
| **chr2** | 44543595 | 44543598 | deletion | AAAG | -:- | 5 | -:AAAG | 144 |
| **chr2** | 44543671 | 44543671 | SNV | T | C:T | 5 | | 0 |
| **chr2** | 44544024 | 44544024 | SNV | A | A:G | 27 | | 0 |
| **chr2** | 44544115 | 44544115 | SNV | T | A:A | 15 | A:T | 225 |
| **chr2** | 44544174 | 44544174 | SNV | G | A:A | 5 | A:G | 145 |
| **chr2** | 44544230 | 44544230 | SNV | A | A:G | 34 | | 0 |
| **chr2** | 44544264 | 44544264 | SNV | C | C:G | 145 | G:G | 5 |
| **chr2** | 44544340 | 44544340 | SNV | G | A:A | 1 | A:G | 70 |
| **chr2** | 44544478 | 44544478 | SNV | G | T:T | 1 | G:T | 106 |
| **chr2** | 44544511 | 44544511 | SNV | G | A:G | 1 | | 0 |
| **chr2** | 44544592 | 44544592 | SNV | T | C:T | 9 | | 0 |
| **chr2** | 44544598 | 44544598 | insertion | - | -:AA | 1212 | AA:AA | 818 |
| **chr2** | 44544785 | 44544785 | SNV | T | A:T | 38 | A:A | 1 |
| **chr2** | 44544857 | 44544857 | SNV | G | C:G | 6 | | 0 |
| **chr2** | 44544901 | 44544901 | SNV | T | G:T | 2 | | 0 |
| **chr2** | 44544914 | 44544914 | SNV | A | A:C | 276 | C:C | 13 |
| **chr2** | 44544936 | 44544936 | SNV | C | C:T | 17 | | 0 |
| **chr2** | 44544949 | 44544949 | SNV | G | A:A | 1 | A:G | 51 |
| **chr2** | 44544955 | 44544955 | SNV | G | A:G | 1190 | A:A | 800 |
| **chr2** | 44545036 | 44545036 | SNV | A | A:G | 6 | | 0 |
| **chr2** | 44545076 | 44545076 | SNV | T | C:C | 1 | C:T | 26 |
| **chr2** | 44545099 | 44545101 | deletion | AAT | -:AAT | 105 | -:- | 1 |
| **chr2** | 44545112 | 44545112 | SNV | T | G:G | 1 | G:T | 68 |
| **chr2** | 44545220 | 44545220 | SNV | A | A:C | 1 | | 0 |
| **chr2** | 44545234 | 44545234 | SNV | G | T:T | 24 | G:T | 216 |
| **chr2** | 44545235 | 44545235 | deletion | T | -:- | 56 | -:T | 1624 |
| **chr2** | 44545298 | 44545298 | SNV | C | T:T | 16 | C:T | 217 |
| **chr2** | 44545379 | 44545379 | SNV | G | A:G | 4 | | 0 |
| **chr2** | 44545391 | 44545391 | SNV | G | A:G | 47 | | 0 |
| **chr2** | 44545451 | 44545451 | SNV | G | C:G | 33 | | 0 |
| **chr2** | 44545483 | 44545483 | deletion | G | -:G | 21 | | 0 |
| **chr2** | 44545484 | 44545484 | SNV | C | A:C | 21 | | 0 |
| **chr2** | 44545506 | 44545506 | SNV | G | C:G | 1 | | 0 |
| **chr2** | 44545545 | 44545545 | SNV | T | G:T | 87 | G:G | 6 |
| **chr2** | 44545576 | 44545576 | SNV | T | C:C | 899 | C:T | 1150 |
| **chr2** | 44545705 | 44545705 | SNV | C | C:G | 63 | G:G | 1 |
| **chr2** | 44545712 | 44545712 | SNV | T | G:T | 23 | G:G | 1 |
| **chr2** | 44545737 | 44545737 | insertion | - | TGTT:TGTT | 1233 | -:TGTT | 908 |
| **chr2** | 44545740 | 44545740 | SNV | A | C:C | 1233 | A:C | 908 |
| **chr2** | 44545741 | 44545741 | SNV | A | T:T | 1159 | A:T | 824 |
| **chr2** | 44545743 | 44545743 | insertion | - | A:A | 1233 | -:A | 908 |
| **chr2** | 44545745 | 44545745 | SNV | A | G:G | 1233 | A:G | 908 |
| **chr2** | 44545746 | 44545746 | SNV | T | C:C | 1233 | C:T | 908 |
| **chr2** | 44545750 | 44545750 | SNV | T | G:T | 908 | G:G | 1233 |
| **chr2** | 44545752 | 44545752 | SNV | C | A:C | 908 | A:A | 1233 |
| **chr2** | 44545753 | 44545753 | SNV | T | A:A | 1151 | A:T | 831 |
| **chr2** | 44545755 | 44545758 | deletion | TGCC | -:- | 1233 | -:TGCC | 908 |
| **chr2** | 44545762 | 44545762 | SNV | G | C:G | 910 | C:C | 1231 |
| **chr2** | 44545763 | 44545763 | SNV | C | A:C | 910 | A:A | 1231 |
| **chr2** | 44545764 | 44545764 | SNV | A | T:T | 1230 | A:T | 909 |
| **chr2** | 44545765 | 44545765 | SNV | A | T:T | 1230 | A:T | 909 |
| **chr2** | 44545766 | 44545766 | SNV | A | T:T | 1230 | A:T | 909 |
| **chr2** | 44545776 | 44545776 | SNV | C | A:A | 1 | A:C | 67 |
| **chr2** | 44545936 | 44545936 | SNV | T | C:T | 1 | | 0 |
| **chr2** | 44546016 | 44546016 | SNV | C | T:T | 1243 | C:T | 892 |
| **chr2** | 44546176 | 44546177 | deletion | AT | -:AT | 34 | | 0 |
| **chr2** | 44546177 | 44546177 | SNV | T | C:T | 2 | | 0 |
| **chr2** | 44546369 | 44546369 | SNV | C | C:G | 14 | | 0 |
| **chr2** | 44546403 | 44546403 | SNV | T | A:T | 32 | | 0 |
| **chr2** | 44546567 | 44546567 | SNV | T | C:T | 3 | | 0 |
| **chr2** | 44546601 | 44546601 | SNV | C | A:A | 1 | A:C | 45 |
| **chr2** | 44546633 | 44546636 | deletion | ATTT | -:- | 8 | -:ATTT | 233 |
| **chr2** | 44546661 | 44546661 | SNV | T | C:T | 7 | | 0 |
| **chr2** | 44546741 | 44546741 | SNV | T | C:C | 640 | C:T | 1056 |
| **chr2** | 44547009 | 44547009 | SNV | T | C:T | 5 | | 0 |
| **chr2** | 44547131 | 44547131 | SNV | A | G:G | 920 | A:G | 1138 |
| **chr2** | 44547142 | 44547142 | SNV | A | G:G | 17 | A:G | 218 |
| **chr2** | 44547236 | 44547236 | SNV | G | C:C | 22 | C:G | 209 |
| **chr2** | 44547353 | 44547353 | SNV | C\|P | C:T\|P:S | 1 | | 0 |
| **chr2** | 44547574 | 44547574 | SNV | G\|M | A:G\|I:M | 1048 | A:A\|I:I | 630 |
| **chr2** | 44547821 | 44547821 | SNV | T | A:T | 3 | | 0 |
| **chr2** | 44547822 | 44547822 | SNV | T | C:T | 3 | | 0 |
| **chr2** | 44547899 | 44547899 | SNV | T | C:T | 3 | | 0 |
| **chr2** | 44547909 | 44547909 | SNV | T | C:T | 1166 | C:C | 858 |
| **chr2** | 44526955 | 44526955 | SNV | T | C:T | 9 | | 0 |
| **chr2** | 44508472 | 44508472 | SNV | A | A:G | 1 | | 0 |
| **chr2** | 44511654 | 44511654 | SNV | C | C:T | 2 | | 0 |
| **chr2** | 44546206 | 44546206 | SNV | G | A:G | 2 | | 0 |
| **chr2** | 44509571 | 44509571 | SNV | G | A:G | 27 | | 0 |

**Supplementary Table 1. Variants of SLC3A1 in the 1KG database**

| Chr | Start Position | End Position | Variant type | Ref Genotype \| Ref Proteotype | Variant Genotype 1\| Variant Proteotype 1 | Count | Variant Genotype 2\| Variant Proteotype 2 | Count |
|---|---|---|---|---|---|---|---|---|
| **chr2** | 44527402 | 44527402 | SNV | G | C:G | 6 | | 0 |
| **chr2** | 44517721 | 44517721 | SNV | C | C:T | 8 | | 0 |
| **chr2** | 44503323 | 44503323 | deletion | G | -:G | 4 | | 0 |
| **chr2** | 44536288 | 44536288 | SNV | A | A:G | 1 | | 0 |
| **chr2** | 44533738 | 44533738 | SNV | G | A:G | 1 | | 0 |
| **chr2** | 44515372 | 44515372 | SNV | C | C:T | 2 | | 0 |
| **chr2** | 44508707 | 44508707 | SNV | A | A:G | 2 | | 0 |
| **chr2** | 44513953 | 44513953 | SNV | C | C:T | 3 | | 0 |
| **chr2** | 44533181 | 44533181 | SNV | C | C:T | 1 | | 0 |
| **chr2** | 44530924 | 44530924 | SNV | G | A:G | 3 | | 0 |
| **chr2** | 44527452 | 44527452 | SNV | C | C:T | 4 | | 0 |
| **chr2** | 44530190 | 44530190 | SNV | A | A:T | 1 | | 0 |
| **chr2** | 44547095 | 44547095 | SNV | A | A:G | 1 | | 0 |
| **chr2** | 44504817 | 44504817 | SNV | C | C:T | 1 | | 0 |
| **chr2** | 44539829 | 44539829 | SNV | C\|Y | C:T\|Y:Y | 1 | | 0 |
| **chr2** | 44515978 | 44515978 | SNV | T | A:T | 1 | | 0 |
| **chr2** | 44540832 | 44540832 | SNV | G | G:T | 1 | | 0 |
| **chr2** | 44524203 | 44524203 | SNV | C | C:G | 1 | | 0 |
| **chr2** | 44540737 | 44540737 | SNV | T | C:T | 4 | | 0 |
| **chr2** | 44532243 | 44532243 | SNV | G | A:G | 2 | | 0 |
| **chr2** | 44536150 | 44536150 | SNV | C | C:T | 12 | | 0 |
| **chr2** | 44520679 | 44520679 | SNV | A | A:T | 1 | | 0 |
| **chr2** | 44512100 | 44512100 | SNV | C | C:G | 1 | | 0 |
| **chr2** | 44508265 | 44508274 | deletion | TCCCCTCCCC | -:TCCCCTCCCC | 7 | | 0 |
| **chr2** | 44508966 | 44508971 | deletion | TCCATG | -:- | 1 | -:TCCATG | 62 |
| **chr2** | 44544316 | 44544316 | SNV | T | C:T | 2 | | 0 |
| **chr2** | 44505869 | 44505869 | SNV | T | C:T | 1 | | 0 |
| **chr2** | 44523495 | 44523495 | SNV | G | A:G | 3 | | 0 |
| **chr2** | 44545266 | 44545266 | SNV | G | A:G | 1 | | 0 |
| **chr2** | 44520876 | 44520876 | SNV | A | A:G | 1 | | 0 |
| **chr2** | 44527392 | 44527392 | SNV | A | A:G | 1 | | 0 |
| **chr2** | 44524090 | 44524090 | SNV | G | A:G | 12 | | 0 |
| **chr2** | 44513722 | 44513722 | SNV | G | A:G | 3 | | 0 |
| **chr2** | 44530956 | 44530956 | SNV | G | A:G | 2 | | 0 |
| **chr2** | 44524409 | 44524409 | SNV | C | C:G | 6 | | 0 |
| **chr2** | 44545528 | 44545528 | SNV | C | A:C | 1 | | 0 |
| **chr2** | 44504008 | 44504008 | SNV | G | A:G | 2 | | 0 |
| **chr2** | 44526381 | 44526381 | SNV | T | C:T | 1 | | 0 |
| **chr2** | 44515564 | 44515564 | SNV | C | A:C | 6 | | 0 |
| **chr2** | 44529858 | 44529858 | SNV | C | C:G | 1 | | 0 |
| **chr2** | 44538644 | 44538644 | insertion | - | -:A | 8 | | 0 |
| **chr2** | 44505903 | 44505903 | SNV | G | A:G | 13 | | 0 |
| **chr2** | 44512995 | 44512995 | SNV | T | C:T | 6 | | 0 |
| **chr2** | 44538007 | 44538007 | SNV | G | A:G | 2 | | 0 |
| **chr2** | 44511129 | 44511129 | SNV | G | A:G | 1 | | 0 |
| **chr2** | 44542714 | 44542714 | SNV | G | A:A | 1 | A:G | 1 |
| **chr2** | 44524308 | 44524308 | SNV | C | C:T | 7 | | 0 |
| **chr2** | 44509489 | 44509489 | SNV | A | A:T | 1 | | 0 |
| **chr2** | 44512288 | 44512288 | SNV | C | C:T | 1 | | 0 |
| **chr2** | 44502905 | 44502905 | SNV | T\|S | A:T\|S:S | 3 | | 0 |
| **chr2** | 44536689 | 44536689 | SNV | A | A:T | 1 | | 0 |
| **chr2** | 44535110 | 44535110 | SNV | G | C:G | 1 | | 0 |
| **chr2** | 44524981 | 44524981 | SNV | C | C:G | 8 | | 0 |
| **chr2** | 44508463 | 44508463 | SNV | G | A:G | 4 | | 0 |
| **chr2** | 44542939 | 44542939 | SNV | T | C:T | 1 | | 0 |
| **chr2** | 44544041 | 44544041 | SNV | T | C:T | 2 | | 0 |
| **chr2** | 44525315 | 44525315 | SNV | C | C:T | 2 | | 0 |
| **chr2** | 44522753 | 44522753 | SNV | C | C:T | 6 | | 0 |
| **chr2** | 44506946 | 44506946 | SNV | C | A:C | 1 | | 0 |
| **chr2** | 44533595 | 44533595 | SNV | C | C:G | 7 | | 0 |
| **chr2** | 44544067 | 44544067 | SNV | A | A:C | 2 | | 0 |
| **chr2** | 44530201 | 44530201 | SNV | G | A:G | 2 | | 0 |
| **chr2** | 44503721 | 44503721 | SNV | C | C:G | 1 | | 0 |
| **chr2** | 44519313 | 44519313 | SNV | C | C:G | 13 | | 0 |
| **chr2** | 44542995 | 44542995 | SNV | C | C:T | 1 | | 0 |
| **chr2** | 44519944 | 44519944 | SNV | G | A:G | 5 | | 0 |
| **chr2** | 44544273 | 44544273 | SNV | T | C:T | 1 | | 0 |
| **chr2** | 44534104 | 44534104 | SNV | A | A:G | 14 | | 0 |
| **chr2** | 44521437 | 44521437 | SNV | A | A:T | 1 | | 0 |
| **chr2** | 44525022 | 44525022 | SNV | C | A:C | 2 | | 0 |
| **chr2** | 44526328 | 44526328 | SNV | C | C:G | 2 | | 0 |
| **chr2** | 44513328 | 44513328 | SNV | T | C:T | 1 | | 0 |
| **chr2** | 44507705 | 44507705 | SNV | C | C:T | 1 | | 0 |
| **chr2** | 44541205 | 44541205 | SNV | T | C:T | 1 | | 0 |
| **chr2** | 44529568 | 44529568 | SNV | T | C:T | 2 | | 0 |
| **chr2** | 44513366 | 44513366 | SNV | T | C:T | 1 | | 0 |
| **chr2** | 44534606 | 44534606 | SNV | C | C:G | 1 | | 0 |
| **chr2** | 44537458 | 44537458 | SNV | T | C:T | 1 | | 0 |
| **chr2** | 44514746 | 44514746 | SNV | T | C:T | 2 | | 0 |
| **chr2** | 44537363 | 44537363 | SNV | G | G:T | 2 | | 0 |
| **chr2** | 44527090 | 44527090 | deletion | T | -:T | 3 | | 0 |
| **chr2** | 44545114 | 44545114 | SNV | G | A:G | 1 | | 0 |
| **chr2** | 44542648 | 44542648 | SNV | A | A:G | 9 | G:G | 1 |
| **chr2** | 44516960 | 44516960 | SNV | G | A:G | 1 | | 0 |
| **chr2** | 44506014 | 44506014 | SNV | C | A:C | 1 | | 0 |
| **chr2** | 44523270 | 44523270 | SNV | C | A:C | 1 | | 0 |
| **chr2** | 44502974 | 44502974 | SNV | C\|L | C:T\|L:L | 8 | | 0 |
| **chr2** | 44538824 | 44538824 | SNV | C | C:T | 5 | | 0 |
| **chr2** | 44546476 | 44546476 | SNV | T | C:T | 4 | | 0 |
| **chr2** | 44531926 | 44531926 | SNV | C | C:T | 1 | | 0 |
| **chr2** | 44546123 | 44546123 | SNV | G | C:G | 1 | | 0 |
| **chr2** | 44535712 | 44535712 | SNV | T | C:T | 2 | | 0 |
| **chr2** | 44538973 | 44538973 | SNV | T | C:T | 2 | | 0 |
| **chr2** | 44524323 | 44524323 | SNV | T | C:T | 3 | | 0 |

**Supplementary Table 1. Variants of SLC3A1 in the 1KG database**

| Chr | Start Position | End Position | Variant type | Ref Genotype \| Ref Proteotype | Variant Genotype 1\| Variant Proteotype 1 | Count | Variant Genotype 2\| Variant Proteotype 2 | Count |
|---|---|---|---|---|---|---|---|---|
| **chr2** | 44545626 | 44545626 | SNV | T | C:T | 10 | | 0 |
| **chr2** | 44539726 | 44539726 | SNV | T\|I | C:T\|T:I | 2 | | 0 |
| **chr2** | 44505535 | 44505535 | SNV | T | G:T | 8 | | 0 |
| **chr2** | 44518869 | 44518869 | SNV | G | A:G | 3 | | 0 |
| **chr2** | 44507044 | 44507044 | SNV | G | C:G | 2 | | 0 |
| **chr2** | 44541576 | 44541576 | SNV | C | C:T | 1 | | 0 |
| **chr2** | 44518670 | 44518670 | SNV | T | G:T | 1 | | 0 |
| **chr2** | 44522011 | 44522011 | insertion | - | -:T | 3 | | 0 |
| **chr2** | 44511436 | 44511436 | SNV | C | A:C | 1 | | 0 |
| **chr2** | 44527072 | 44527072 | SNV | A | A:G | 2 | | 0 |
| **chr2** | 44504400 | 44504400 | SNV | T | C:T | 1 | | 0 |
| **chr2** | 44537295 | 44537295 | SNV | G | A:G | 9 | | 0 |
| **chr2** | 44547261 | 44547261 | deletion | C | -:C | 5 | | 0 |
| **chr2** | 44522426 | 44522426 | SNV | C | C:T | 1 | | 0 |
| **chr2** | 44508595 | 44508595 | SNV | G\|R | A:G\|Q:R | 1 | | 0 |
| **chr2** | 44532734 | 44532734 | deletion | T | -:T | 3 | | 0 |
| **chr2** | 44510808 | 44510808 | SNV | T | C:T | 5 | | 0 |
| **chr2** | 44538615 | 44538615 | SNV | C | A:C | 1 | | 0 |
| **chr2** | 44516000 | 44516000 | SNV | G | G:T | 7 | | 0 |
| **chr2** | 44541430 | 44541430 | insertion | - | -:AGA | 33 | AGA:AGA | 1 |
| **chr2** | 44542639 | 44542639 | SNV | C | C:T | 14 | | 0 |
| **chr2** | 44507566 | 44507566 | SNV | C | C:T | 9 | | 0 |
| **chr2** | 44517254 | 44517254 | SNV | C | C:T | 19 | | 0 |
| **chr2** | 44526412 | 44526412 | SNV | A | A:T | 10 | | 0 |
| **chr2** | 44508490 | 44508490 | SNV | C | A:C | 1 | | 0 |
| **chr2** | 44537091 | 44537091 | SNV | T | A:T | 2 | | 0 |
| **chr2** | 44514210 | 44514210 | SNV | T | C:T | 8 | | 0 |
| **chr2** | 44546068 | 44546072 | deletion | TCTGC | -:TCTGC | 3 | | 0 |
| **chr2** | 44531449 | 44531449 | SNV | T\|M | G:T\|R:M | 1 | | 0 |
| **chr2** | 44530728 | 44530730 | deletion | TTA | -:TTA | 11 | | 0 |
| **chr2** | 44532870 | 44532870 | SNV | T | G:T | 1 | | 0 |
| **chr2** | 44503780 | 44503780 | SNV | G | C:G | 2 | | 0 |
| **chr2** | 44515261 | 44515261 | SNV | G | C:G | 5 | | 0 |
| **chr2** | 44516237 | 44516237 | SNV | G | A:G | 1 | | 0 |
| **chr2** | 44527226 | 44527226 | SNV | C\|I | A:C\|I:I | 1 | | 0 |
| **chr2** | 44518578 | 44518578 | SNV | T | C:T | 8 | | 0 |
| **chr2** | 44514520 | 44514520 | SNV | A | A:G | 1 | | 0 |
| **chr2** | 44525939 | 44525939 | SNV | C | C:T | 1 | | 0 |
| **chr2** | 44538245 | 44538245 | SNV | T | C:T | 1 | | 0 |
| **chr2** | 44516334 | 44516339 | deletion | TTCTTT | -:TTCTTT | 9 | | 0 |
| **chr2** | 44532012 | 44532012 | SNV | G | C:G | 1 | | 0 |
| **chr2** | 44540432 | 44540432 | SNV | C | C:T | 8 | | 0 |
| **chr2** | 44526905 | 44526905 | SNV | G | C:G | 2 | | 0 |
| **chr2** | 44504687 | 44504687 | SNV | C | C:G | 1 | | 0 |
| **chr2** | 44529813 | 44529813 | SNV | A | A:G | 6 | | 0 |
| **chr2** | 44527672 | 44527672 | SNV | C | C:G | 4 | G:G | 1 |
| **chr2** | 44509196 | 44509196 | SNV | A | A:C | 2 | | 0 |
| **chr2** | 44505408 | 44505408 | SNV | C | A:C | 1 | | 0 |
| **chr2** | 44506749 | 44506749 | SNV | T | G:T | 37 | G:G | 1 |
| **chr2** | 44511463 | 44511463 | SNV | C | C:T | 2 | | 0 |
| **chr2** | 44526895 | 44526895 | SNV | A | A:T | 9 | | 0 |
| **chr2** | 44529758 | 44529758 | SNV | T | C:T | 10 | | 0 |
| **chr2** | 44546343 | 44546343 | SNV | A | A:G | 1 | | 0 |
| **chr2** | 44537166 | 44537166 | SNV | G | C:G | 2 | | 0 |
| **chr2** | 44510060 | 44510060 | SNV | G | C:G | 1 | | 0 |
| **chr2** | 44526493 | 44526493 | SNV | T | C:T | 1 | | 0 |
| **chr2** | 44535316 | 44535316 | SNV | T | A:T | 6 | | 0 |
| **chr2** | 44512977 | 44512977 | SNV | A | A:T | 1 | | 0 |
| **chr2** | 44515987 | 44515987 | SNV | T | A:T | 1 | | 0 |
| **chr2** | 44512934 | 44512934 | SNV | T | C:T | 1 | | 0 |
| **chr2** | 44522619 | 44522619 | SNV | C | C:T | 1 | | 0 |
| **chr2** | 44508972 | 44508972 | SNV | T | A:T | 1 | | 0 |
| **chr2** | 44514701 | 44514701 | SNV | G | G:T | 6 | T:T | 1 |
| **chr2** | 44511471 | 44511471 | SNV | C | C:T | 1 | | 0 |
| **chr2** | 44522400 | 44522400 | SNV | G | A:G | 8 | | 0 |
| **chr2** | 44510176 | 44510176 | SNV | C | C:T | 1 | | 0 |
| **chr2** | 44546421 | 44546421 | SNV | T | G:T | 1 | | 0 |
| **chr2** | 44535607 | 44535607 | SNV | A | A:T | 1 | | 0 |
| **chr2** | 44546619 | 44546619 | SNV | T | C:T | 1 | | 0 |
| **chr2** | 44523707 | 44523707 | SNV | G | A:G | 1 | | 0 |
| **chr2** | 44538858 | 44538858 | SNV | A | A:G | 1 | | 0 |
| **chr2** | 44525308 | 44525308 | SNV | C | C:T | 2 | | 0 |
| **chr2** | 44540480 | 44540480 | SNV | A | A:T | 5 | | 0 |
| **chr2** | 44526437 | 44526437 | SNV | C | C:T | 1 | | 0 |
| **chr2** | 44509807 | 44509807 | SNV | C | C:G | 1 | | 0 |
| **chr2** | 44539142 | 44539142 | SNV | A | A:C | 1 | | 0 |
| **chr2** | 44532467 | 44532467 | SNV | T | C:T | 2 | | 0 |
| **chr2** | 44526987 | 44526987 | SNV | G | A:G | 1 | | 0 |
| **chr2** | 44508443 | 44508443 | SNV | G | A:G | 4 | | 0 |
| **chr2** | 44519389 | 44519389 | SNV | C | C:G | 4 | | 0 |
| **chr2** | 44513193 | 44513193 | SNV | G\|S | C:G\|T:S | 4 | | 0 |
| **chr2** | 44507891 | 44507891 | SNV | A\|N | A:G\|N:S | 1 | | 0 |
| **chr2** | 44504543 | 44504543 | SNV | G | A:G | 11 | | 0 |
| **chr2** | 44530508 | 44530508 | SNV | G | A:G | 2 | | 0 |
| **chr2** | 44538021 | 44538021 | SNV | T | G:T | 1 | | 0 |
| **chr2** | 44538616 | 44538616 | SNV | G | A:G | 5 | | 0 |
| **chr2** | 44514503 | 44514503 | SNV | G | A:G | 3 | | 0 |
| **chr2** | 44517541 | 44517541 | SNV | A | A:G | 1 | | 0 |
| **chr2** | 44532305 | 44532305 | SNV | T | G:T | 2 | | 0 |
| **chr2** | 44525516 | 44525516 | SNV | C | C:T | 3 | | 0 |
| **chr2** | 44505828 | 44505828 | SNV | A | A:C | 1 | | 0 |
| **chr2** | 44547566 | 44547566 | SNV | G\|A | A:G\|T:A | 3 | | 0 |
| **chr2** | 44504442 | 44504442 | SNV | C | C:G | 1 | | 0 |
| **chr2** | 44547233 | 44547233 | SNV | T | C:T | 2 | | 0 |

**Supplementary Table 1. Variants of SLC3A1 in the 1KG database**

| Chr | Start Position | End Position | Variant type | Ref Genotype \| Ref Proteotype | Variant Genotype 1\| Variant Proteotype 1 | Count | Variant Genotype 2\| Variant Proteotype 2 | Count |
|---|---|---|---|---|---|---|---|---|
| **chr2** | 44542312 | 44542312 | SNV | T | C:T | 7 | | 0 |
| **chr2** | 44546773 | 44546773 | SNV | T | A:T | 13 | | 0 |
| **chr2** | 44508225 | 44508225 | SNV | T | C:T | 6 | | 0 |
| **chr2** | 44516632 | 44516632 | SNV | G | A:G | 2 | | 0 |
| **chr2** | 44529086 | 44529086 | SNV | C | C:T | 2 | | 0 |
| **chr2** | 44536522 | 44536522 | SNV | T | C:T | 1 | | 0 |
| **chr2** | 44544212 | 44544212 | SNV | G | C:G | 9 | | 0 |
| **chr2** | 44541148 | 44541148 | SNV | G | C:G | 9 | | 0 |
| **chr2** | 44533323 | 44533323 | SNV | C | C:T | 4 | | 0 |
| **chr2** | 44540146 | 44540146 | SNV | T | C:T | 2 | | 0 |
| **chr2** | 44546290 | 44546290 | SNV | C | A:C | 1 | | 0 |
| **chr2** | 44525469 | 44525470 | deletion | CT | -:CT | 3 | | 0 |
| **chr2** | 44533859 | 44533859 | insertion | - | -:T | 14 | | 0 |
| **chr2** | 44542113 | 44542113 | SNV | C | C:T | 6 | | 0 |
| **chr2** | 44519531 | 44519531 | SNV | C | C:T | 1 | | 0 |
| **chr2** | 44539643 | 44539643 | SNV | G | A:G | 2 | | 0 |
| **chr2** | 44512882 | 44512882 | SNV | T | C:T | 7 | | 0 |
| **chr2** | 44503779 | 44503779 | SNV | C | C:T | 1 | | 0 |
| **chr2** | 44541471 | 44541471 | insertion | - | ATCA:ATCA | 2 | -:ATCA | 45 |
| **chr2** | 44525237 | 44525237 | SNV | G | A:G | 2 | | 0 |
| **chr2** | 44531274 | 44531274 | SNV | T | C:T | 1 | | 0 |
| **chr2** | 44530516 | 44530516 | SNV | G | A:G | 5 | | 0 |
| **chr2** | 44544771 | 44544771 | SNV | G | A:G | 1 | | 0 |
| **chr2** | 44517114 | 44517114 | SNV | G | A:G | 7 | | 0 |
| **chr2** | 44522372 | 44522372 | SNV | G | A:G | 2 | | 0 |
| **chr2** | 44524795 | 44524795 | SNV | A | A:G | 1 | | 0 |
| **chr2** | 44522171 | 44522171 | SNV | C | C:G | 8 | | 0 |
| **chr2** | 44520686 | 44520686 | SNV | G | A:G | 7 | | 0 |
| **chr2** | 44505184 | 44505184 | SNV | T | A:T | 1 | | 0 |
| **chr2** | 44540609 | 44540609 | SNV | T | C:T | 10 | | 0 |
| **chr2** | 44546390 | 44546390 | SNV | G | C:G | 5 | | 0 |
| **chr2** | 44539500 | 44539500 | SNV | T | C:T | 1 | | 0 |
| **chr2** | 44537411 | 44537411 | SNV | T | A:T | 1 | | 0 |
| **chr2** | 44514441 | 44514441 | SNV | T | C:T | 1 | | 0 |
| **chr2** | 44537869 | 44537869 | SNV | C | A:C | 3 | | 0 |
| **chr2** | 44517203 | 44517203 | SNV | A | A:G | 1 | | 0 |
| **chr2** | 44547162 | 44547163 | deletion | TA | -:TA | 3 | | 0 |
| **chr2** | 44527138 | 44527138 | SNV | G\|G | G:T\|G:V | 1 | | 0 |
| **chr2** | 44529820 | 44529820 | SNV | G | A:G | 1 | | 0 |
| **chr2** | 44527701 | 44527701 | SNV | C | C:T | 3 | | 0 |
| **chr2** | 44546793 | 44546793 | SNV | C | C:T | 2 | | 0 |
| **chr2** | 44504591 | 44504591 | SNV | A | A:G | 8 | | 0 |
| **chr2** | 44520483 | 44520483 | SNV | C | C:T | 1 | | 0 |
| **chr2** | 44543267 | 44543267 | SNV | C | A:C | 1 | | 0 |
| **chr2** | 44519344 | 44519344 | SNV | A | A:C | 16 | | 0 |
| **chr2** | 44502927 | 44502927 | SNV | C\|P | C:T\|P:S | 1 | | 0 |
| **chr2** | 44546523 | 44546523 | SNV | A | A:C | 1 | | 0 |
| **chr2** | 44513804 | 44513804 | SNV | G | A:G | 2 | | 0 |
| **chr2** | 44541580 | 44541580 | SNV | G | A:G | 1 | | 0 |
| **chr2** | 44507463 | 44507463 | SNV | C | C:T | 3 | | 0 |
| **chr2** | 44510898 | 44510898 | SNV | G | A:G | 3 | | 0 |
| **chr2** | 44505287 | 44505287 | SNV | A | A:G | 2 | | 0 |
| **chr2** | 44520583 | 44520583 | SNV | G | A:G | 4 | | 0 |
| **chr2** | 44523636 | 44523636 | SNV | A | A:G | 4 | | 0 |
| **chr2** | 44534904 | 44534904 | SNV | C | A:C | 1 | | 0 |
| **chr2** | 44535749 | 44535749 | SNV | T | C:T | 1 | | 0 |
| **chr2** | 44529840 | 44529840 | SNV | A | A:G | 8 | | 0 |
| **chr2** | 44546833 | 44546833 | SNV | T | C:T | 41 | C:C | 1 |
| **chr2** | 44506568 | 44506568 | SNV | G | G:T | 1 | | 0 |
| **chr2** | 44512664 | 44512664 | SNV | A | A:G | 8 | | 0 |
| **chr2** | 44535142 | 44535142 | SNV | G | C:G | 1 | | 0 |
| **chr2** | 44520072 | 44520072 | SNV | G | A:G | 1 | | 0 |
| **chr2** | 44537301 | 44537301 | SNV | G | A:G | 1 | | 0 |
| **chr2** | 44546757 | 44546757 | SNV | T | C:T | 1 | | 0 |
| **chr2** | 44516546 | 44516546 | SNV | C | C:T | 9 | | 0 |
| **chr2** | 44529011 | 44529011 | SNV | C | C:T | 7 | | 0 |
| **chr2** | 44529284 | 44529284 | SNV | C | C:T | 5 | | 0 |
| **chr2** | 44544039 | 44544039 | SNV | G | G:T | 2 | | 0 |
| **chr2** | 44532861 | 44532861 | SNV | C | C:T | 1 | | 0 |
| **chr2** | 44541280 | 44541280 | SNV | A | A:G | 4 | | 0 |
| **chr2** | 44533546 | 44533546 | SNV | C | A:C | 2 | | 0 |
| **chr2** | 44542836 | 44542836 | SNV | C | C:T | 2 | | 0 |
| **chr2** | 44524383 | 44524383 | SNV | C | C:T | 2 | | 0 |
| **chr2** | 44518634 | 44518634 | SNV | C | C:G | 2 | | 0 |
| **chr2** | 44546506 | 44546506 | SNV | T | C:T | 30 | | 0 |
| **chr2** | 44513129 | 44513129 | SNV | A | A:G | 4 | | 0 |
| **chr2** | 44530028 | 44530028 | SNV | G | A:G | 14 | | 0 |
| **chr2** | 44506305 | 44506305 | SNV | T | A:T | 1 | | 0 |
| **chr2** | 44526097 | 44526097 | insertion | - | -:T | 3 | | 0 |
| **chr2** | 44536141 | 44536141 | SNV | G | G:T | 1 | | 0 |
| **chr2** | 44542475 | 44542475 | SNV | G | G:T | 3 | | 0 |
| **chr2** | 44520604 | 44520604 | SNV | C | C:T | 1 | | 0 |
| **chr2** | 44522451 | 44522451 | SNV | A | A:T | 7 | | 0 |
| **chr2** | 44538320 | 44538320 | SNV | G | A:G | 1 | | 0 |
| **chr2** | 44542838 | 44542864 | deletion | AAAACATACAGGAGAAAAACAGTTCTA | -:AAAACATACAGGAGAAAAACAGTTCTA | 4 | | 0 |
| **chr2** | 44505673 | 44505673 | SNV | G | A:G | 2 | | 0 |
| **chr2** | 44539792 | 44539792 | SNV | T\|M | C:T\|T:M | 2 | | 0 |
| **chr2** | 44514687 | 44514687 | SNV | G | G:T | 1 | | 0 |
| **chr2** | 44528937 | 44528937 | SNV | T | C:T | 9 | | 0 |
| **chr2** | 44546652 | 44546652 | SNV | T | C:T | 2 | | 0 |
| **chr2** | 44542174 | 44542174 | SNV | G | A:G | 1 | | 0 |
| **chr2** | 44528580 | 44528580 | SNV | T | G:T | 1 | | 0 |
| **chr2** | 44512718 | 44512718 | SNV | T | G:T | 1 | | 0 |
| **chr2** | 44518143 | 44518143 | SNV | C | C:T | 3 | | 0 |

**Supplementary Table 1. Variants of SLC3A1 in the 1KG database**

| Chr | Start Position | End Position | Variant type | Ref Genotype \| Ref Proteotype | Variant Genotype 1\| Variant Proteotype 1 | Count | Variant Genotype 2\| Variant Proteotype 2 | Count |
|---|---|---|---|---|---|---|---|---|
| **chr2** | 44535994 | 44535994 | SNV | C | C:T | 1 | | 0 |
| **chr2** | 44529891 | 44529891 | SNV | C | C:T | 4 | | 0 |
| **chr2** | 44530260 | 44530260 | SNV | C | C:G | 1 | | 0 |
| **chr2** | 44539713 | 44539713 | SNV | T | A:T | 10 | | 0 |
| **chr2** | 44507885 | 44507885 | SNV | C\|A | C:G\|A:G | 1 | | 0 |
| **chr2** | 44531461 | 44531461 | SNV | A\|K | A:T\|K:I | 2 | | 0 |
| **chr2** | 44516768 | 44516768 | SNV | A | G:G | 3 | A:G | 60 |
| **chr2** | 44545727 | 44545727 | SNV | A | A:G | 11 | | 0 |
| **chr2** | 44507991 | 44507991 | SNV | G\|T | A:G\|T:T | 1 | | 0 |
| **chr2** | 44529424 | 44529424 | SNV | A | A:G | 22 | | 0 |
| **chr2** | 44529576 | 44529576 | SNV | C | C:T | 1 | | 0 |
| **chr2** | 44514659 | 44514659 | SNV | A | A:C | 1 | | 0 |
| **chr2** | 44530604 | 44530604 | SNV | G | G:T | 1 | | 0 |
| **chr2** | 44540185 | 44540185 | SNV | A | A:C | 1 | | 0 |
| **chr2** | 44512267 | 44512267 | SNV | T | G:T | 7 | | 0 |
| **chr2** | 44502601 | 44502601 | SNV | C | C:G | 1 | | 0 |
| **chr2** | 44525166 | 44525166 | SNV | T | A:T | 1 | | 0 |
| **chr2** | 44539097 | 44539097 | SNV | G | A:G | 1 | | 0 |
| **chr2** | 44523017 | 44523018 | deletion | TT | -:TT | 4 | | 0 |
| **chr2** | 44525323 | 44525323 | SNV | C | G:G | 2 | C:G | 44 |
| **chr2** | 44506648 | 44506648 | SNV | C | C:G | 3 | | 0 |
| **chr2** | 44542044 | 44542044 | SNV | T | C:T | 14 | | 0 |
| **chr2** | 44518800 | 44518800 | SNV | G | G:T | 1 | | 0 |
| **chr2** | 44530628 | 44530628 | SNV | G | G:T | 2 | | 0 |
| **chr2** | 44535483 | 44535483 | SNV | T | A:T | 1 | | 0 |
| **chr2** | 44507793 | 44507793 | insertion | - | -:T | 39 | | 0 |
| **chr2** | 44533059 | 44533059 | SNV | A | A:G | 2 | | 0 |
| **chr2** | 44519614 | 44519614 | SNV | G | G:T | 8 | | 0 |
| **chr2** | 44524053 | 44524053 | SNV | C | C:T | 1 | | 0 |
| **chr2** | 44541621 | 44541621 | SNV | A | A:G | 2 | | 0 |
| **chr2** | 44532866 | 44532866 | SNV | C | A:C | 1 | | 0 |
| **chr2** | 44539321 | 44539321 | SNV | A | A:G | 6 | | 0 |
| **chr2** | 44533043 | 44533043 | SNV | C | C:G | 1 | | 0 |
| **chr2** | 44531863 | 44531863 | SNV | G | C:G | 1 | | 0 |
| **chr2** | 44546145 | 44546145 | SNV | C | C:T | 1 | | 0 |
| **chr2** | 44511257 | 44511257 | SNV | C | C:T | 9 | | 0 |
| **chr2** | 44507809 | 44507809 | SNV | C | A:C | 6 | | 0 |
| **chr2** | 44530939 | 44530939 | SNV | A | A:T | 1 | | 0 |
| **chr2** | 44506624 | 44506624 | SNV | G | A:G | 13 | | 0 |
| **chr2** | 44533583 | 44533583 | SNV | C | C:T | 2 | | 0 |
| **chr2** | 44523928 | 44523928 | SNV | C | C:T | 23 | | 0 |
| **chr2** | 44545505 | 44545505 | SNV | C | C:T | 4 | | 0 |
| **chr2** | 44522859 | 44522859 | SNV | C | C:T | 1 | | 0 |
| **chr2** | 44504769 | 44504769 | SNV | A | A:T | 6 | | 0 |
| **chr2** | 44536852 | 44536852 | SNV | C | C:G | 1 | | 0 |
| **chr2** | 44524280 | 44524280 | SNV | G | C:G | 1 | | 0 |
| **chr2** | 44544667 | 44544667 | SNV | G | A:G | 1 | | 0 |
| **chr2** | 44538635 | 44538635 | SNV | A | A:G | 2 | | 0 |
| **chr2** | 44507473 | 44507473 | SNV | G | C:G | 3 | | 0 |
| **chr2** | 44530366 | 44530366 | SNV | A | A:G | 1 | | 0 |
| **chr2** | 44542334 | 44542334 | SNV | G | G:T | 1 | | 0 |
| **chr2** | 44504743 | 44504743 | SNV | C | C:T | 3 | | 0 |
| **chr2** | 44514217 | 44514217 | SNV | C | C:T | 1 | | 0 |
| **chr2** | 44508100 | 44508100 | SNV | A | A:G | 1 | | 0 |
| **chr2** | 44517016 | 44517016 | SNV | C | C:T | 12 | | 0 |
| **chr2** | 44511802 | 44511802 | SNV | A | A:G | 1 | | 0 |
| **chr2** | 44535384 | 44535384 | SNV | T | G:T | 1 | | 0 |
| **chr2** | 44513735 | 44513735 | SNV | G | C:G | 9 | | 0 |
| **chr2** | 44533561 | 44533561 | SNV | G | A:G | 2 | | 0 |
| **chr2** | 44518185 | 44518185 | SNV | A | A:C | 1 | | 0 |
| **chr2** | 44513734 | 44513734 | SNV | G | A:G | 9 | | 0 |
| **chr2** | 44524052 | 44524052 | SNV | G | G:T | 1 | | 0 |
| **chr2** | 44524642 | 44524642 | SNV | C | C:T | 1 | | 0 |
| **chr2** | 44507417 | 44507417 | SNV | G | A:G | 3 | | 0 |
| **chr2** | 44536164 | 44536164 | SNV | C | C:G | 5 | | 0 |
| **chr2** | 44522741 | 44522741 | SNV | A | A:C | 8 | | 0 |
| **chr2** | 44545092 | 44545092 | SNV | G | C:G | 2 | | 0 |
| **chr2** | 44533197 | 44533197 | SNV | C | C:T | 1 | | 0 |
| **chr2** | 44527547 | 44527547 | SNV | C | C:T | 1 | | 0 |
| **chr2** | 44526544 | 44526544 | SNV | G | A:G | 33 | A:A | 1 |
| **chr2** | 44520540 | 44520540 | SNV | C | C:T | 1 | | 0 |
| **chr2** | 44508364 | 44508364 | SNV | C | C:T | 2 | | 0 |
| **chr2** | 44512084 | 44512084 | SNV | C | A:C | 7 | | 0 |
| **chr2** | 44536860 | 44536860 | SNV | C | C:G | 1 | | 0 |
| **chr2** | 44546476 | 44546477 | deletion | TT | -:TT | 6 | | 0 |
| **chr2** | 44515750 | 44515750 | SNV | T | A:T | 13 | | 0 |
| **chr2** | 44529906 | 44529906 | SNV | A | A:G | 1 | | 0 |
| **chr2** | 44531907 | 44531907 | SNV | C | C:T | 34 | T:T | 1 |
| **chr2** | 44545495 | 44545495 | SNV | G | C:G | 4 | | 0 |
| **chr2** | 44511942 | 44511942 | SNV | C | C:G | 2 | | 0 |
| **chr2** | 44511336 | 44511336 | SNV | A | A:G | 3 | | 0 |
| **chr2** | 44527409 | 44527409 | SNV | T | C:T | 1 | | 0 |
| **chr2** | 44508488 | 44508488 | SNV | A | A:T | 7 | | 0 |
| **chr2** | 44519665 | 44519665 | SNV | G | C:G | 6 | | 0 |
| **chr2** | 44547140 | 44547140 | SNV | G | A:G | 10 | | 0 |
| **chr2** | 44539703 | 44539703 | SNV | C | C:G | 3 | | 0 |
| **chr2** | 44524114 | 44524114 | insertion | - | -:C | 14 | | 0 |
| **chr2** | 44526515 | 44526515 | SNV | C | C:T | 2 | | 0 |
| **chr2** | 44510407 | 44510407 | SNV | G | A:G | 14 | | 0 |
| **chr2** | 44531415 | 44531415 | SNV | G\|V | C:G\|L:V | 1 | | 0 |
| **chr2** | 44524791 | 44524791 | SNV | C | C:T | 2 | | 0 |
| **chr2** | 44537800 | 44537800 | SNV | T | A:T | 1 | | 0 |
| **chr2** | 44509883 | 44509883 | SNV | G | A:G | 8 | | 0 |
| **chr2** | 44525398 | 44525398 | SNV | C | C:G | 1 | | 0 |

**Supplementary Table 1. Variants of SLC3A1 in the 1KG database**

| Chr | Start Position | End Position | Variant type | Ref Genotype \| Ref Proteotype | Variant Genotype 1\| Variant Proteotype 1 | Count | Variant Genotype 2\| Variant Proteotype 2 | Count |
|---|---|---|---|---|---|---|---|---|
| **chr2** | 44509324 | 44509324 | SNV | T | A:T | 1 | | 0 |
| **chr2** | 44547386 | 44547386 | SNV | A\|S | A:G\|S:G | 1 | | 0 |
| **chr2** | 44510932 | 44510932 | SNV | A | A:G | 1 | | 0 |
| **chr2** | 44536208 | 44536208 | SNV | A | A:G | 9 | | 0 |
| **chr2** | 44505721 | 44505721 | SNV | C | C:T | 13 | | 0 |
| **chr2** | 44541704 | 44541704 | SNV | T | G:T | 1 | | 0 |
| **chr2** | 44539044 | 44539044 | SNV | A | A:T | 1 | | 0 |
| **chr2** | 44531597 | 44531597 | SNV | T | C:T | 1 | | 0 |
| **chr2** | 44507416 | 44507416 | SNV | C | C:T | 1 | | 0 |
| **chr2** | 44523759 | 44523759 | SNV | A | A:G | 3 | | 0 |
| **chr2** | 44524853 | 44524853 | SNV | G | A:G | 2 | | 0 |
| **chr2** | 44517351 | 44517351 | SNV | C | A:C | 4 | | 0 |
| **chr2** | 44529574 | 44529574 | SNV | C | C:G | 1 | | 0 |
| **chr2** | 44510158 | 44510158 | SNV | A | A:G | 1 | | 0 |
| **chr2** | 44526327 | 44526327 | SNV | G | A:G | 8 | | 0 |
| **chr2** | 44506778 | 44506780 | deletion | AAG | -:AAG | 5 | | 0 |
| **chr2** | 44546622 | 44546622 | SNV | T | G:T | 2 | | 0 |
| **chr2** | 44545126 | 44545126 | SNV | T | C:T | 12 | | 0 |
| **chr2** | 44504766 | 44504766 | SNV | A | A:G | 1 | | 0 |
| **chr2** | 44542605 | 44542605 | SNV | T | G:T | 11 | | 0 |
| **chr2** | 44527097 | 44527097 | SNV | T | G:T | 1 | | 0 |
| **chr2** | 44517857 | 44517857 | SNV | C | C:T | 2 | | 0 |
| **chr2** | 44515290 | 44515290 | SNV | C | C:T | 3 | | 0 |
| **chr2** | 44528119 | 44528119 | SNV | C | C:T | 12 | | 0 |
| **chr2** | 44537775 | 44537775 | SNV | T | C:T | 1 | | 0 |
| **chr2** | 44527765 | 44527765 | SNV | G | A:G | 2 | | 0 |
| **chr2** | 44520664 | 44520664 | SNV | A | A:G | 14 | | 0 |
| **chr2** | 44525956 | 44525956 | SNV | C | C:G | 3 | | 0 |
| **chr2** | 44514511 | 44514511 | SNV | A | A:G | 1 | | 0 |
| **chr2** | 44545367 | 44545367 | SNV | T | C:T | 6 | | 0 |
| **chr2** | 44506727 | 44506727 | SNV | G | A:G | 1 | | 0 |
| **chr2** | 44527006 | 44527006 | SNV | A | A:G | 2 | | 0 |
| **chr2** | 44544789 | 44544791 | deletion | AAT | -:AAT | 7 | | 0 |
| **chr2** | 44519294 | 44519294 | SNV | T | C:T | 2 | | 0 |
| **chr2** | 44504489 | 44504489 | insertion | - | -:C | 8 | | 0 |
| **chr2** | 44525292 | 44525292 | SNV | T | A:T | 1 | | 0 |
| **chr2** | 44502696 | 44502696 | SNV | A\|R | A:G\|R:G | 1 | | 0 |
| **chr2** | 44536995 | 44536995 | SNV | C | C:T | 1 | | 0 |
| **chr2** | 44539773 | 44539773 | SNV | T\|Y | C:T\|H:Y | 4 | | 0 |
| **chr2** | 44544925 | 44544925 | SNV | T | C:T | 1 | | 0 |
| **chr2** | 44523050 | 44523050 | SNV | A | A:C | 2 | | 0 |
| **chr2** | 44525085 | 44525085 | SNV | T | C:T | 3 | | 0 |
| **chr2** | 44524897 | 44524897 | SNV | C | C:T | 8 | | 0 |
| **chr2** | 44541037 | 44541037 | SNV | G\|A | G:T\|A:S | 1 | | 0 |
| **chr2** | 44507412 | 44507412 | SNV | C | C:G | 1 | | 0 |
| **chr2** | 44517437 | 44517437 | SNV | C | C:T | 1 | | 0 |
| **chr2** | 44528058 | 44528058 | SNV | A | A:G | 3 | | 0 |
| **chr2** | 44535992 | 44535992 | SNV | C | C:T | 1 | | 0 |
| **chr2** | 44522098 | 44522098 | SNV | G | G:T | 9 | | 0 |
| **chr2** | 44508931 | 44508931 | SNV | C | C:G | 1 | | 0 |
| **chr2** | 44525378 | 44525378 | SNV | G | A:G | 8 | | 0 |
| **chr2** | 44512600 | 44512600 | SNV | C | A:C | 1 | | 0 |
| **chr2** | 44535278 | 44535278 | SNV | A | A:G | 14 | | 0 |
| **chr2** | 44531523 | 44531523 | SNV | T | G:T | 1 | | 0 |
| **chr2** | 44547910 | 44547910 | SNV | G | A:G | 1 | | 0 |
| **chr2** | 44521462 | 44521468 | deletion | GGTGAAA | -:GGTGAAA | 6 | | 0 |
| **chr2** | 44547273 | 44547273 | SNV | T | A:T | 1 | | 0 |
| **chr2** | 44533610 | 44533610 | SNV | A | A:T | 4 | | 0 |
| **chr2** | 44538696 | 44538696 | SNV | T | G:T | 1 | | 0 |
| **chr2** | 44539790 | 44539790 | SNV | C\|N | C:T\|N:N | 2 | | 0 |
| **chr2** | 44521316 | 44521316 | SNV | G | C:G | 3 | | 0 |
| **chr2** | 44526092 | 44526092 | SNV | C | C:G | 2 | | 0 |
| **chr2** | 44544674 | 44544674 | SNV | C | C:T | 2 | | 0 |
| **chr2** | 44535797 | 44535797 | SNV | A | A:G | 1 | | 0 |
| **chr2** | 44520663 | 44520663 | SNV | A | A:G | 1 | | 0 |
| **chr2** | 44507567 | 44507567 | SNV | G | A:G | 5 | | 0 |
| **chr2** | 44506147 | 44506147 | SNV | T | C:T | 1 | | 0 |
| **chr2** | 44518858 | 44518858 | SNV | G | A:G | 1 | | 0 |
| **chr2** | 44533100 | 44533100 | SNV | C | C:T | 1 | | 0 |
| **chr2** | 44526242 | 44526242 | SNV | A | A:T | 4 | | 0 |
| **chr2** | 44539039 | 44539039 | SNV | G | A:G | 1 | | 0 |
| **chr2** | 44539866 | 44539866 | SNV | G\|A | A:G\|T:A | 1 | | 0 |
| **chr2** | 44525234 | 44525234 | SNV | T | C:T | 3 | | 0 |
| **chr2** | 44508340 | 44508340 | SNV | C | C:T | 1 | | 0 |
| **chr2** | 44513595 | 44513595 | SNV | C | C:T | 6 | | 0 |
| **chr2** | 44544735 | 44544735 | SNV | G | A:G | 2 | | 0 |
| **chr2** | 44537210 | 44537210 | SNV | T | G:T | 1 | | 0 |
| **chr2** | 44513168 | 44513168 | SNV | C | C:T | 1 | | 0 |
| **chr2** | 44540754 | 44540754 | SNV | T | C:T | 1 | | 0 |
| **chr2** | 44537217 | 44537217 | SNV | G | G:T | 1 | | 0 |
| **chr2** | 44522291 | 44522291 | SNV | G | G:T | 1 | | 0 |
| **chr2** | 44506706 | 44506706 | SNV | C | C:T | 1 | | 0 |
| **chr2** | 44519830 | 44519830 | SNV | A | A:G | 1 | | 0 |
| **chr2** | 44525250 | 44525250 | SNV | G | A:G | 5 | | 0 |
| **chr2** | 44545620 | 44545620 | SNV | C | C:T | 1 | | 0 |
| **chr2** | 44502964 | 44502964 | SNV | T\|V | C:T\|A:V | 2 | | 0 |
| **chr2** | 44506511 | 44506511 | SNV | C | C:T | 8 | | 0 |
| **chr2** | 44543457 | 44543457 | SNV | T | C:T | 2 | | 0 |
| **chr2** | 44515288 | 44515288 | SNV | T | C:T | 4 | | 0 |
| **chr2** | 44541539 | 44541539 | SNV | T | C:T | 1 | | 0 |
| **chr2** | 44519054 | 44519054 | SNV | G | A:G | 1 | | 0 |
| **chr2** | 44529545 | 44529545 | SNV | T | A:T | 1 | | 0 |
| **chr2** | 44547082 | 44547082 | SNV | A | A:T | 4 | | 0 |
| **chr2** | 44526486 | 44526486 | SNV | C | C:T | 1 | | 0 |

**Supplementary Table 1. Variants of SLC3A1 in the 1KG database**

| Chr | Start Position | End Position | Variant type | Ref Genotype \| Ref Proteotype | Variant Genotype 1\| Variant Proteotype 1 | Count | Variant Genotype 2\| Variant Proteotype 2 | Count |
|---|---|---|---|---|---|---|---|---|
| **chr2** | 44512887 | 44512887 | SNV | G | C:G | 2 | | 0 |
| **chr2** | 44542190 | 44542190 | SNV | C | T:T | 1 | C:T | 33 |
| **chr2** | 44524258 | 44524258 | SNV | T | A:T | 4 | | 0 |
| **chr2** | 44539465 | 44539465 | SNV | G | G:T | 1 | | 0 |
| **chr2** | 44547699 | 44547699 | SNV | C\|T | C:G\|T:R | 1 | | 0 |
| **chr2** | 44515209 | 44515209 | SNV | C | C:T | 2 | | 0 |
| **chr2** | 44516947 | 44516947 | SNV | T | G:T | 2 | | 0 |
| **chr2** | 44510894 | 44510894 | SNV | C | A:C | 4 | | 0 |
| **chr2** | 44539419 | 44539419 | SNV | G | C:G | 8 | | 0 |
| **chr2** | 44507978 | 44507978 | SNV | C\|P | A:C\|H:P | 1 | | 0 |
| **chr2** | 44502916 | 44502916 | SNV | G\|R | A:G\|H:R | 1 | | 0 |
| **chr2** | 44527813 | 44527813 | SNV | A | A:G | 1 | | 0 |
| **chr2** | 44547196 | 44547196 | SNV | A | A:T | 1 | | 0 |
| **chr2** | 44521724 | 44521724 | SNV | T | C:T | 1 | | 0 |
| **chr2** | 44514488 | 44514488 | SNV | G | C:G | 1 | | 0 |
| **chr2** | 44505045 | 44505045 | SNV | A | A:G | 33 | | 0 |
| **chr2** | 44503123 | 44503123 | SNV | T | A:T | 1 | | 0 |
| **chr2** | 44525904 | 44525904 | SNV | C | C:T | 1 | | 0 |
| **chr2** | 44503634 | 44503634 | SNV | G | A:G | 1 | | 0 |
| **chr2** | 44535323 | 44535323 | SNV | C | C:G | 3 | | 0 |
| **chr2** | 44524214 | 44524214 | SNV | A | A:G | 9 | | 0 |
| **chr2** | 44536072 | 44536072 | SNV | C | C:T | 15 | | 0 |
| **chr2** | 44527450 | 44527450 | SNV | G | G:T | 3 | | 0 |
| **chr2** | 44532885 | 44532885 | SNV | A | A:T | 1 | | 0 |
| **chr2** | 44541606 | 44541606 | SNV | T | C:T | 2 | | 0 |
| **chr2** | 44509911 | 44509911 | SNV | C | C:T | 2 | | 0 |
| **chr2** | 44506965 | 44506965 | SNV | G | A:G | 1 | | 0 |
| **chr2** | 44528099 | 44528099 | SNV | A | A:T | 1 | | 0 |
| **chr2** | 44544840 | 44544840 | SNV | A | A:G | 1 | | 0 |
| **chr2** | 44528193 | 44528193 | SNV | G\|V | A:G\|M:V | 1 | | 0 |
| **chr2** | 44528601 | 44528601 | SNV | C | C:T | 5 | | 0 |
| **chr2** | 44545291 | 44545291 | SNV | G | A:G | 1 | | 0 |
| **chr2** | 44530353 | 44530353 | insertion | - | -:A | 4 | | 0 |
| **chr2** | 44547327 | 44547327 | SNV | T | A:T | 4 | | 0 |
| **chr2** | 44538121 | 44538121 | SNV | C | A:C | 2 | | 0 |
| **chr2** | 44506000 | 44506000 | SNV | A | A:G | 1 | | 0 |
| **chr2** | 44546593 | 44546593 | SNV | G | C:G | 8 | | 0 |
| **chr2** | 44505064 | 44505064 | SNV | A | A:C | 1 | | 0 |
| **chr2** | 44547931 | 44547931 | SNV | T | C:T | 1 | | 0 |
| **chr2** | 44506992 | 44506992 | SNV | A | C:C | 1 | A:C | 7 |
| **chr2** | 44530065 | 44530065 | SNV | C | C:T | 2 | | 0 |
| **chr2** | 44510127 | 44510127 | SNV | C | C:T | 1 | | 0 |
| **chr2** | 44525201 | 44525201 | SNV | C | C:T | 7 | | 0 |
| **chr2** | 44519049 | 44519049 | SNV | G | A:G | 61 | A:A | 2 |
| **chr2** | 44518802 | 44518802 | SNV | G | A:G | 6 | | 0 |
| **chr2** | 44505492 | 44505492 | SNV | C | C:T | 1 | | 0 |
| **chr2** | 44531251 | 44531251 | SNV | C | C:T | 4 | | 0 |
| **chr2** | 44546516 | 44546516 | SNV | A | A:G | 1 | | 0 |
| **chr2** | 44505410 | 44505410 | SNV | C | C:T | 3 | | 0 |
| **chr2** | 44518084 | 44518084 | SNV | C | C:T | 1 | | 0 |
| **chr2** | 44508200 | 44508200 | SNV | C | C:T | 15 | | 0 |
| **chr2** | 44529897 | 44529897 | SNV | C | C:T | 1 | | 0 |
| **chr2** | 44530195 | 44530195 | SNV | C | C:T | 7 | | 0 |
| **chr2** | 44532522 | 44532522 | SNV | G | A:G | 4 | | 0 |
| **chr2** | 44503202 | 44503202 | SNV | A | A:G | 1 | | 0 |
| **chr2** | 44546648 | 44546648 | SNV | A | A:C | 1 | | 0 |
| **chr2** | 44529506 | 44529506 | SNV | A | A:G | 1 | | 0 |
| **chr2** | 44522828 | 44522828 | SNV | T | C:T | 9 | | 0 |
| **chr2** | 44515502 | 44515502 | SNV | C | C:T | 1 | | 0 |
| **chr2** | 44514534 | 44514534 | SNV | G | A:A | 2 | A:G | 60 |
| **chr2** | 44516418 | 44516418 | SNV | C | C:T | 3 | | 0 |
| **chr2** | 44541668 | 44541668 | SNV | T | A:T | 1 | | 0 |
| **chr2** | 44544246 | 44544247 | deletion | CT | -:CT | 8 | | 0 |
| **chr2** | 44540855 | 44540855 | SNV | G | A:G | 1 | | 0 |
| **chr2** | 44520650 | 44520650 | SNV | C | C:T | 1 | | 0 |
| **chr2** | 44536061 | 44536061 | SNV | C | C:T | 3 | | 0 |
| **chr2** | 44540745 | 44540745 | SNV | C | C:T | 3 | | 0 |
| **chr2** | 44527871 | 44527871 | SNV | C | C:T | 2 | | 0 |
| **chr2** | 44547122 | 44547122 | SNV | T | C:T | 1 | | 0 |
| **chr2** | 44522036 | 44522036 | insertion | - | -:T | 24 | | 0 |
| **chr2** | 44534232 | 44534232 | SNV | C | C:G | 1 | | 0 |
| **chr2** | 44533997 | 44533997 | SNV | A | A:C | 10 | | 0 |
| **chr2** | 44536081 | 44536081 | SNV | C | C:T | 1 | | 0 |
| **chr2** | 44534564 | 44534564 | SNV | C | A:C | 8 | | 0 |
| **chr2** | 44542660 | 44542660 | SNV | G | C:G | 1 | | 0 |
| **chr2** | 44524652 | 44524652 | SNV | T | G:T | 8 | | 0 |
| **chr2** | 44547263 | 44547263 | SNV | T | G:T | 5 | | 0 |
| **chr2** | 44502987 | 44502987 | SNV | A\|I | A:G\|I:V | 2 | | 0 |
| **chr2** | 44513854 | 44513854 | SNV | T | C:T | 9 | | 0 |
| **chr2** | 44513637 | 44513637 | SNV | C | C:T | 1 | | 0 |
| **chr2** | 44515434 | 44515434 | SNV | G | C:G | 8 | | 0 |
| **chr2** | 44503745 | 44503745 | SNV | G | A:G | 1 | | 0 |
| **chr2** | 44544571 | 44544571 | SNV | A | A:G | 1 | | 0 |
| **chr2** | 44546571 | 44546571 | SNV | C | A:C | 1 | | 0 |
| **chr2** | 44541415 | 44541415 | SNV | C | A:C | 1 | | 0 |
| **chr2** | 44526721 | 44526721 | SNV | T | C:T | 5 | | 0 |
| **chr2** | 44507578 | 44507578 | SNV | A | A:T | 1 | | 0 |
| **chr2** | 44527352 | 44527356 | deletion | TGATT | -:TGATT | 3 | | 0 |
| **chr2** | 44532707 | 44532707 | SNV | C | C:G | 1 | | 0 |
| **chr2** | 44527697 | 44527697 | SNV | A | A:G | 2 | | 0 |
| **chr2** | 44541751 | 44541751 | SNV | T | A:T | 33 | A:A | 1 |
| **chr2** | 44524281 | 44524281 | SNV | C | C:T | 1 | | 0 |
| **chr2** | 44510145 | 44510145 | SNV | G | G:T | 1 | | 0 |
| **chr2** | 44520361 | 44520361 | SNV | G | A:G | 2 | | 0 |

**Supplementary Table 1. Variants of SLC3A1 in the 1KG database**

| Chr | Start Position | End Position | Variant type | Ref Genotype \| Ref Proteotype | Variant Genotype 1\| Variant Proteotype 1 | Count | Variant Genotype 2\| Variant Proteotype 2 | Count |
|---|---|---|---|---|---|---|---|---|
| **chr2** | 44547487 | 44547487 | SNV | C\|I | C:T\|I:I | 1 | | 0 |
| **chr2** | 44515349 | 44515349 | SNV | G | C:G | 1 | | 0 |
| **chr2** | 44516692 | 44516692 | SNV | T | C:T | 1 | | 0 |
| **chr2** | 44539782 | 44539782 | SNV | G\|V | A:G\|M:V | 1 | | 0 |
| **chr2** | 44525968 | 44525968 | SNV | G | G:T | 1 | | 0 |
| **chr2** | 44505467 | 44505467 | SNV | T | G:T | 1 | | 0 |
| **chr2** | 44530754 | 44530754 | SNV | T | C:T | 1 | | 0 |
| **chr2** | 44515546 | 44515546 | SNV | C | A:C | 1 | | 0 |
| **chr2** | 44534777 | 44534777 | SNV | T | C:T | 1 | | 0 |
| **chr2** | 44538901 | 44538901 | SNV | A | A:G | 1 | | 0 |
| **chr2** | 44503231 | 44503231 | SNV | A | A:C | 1 | | 0 |
| **chr2** | 44524988 | 44524988 | SNV | C | C:T | 1 | | 0 |
| **chr2** | 44545878 | 44545878 | SNV | G | C:G | 1 | | 0 |
| **chr2** | 44545877 | 44545877 | SNV | T | A:T | 2 | | 0 |
| **chr2** | 44547172 | 44547172 | SNV | T | C:T | 1 | | 0 |
| **chr2** | 44506019 | 44506019 | SNV | C | C:G | 1 | | 0 |
| **chr2** | 44531514 | 44531514 | SNV | C | A:C | 1 | | 0 |
| **chr2** | 44529153 | 44529153 | SNV | C | C:T | 1 | | 0 |
| **chr2** | 44539898 | 44539898 | SNV | T | C:T | 1 | | 0 |
| **chr2** | 44547115 | 44547115 | SNV | G | A:G | 1 | | 0 |
| **chr2** | 44521929 | 44521929 | SNV | C | C:T | 1 | | 0 |
| **chr2** | 44534083 | 44534083 | SNV | A | A:C | 1 | | 0 |
| **chr2** | 44517281 | 44517281 | SNV | C | A:C | 2 | | 0 |
| **chr2** | 44534155 | 44534155 | SNV | C | A:C | 1 | | 0 |
| **chr2** | 44520943 | 44520943 | SNV | C | A:C | 1 | | 0 |
| **chr2** | 44504505 | 44504505 | SNV | C | C:T | 1 | | 0 |
| **chr2** | 44545421 | 44545421 | SNV | G | A:G | 1 | | 0 |
| **chr2** | 44502924 | 44502924 | SNV | A\|I | A:G\|I:V | 1 | | 0 |
| **chr2** | 44533046 | 44533046 | SNV | C | C:T | 3 | | 0 |
| **chr2** | 44511469 | 44511469 | SNV | G | A:G | 1 | | 0 |
| **chr2** | 44536341 | 44536341 | SNV | A | A:C | 1 | | 0 |
| **chr2** | 44522812 | 44522812 | SNV | A | A:C | 1 | | 0 |
| **chr2** | 44512036 | 44512036 | SNV | A | A:C | 1 | | 0 |
| **chr2** | 44544122 | 44544122 | SNV | C | C:G | 1 | | 0 |
| **chr2** | 44521572 | 44521572 | SNV | A | A:C | 1 | | 0 |
| **chr2** | 44506418 | 44506418 | SNV | C | C:T | 1 | | 0 |
| **chr2** | 44518662 | 44518662 | SNV | A | A:G | 1 | | 0 |
| **chr2** | 44506493 | 44506493 | SNV | C | C:T | 2 | | 0 |
| **chr2** | 44503050 | 44503050 | SNV | A\|I | A:G\|I:V | 1 | | 0 |
| **chr2** | 44516575 | 44516575 | SNV | C | C:T | 1 | | 0 |
| **chr2** | 44503652 | 44503652 | SNV | C | C:T | 1 | | 0 |
| **chr2** | 44514076 | 44514076 | SNV | T | G:T | 1 | | 0 |
| **chr2** | 44531279 | 44531279 | SNV | T | C:T | 1 | | 0 |
| **chr2** | 44508743 | 44508743 | SNV | T | G:T | 1 | | 0 |
| **chr2** | 44504125 | 44504125 | SNV | G | A:G | 2 | | 0 |
| **chr2** | 44512431 | 44512431 | SNV | T | A:T | 1 | | 0 |
| **chr2** | 44506924 | 44506924 | SNV | A | A:G | 1 | | 0 |
| **chr2** | 44503675 | 44503675 | SNV | G | C:G | 1 | | 0 |
| **chr2** | 44530318 | 44530318 | SNV | C | C:T | 1 | | 0 |
| **chr2** | 44517260 | 44517260 | SNV | G | A:G | 1 | | 0 |
| **chr2** | 44513092 | 44513092 | SNV | G | G:T | 6 | | 0 |
| **chr2** | 44525413 | 44525413 | SNV | T | C:T | 3 | | 0 |
| **chr2** | 44514773 | 44514773 | SNV | C | C:T | 1 | | 0 |
| **chr2** | 44503091 | 44503091 | SNV | C\|N | C:T\|N:N | 2 | | 0 |
| **chr2** | 44503041 | 44503041 | SNV | A\|M | A:G\|M:V | 2 | | 0 |
| **chr2** | 44525750 | 44525750 | SNV | C | C:T | 1 | | 0 |
| **chr2** | 44533277 | 44533277 | SNV | T | G:T | 1 | | 0 |
| **chr2** | 44527164 | 44527164 | SNV | G\|V | A:G\|I:V | 1 | | 0 |
| **chr2** | 44546480 | 44546480 | SNV | A | A:G | 1 | | 0 |
| **chr2** | 44532018 | 44532018 | SNV | T | C:T | 2 | | 0 |
| **chr2** | 44531775 | 44531775 | SNV | T | C:T | 1 | | 0 |
| **chr2** | 44538902 | 44538902 | SNV | G | C:G | 2 | | 0 |
| **chr2** | 44534441 | 44534441 | SNV | G | G:T | 1 | | 0 |
| **chr2** | 44516473 | 44516473 | SNV | C | A:C | 1 | | 0 |
| **chr2** | 44538741 | 44538741 | SNV | A | A:G | 1 | | 0 |
| **chr2** | 44542323 | 44542323 | SNV | C | C:T | 1 | | 0 |
| **chr2** | 44525287 | 44525287 | SNV | T | C:T | 1 | | 0 |
| **chr2** | 44544502 | 44544502 | SNV | G | A:G | 2 | | 0 |
| **chr2** | 44521738 | 44521738 | SNV | A | A:C | 1 | | 0 |
| **chr2** | 44546701 | 44546701 | SNV | T | C:T | 1 | | 0 |
| **chr2** | 44505172 | 44505172 | SNV | C | C:G | 1 | | 0 |
| **chr2** | 44515765 | 44515765 | SNV | G | A:G | 1 | | 0 |
| **chr2** | 44535730 | 44535730 | SNV | A | A:G | 1 | | 0 |
| **chr2** | 44532715 | 44532715 | SNV | C | C:T | 1 | | 0 |
| **chr2** | 44546179 | 44546179 | SNV | T | C:T | 3 | | 0 |
| **chr2** | 44522215 | 44522215 | SNV | G | G:T | 1 | | 0 |
| **chr2** | 44514957 | 44514957 | SNV | C | A:C | 1 | | 0 |
| **chr2** | 44526813 | 44526813 | SNV | C | C:G | 5 | | 0 |
| **chr2** | 44516579 | 44516579 | SNV | G | A:G | 1 | | 0 |
| **chr2** | 44524528 | 44524528 | SNV | G | C:G | 1 | | 0 |
| **chr2** | 44505039 | 44505039 | SNV | T | C:T | 1 | | 0 |
| **chr2** | 44528161 | 44528161 | SNV | C\|S | C:T\|S:L | 2 | | 0 |
| **chr2** | 44527124 | 44527124 | SNV | C\|F | C:T\|F:F | 1 | | 0 |
| **chr2** | 44526977 | 44526977 | SNV | T | G:T | 3 | | 0 |
| **chr2** | 44518406 | 44518406 | SNV | C | C:T | 1 | | 0 |
| **chr2** | 44506821 | 44506821 | SNV | A | A:T | 1 | | 0 |
| **chr2** | 44527882 | 44527882 | SNV | C | C:T | 2 | | 0 |
| **chr2** | 44541457 | 44541457 | SNV | G | A:G | 1 | | 0 |
| **chr2** | 44537770 | 44537770 | SNV | G | C:G | 1 | | 0 |
| **chr2** | 44547624 | 44547624 | SNV | C\|T | C:T\|T:I | 1 | | 0 |
| **chr2** | 44535397 | 44535397 | SNV | T | G:T | 1 | | 0 |
| **chr2** | 44533294 | 44533294 | SNV | T | C:T | 1 | | 0 |
| **chr2** | 44545876 | 44545876 | SNV | T | C:T | 1 | | 0 |
| **chr2** | 44532045 | 44532045 | SNV | C | C:T | 1 | | 0 |

**Supplementary Table 1. Variants of SLC3A1 in the 1KG database**

| Chr | Start Position | End Position | Variant type | Ref Genotype \| Ref Proteotype | Variant Genotype 1\| Variant Proteotype 1 | Count | Variant Genotype 2\| Variant Proteotype 2 | Count |
|---|---|---|---|---|---|---|---|---|
| **chr2** | 44521471 | 44521471 | SNV | C | C:G | 1 | | 0 |
| **chr2** | 44538447 | 44538447 | SNV | T | C:T | 1 | | 0 |
| **chr2** | 44544866 | 44544866 | SNV | A | A:T | 3 | | 0 |
| **chr2** | 44502685 | 44502685 | SNV | A\|D | A:G\|D:G | 1 | | 0 |
| **chr2** | 44528777 | 44528777 | SNV | C | C:T | 1 | | 0 |
| **chr2** | 44509158 | 44509158 | SNV | G | A:G | 1 | | 0 |
| **chr2** | 44524307 | 44524307 | SNV | G | C:G | 1 | | 0 |
| **chr2** | 44520501 | 44520501 | SNV | C | C:G | 1 | | 0 |
| **chr2** | 44544706 | 44544706 | SNV | T | G:T | 1 | | 0 |
| **chr2** | 44507569 | 44507569 | SNV | G | A:G | 1 | | 0 |
| **chr2** | 44522609 | 44522609 | SNV | C | C:G | 1 | | 0 |
| **chr2** | 44509118 | 44509118 | SNV | C | A:C | 1 | | 0 |
| **chr2** | 44505510 | 44505510 | SNV | C | A:C | 7 | | 0 |
| **chr2** | 44506661 | 44506661 | SNV | T | A:T | 2 | | 0 |
| **chr2** | 44511395 | 44511395 | SNV | C | C:G | 1 | | 0 |
| **chr2** | 44517504 | 44517504 | SNV | A | A:G | 1 | | 0 |
| **chr2** | 44507570 | 44507570 | SNV | G | G:T | 2 | | 0 |
| **chr2** | 44513785 | 44513785 | SNV | C | C:T | 1 | | 0 |
| **chr2** | 44537442 | 44537442 | SNV | C | A:C | 1 | | 0 |
| **chr2** | 44537570 | 44537570 | SNV | C | A:C | 1 | | 0 |
| **chr2** | 44528305 | 44528305 | SNV | A | A:T | 1 | | 0 |
| **chr2** | 44537440 | 44537440 | SNV | T | G:T | 1 | | 0 |
| **chr2** | 44529349 | 44529349 | SNV | C | A:C | 1 | | 0 |
| **chr2** | 44531086 | 44531086 | SNV | G | A:G | 1 | | 0 |
| **chr2** | 44512678 | 44512678 | SNV | T | A:T | 1 | | 0 |
| **chr2** | 44510920 | 44510920 | SNV | A | A:G | 1 | | 0 |
| **chr2** | 44528076 | 44528076 | SNV | T | C:T | 1 | | 0 |
| **chr2** | 44530223 | 44530223 | SNV | C | C:G | 1 | | 0 |
| **chr2** | 44521845 | 44521845 | SNV | C | C:T | 1 | | 0 |
| **chr2** | 44503659 | 44503659 | SNV | A | A:G | 3 | | 0 |
| **chr2** | 44504047 | 44504047 | SNV | T | C:T | 1 | | 0 |
| **chr2** | 44534527 | 44534527 | SNV | C | C:G | 1 | | 0 |
| **chr2** | 44541927 | 44541927 | SNV | G | C:G | 1 | | 0 |
| **chr2** | 44518774 | 44518774 | SNV | C | A:C | 1 | | 0 |
| **chr2** | 44514213 | 44514213 | SNV | C | C:T | 1 | | 0 |
| **chr2** | 44544523 | 44544523 | SNV | C | C:G | 2 | | 0 |
| **chr2** | 44510386 | 44510386 | SNV | C | A:C | 1 | | 0 |
| **chr2** | 44520643 | 44520643 | SNV | A | A:C | 1 | | 0 |
| **chr2** | 44503857 | 44503857 | SNV | G | A:G | 2 | | 0 |
| **chr2** | 44547609 | 44547609 | SNV | G\|G | A:G\|D:G | 11 | | 0 |
| **chr2** | 44519690 | 44519690 | SNV | C | C:T | 2 | | 0 |
| **chr2** | 44527093 | 44527093 | SNV | T | G:T | 2 | | 0 |
| **chr2** | 44546084 | 44546084 | SNV | A | A:C | 2 | | 0 |
| **chr2** | 44505000 | 44505000 | SNV | G | A:G | 1 | | 0 |
| **chr2** | 44524898 | 44524898 | SNV | G | A:G | 1 | | 0 |
| **chr2** | 44524179 | 44524179 | SNV | C | A:C | 2 | | 0 |
| **chr2** | 44528359 | 44528359 | SNV | A | A:G | 1 | | 0 |
| **chr2** | 44514221 | 44514221 | SNV | G | C:G | 1 | | 0 |
| **chr2** | 44528686 | 44528686 | SNV | T | A:T | 1 | | 0 |
| **chr2** | 44545296 | 44545296 | SNV | C | C:T | 2 | | 0 |
| **chr2** | 44529791 | 44529791 | SNV | C | A:C | 1 | | 0 |
| **chr2** | 44502996 | 44502996 | SNV | A\|I | A:G\|I:V | 1 | | 0 |
| **chr2** | 44534925 | 44534925 | SNV | C | C:G | 1 | | 0 |
| **chr2** | 44536385 | 44536385 | SNV | T | A:T | 1 | | 0 |
| **chr2** | 44505083 | 44505083 | SNV | C | C:T | 1 | | 0 |
| **chr2** | 44536646 | 44536646 | SNV | C | C:G | 1 | | 0 |
| **chr2** | 44543371 | 44543371 | SNV | G | A:G | 1 | | 0 |
| **chr2** | 44536332 | 44536332 | SNV | T | G:T | 1 | | 0 |
| **chr2** | 44529118 | 44529118 | SNV | G | A:G | 1 | | 0 |
| **chr2** | 44515034 | 44515034 | SNV | C | A:C | 1 | | 0 |
| **chr2** | 44508059 | 44508059 | SNV | G | A:G | 1 | | 0 |
| **chr2** | 44547546 | 44547546 | SNV | A\|N | A:G\|N:S | 2 | | 0 |
| **chr2** | 44508722 | 44508722 | SNV | G | A:G | 1 | | 0 |
| **chr2** | 44533287 | 44533287 | SNV | G | C:G | 2 | | 0 |
| **chr2** | 44515737 | 44515737 | SNV | T | A:T | 1 | | 0 |
| **chr2** | 44525839 | 44525839 | SNV | T | A:T | 1 | | 0 |
| **chr2** | 44546371 | 44546371 | SNV | C | C:G | 2 | | 0 |
| **chr2** | 44520612 | 44520612 | SNV | T | C:T | 1 | | 0 |
| **chr2** | 44540658 | 44540658 | SNV | G | A:G | 5 | | 0 |
| **chr2** | 44528244 | 44528244 | SNV | A\|S | A:G\|S:G | 1 | | 0 |
| **chr2** | 44544865 | 44544865 | SNV | G | A:G | 3 | | 0 |
| **chr2** | 44502931 | 44502931 | SNV | G\|R | A:G\|Q:R | 2 | | 0 |
| **chr2** | 44538533 | 44538533 | SNV | A | A:G | 1 | | 0 |
| **chr2** | 44502972 | 44502972 | SNV | C\|L | C:T\|L:F | 1 | | 0 |
| **chr2** | 44503859 | 44503859 | SNV | G | G:T | 1 | | 0 |
| **chr2** | 44511823 | 44511823 | SNV | C | C:T | 1 | | 0 |
| **chr2** | 44522724 | 44522724 | SNV | A | A:G | 1 | | 0 |
| **chr2** | 44527512 | 44527512 | SNV | T | C:T | 1 | | 0 |
| **chr2** | 44539545 | 44539545 | SNV | A | A:C | 1 | | 0 |
| **chr2** | 44514183 | 44514183 | SNV | T | G:T | 1 | | 0 |
| **chr2** | 44525203 | 44525203 | SNV | T | A:T | 1 | | 0 |
| **chr2** | 44528104 | 44528104 | SNV | A | A:T | 1 | | 0 |
| **chr2** | 44525256 | 44525256 | SNV | T | C:T | 1 | | 0 |
| **chr2** | 44510768 | 44510768 | SNV | A | A:G | 1 | | 0 |
| **chr2** | 44528478 | 44528478 | SNV | T | C:T | 1 | | 0 |
| **chr2** | 44503398 | 44503398 | SNV | G | G:T | 1 | | 0 |
| **chr2** | 44531676 | 44531676 | SNV | A | A:T | 1 | | 0 |
| **chr2** | 44545771 | 44545771 | SNV | T | A:T | 1 | | 0 |
| **chr2** | 44511215 | 44511215 | SNV | C | C:T | 2 | | 0 |
| **chr2** | 44516723 | 44516723 | SNV | T | C:T | 1 | | 0 |
| **chr2** | 44544946 | 44544946 | SNV | A | A:G | 1 | | 0 |
| **chr2** | 44522055 | 44522055 | SNV | G | C:G | 1 | | 0 |
| **chr2** | 44535550 | 44535550 | SNV | A | A:T | 1 | | 0 |
| **chr2** | 44525900 | 44525900 | SNV | G | G:T | 1 | | 0 |

**Supplementary Table 1. Variants of SLC3A1 in the 1KG database**

| Chr | Start Position | End Position | Variant type | Ref Genotype \| Ref Proteotype | Variant Genotype 1\| Variant Proteotype 1 | Count | Variant Genotype 2\| Variant Proteotype 2 | Count |
|---|---|---|---|---|---|---|---|---|
| **chr2** | 44536062 | 44536062 | SNV | G | A:G | 1 | | 0 |
| **chr2** | 44504888 | 44504888 | SNV | C | C:T | 1 | | 0 |
| **chr2** | 44520428 | 44520428 | SNV | C | C:G | 2 | | 0 |
| **chr2** | 44543700 | 44543700 | SNV | T | C:T | 1 | | 0 |
| **chr2** | 44538114 | 44538114 | SNV | T | C:T | 1 | | 0 |
| **chr2** | 44539178 | 44539178 | SNV | T | G:T | 1 | | 0 |
| **chr2** | 44508129 | 44508129 | SNV | A | A:G | 2 | | 0 |
| **chr2** | 44512717 | 44512717 | SNV | A | A:G | 1 | | 0 |
| **chr2** | 44507996 | 44507996 | SNV | A\|E | A:G\|E:G | 1 | | 0 |
| **chr2** | 44537807 | 44537807 | SNV | A | A:C | 1 | | 0 |
| **chr2** | 44513478 | 44513478 | SNV | A | A:G | 1 | | 0 |
| **chr2** | 44521337 | 44521337 | SNV | T | C:T | 1 | | 0 |
| **chr2** | 44531235 | 44531235 | SNV | G | A:G | 1 | | 0 |
| **chr2** | 44534330 | 44534330 | SNV | T | C:T | 1 | | 0 |
| **chr2** | 44519579 | 44519579 | SNV | G | C:G | 1 | | 0 |
| **chr2** | 44525000 | 44525000 | SNV | T | C:T | 1 | | 0 |
| **chr2** | 44544326 | 44544326 | SNV | G | G:T | 1 | | 0 |
| **chr2** | 44528788 | 44528788 | SNV | G | A:G | 1 | | 0 |
| **chr2** | 44530746 | 44530746 | SNV | T | G:T | 1 | | 0 |
| **chr2** | 44512693 | 44512693 | SNV | A | A:G | 1 | | 0 |
| **chr2** | 44547854 | 44547854 | SNV | T | G:T | 1 | | 0 |
| **chr2** | 44532317 | 44532317 | SNV | C | C:G | 1 | | 0 |
| **chr2** | 44515998 | 44515998 | SNV | G | C:G | 1 | | 0 |
| **chr2** | 44505112 | 44505112 | SNV | A | A:T | 1 | | 0 |
| **chr2** | 44535000 | 44535000 | SNV | G | C:G | 1 | | 0 |
| **chr2** | 44526630 | 44526630 | SNV | C | C:T | 1 | | 0 |
| **chr2** | 44535549 | 44535549 | SNV | G | C:G | 1 | | 0 |
| **chr2** | 44532265 | 44532265 | SNV | C | C:T | 1 | | 0 |
| **chr2** | 44506297 | 44506297 | SNV | C | C:G | 1 | | 0 |
| **chr2** | 44545290 | 44545290 | SNV | C | C:T | 1 | | 0 |
| **chr2** | 44514640 | 44514640 | SNV | G | G:T | 1 | | 0 |
| **chr2** | 44547258 | 44547258 | SNV | A | A:G | 1 | | 0 |
| **chr2** | 44524047 | 44524047 | SNV | C | C:T | 1 | | 0 |
| **chr2** | 44547222 | 44547222 | SNV | A | A:G | 1 | | 0 |
| **chr2** | 44507965 | 44507965 | SNV | C\|R | C:T\|R:W | 2 | | 0 |
| **chr2** | 44524947 | 44524947 | SNV | G | A:G | 4 | | 0 |
| **chr2** | 44544345 | 44544345 | SNV | T | C:T | 1 | | 0 |
| **chr2** | 44546039 | 44546039 | SNV | A | A:G | 1 | | 0 |
| **chr2** | 44543660 | 44543660 | SNV | A | A:G | 1 | | 0 |
| **chr2** | 44543412 | 44543412 | SNV | A | A:T | 1 | | 0 |
| **chr2** | 44520593 | 44520593 | SNV | T | C:T | 2 | | 0 |
| **chr2** | 44536737 | 44536737 | SNV | C | C:T | 1 | | 0 |
| **chr2** | 44511258 | 44511258 | SNV | G | A:G | 4 | | 0 |
| **chr2** | 44503113 | 44503113 | SNV | C | C:T | 1 | | 0 |
| **chr2** | 44509874 | 44509874 | SNV | G | A:G | 1 | | 0 |
| **chr2** | 44529890 | 44529890 | SNV | A | A:G | 1 | | 0 |
| **chr2** | 44525171 | 44525171 | SNV | T | C:T | 1 | | 0 |
| **chr2** | 44544019 | 44544019 | SNV | A | A:G | 1 | | 0 |
| **chr2** | 44534803 | 44534803 | SNV | G | G:T | 3 | | 0 |
| **chr2** | 44547135 | 44547135 | SNV | A | A:G | 1 | | 0 |
| **chr2** | 44537551 | 44537551 | SNV | G | G:T | 1 | | 0 |
| **chr2** | 44519513 | 44519513 | SNV | G | A:G | 2 | | 0 |
| **chr2** | 44513046 | 44513046 | SNV | C | C:G | 1 | | 0 |
| **chr2** | 44525375 | 44525375 | SNV | C | C:G | 2 | | 0 |
| **chr2** | 44510662 | 44510662 | SNV | T | A:T | 1 | | 0 |
| **chr2** | 44541133 | 44541133 | SNV | A | A:G | 1 | | 0 |
| **chr2** | 44515409 | 44515409 | SNV | C | A:C | 1 | | 0 |
| **chr2** | 44539264 | 44539264 | SNV | A | A:G | 1 | | 0 |
| **chr2** | 44534808 | 44534808 | SNV | G | C:G | 1 | | 0 |
| **chr2** | 44546958 | 44546958 | SNV | G | A:G | 1 | | 0 |
| **chr2** | 44532081 | 44532081 | SNV | C | C:T | 1 | | 0 |
| **chr2** | 44524498 | 44524498 | SNV | T | A:T | 1 | | 0 |
| **chr2** | 44508507 | 44508507 | SNV | A | A:C | 1 | | 0 |
| **chr2** | 44529593 | 44529593 | SNV | C | C:T | 10 | | 0 |
| **chr2** | 44508849 | 44508849 | SNV | C | C:T | 1 | | 0 |
| **chr2** | 44539685 | 44539685 | SNV | T | C:T | 1 | | 0 |
| **chr2** | 44521951 | 44521951 | SNV | C | A:C | 1 | | 0 |
| **chr2** | 44540141 | 44540141 | SNV | C | T:T | 1 | | 0 |
| **chr2** | 44524860 | 44524860 | SNV | G | G:T | 1 | | 0 |
| **chr2** | 44546566 | 44546566 | SNV | A | A:G | 1 | | 0 |
| **chr2** | 44545005 | 44545005 | SNV | G | A:G | 1 | | 0 |
| **chr2** | 44527082 | 44527082 | SNV | A | A:G | 2 | | 0 |
| **chr2** | 44536259 | 44536259 | SNV | T | C:T | 1 | | 0 |
| **chr2** | 44530872 | 44530872 | SNV | C | C:T | 1 | | 0 |
| **chr2** | 44520215 | 44520215 | SNV | T | C:T | 1 | | 0 |
| **chr2** | 44505235 | 44505235 | SNV | G | G:T | 1 | | 0 |
| **chr2** | 44519072 | 44519072 | SNV | T | C:T | 1 | | 0 |
| **chr2** | 44506075 | 44506075 | SNV | G | A:G | 1 | | 0 |
| **chr2** | 44518564 | 44518564 | SNV | G | C:G | 1 | | 0 |
| **chr2** | 44505918 | 44505918 | SNV | C | A:C | 1 | | 0 |
| **chr2** | 44527343 | 44527343 | SNV | C | A:C | 1 | | 0 |
| **chr2** | 44536039 | 44536039 | SNV | C | A:C | 1 | | 0 |
| **chr2** | 44507597 | 44507597 | SNV | C | C:T | 1 | | 0 |
| **chr2** | 44540965 | 44540965 | SNV | T | C:T | 1 | | 0 |
| **chr2** | 44524023 | 44524023 | SNV | A | A:C | 1 | | 0 |
| **chr2** | 44526034 | 44526034 | SNV | T | A:T | 1 | | 0 |
| **chr2** | 44518058 | 44518058 | SNV | C | C:T | 1 | | 0 |
| **chr2** | 44510779 | 44510779 | SNV | C | C:T | 1 | | 0 |
| **chr2** | 44502640 | 44502640 | SNV | C | C:T | 1 | | 0 |
| **chr2** | 44543731 | 44543731 | SNV | A | A:G | 1 | | 0 |
| **chr2** | 44540534 | 44540534 | SNV | C | A:C | 1 | | 0 |
| **chr2** | 44511280 | 44511280 | SNV | G | A:G | 1 | | 0 |
| **chr2** | 44508587 | 44508587 | SNV | A\|Q | A:G\|Q:Q | 7 | | 0 |
| **chr2** | 44502675 | 44502675 | SNV | A\|M | A:G\|M:V | 1 | | 0 |

**Supplementary Table 1. Variants of SLC3A1 in the 1KG database**

| Chr | Start Position | End Position | Variant type | Ref Genotype \| Ref Proteotype | Variant Genotype 1\| Variant Proteotype 1 | Count | Variant Genotype 2\| Variant Proteotype 2 | Count |
|---|---|---|---|---|---|---|---|---|
| chr2 | 44531908 | 44531908 | SNV | G | A:G | 1 | | 0 |
| chr2 | 44532650 | 44532650 | SNV | T | G:T | 1 | | 0 |
| chr2 | 44524062 | 44524062 | SNV | A | A:G | 1 | | 0 |
| chr2 | 44506498 | 44506498 | SNV | C | C:G | 2 | | 0 |
| chr2 | 44530771 | 44530771 | SNV | C | C:T | 2 | | 0 |
| chr2 | 44543476 | 44543476 | SNV | G | A:G | 2 | | 0 |
| chr2 | 44504671 | 44504671 | SNV | C | C:T | 8 | | 0 |
| chr2 | 44536170 | 44536170 | SNV | C | C:G | 1 | | 0 |
| chr2 | 44518577 | 44518577 | SNV | A | A:T | 7 | | 0 |
| chr2 | 44539297 | 44539297 | SNV | T | G:T | 1 | | 0 |
| chr2 | 44527613 | 44527613 | SNV | T | G:T | 1 | | 0 |
| chr2 | 44533351 | 44533351 | SNV | C | C:G | 1 | | 0 |
| chr2 | 44537195 | 44537195 | SNV | G | A:G | 1 | | 0 |
| chr2 | 44535640 | 44535640 | SNV | T | C:T | 1 | | 0 |
| chr2 | 44537784 | 44537784 | SNV | T | G:T | 1 | | 0 |
| chr2 | 44543484 | 44543484 | SNV | C | C:T | 1 | | 0 |
| chr2 | 44520227 | 44520227 | SNV | G | A:G | 1 | | 0 |
| chr2 | 44512857 | 44512857 | SNV | A | A:C | 1 | | 0 |
| chr2 | 44513589 | 44513589 | SNV | T | C:T | 1 | | 0 |
| chr2 | 44530734 | 44530734 | SNV | T | C:T | 1 | | 0 |
| chr2 | 44524406 | 44524406 | SNV | T | C:T | 1 | | 0 |
| chr2 | 44502949 | 44502949 | SNV | T\|L | C:T\|P:L | 1 | | 0 |
| chr2 | 44510787 | 44510787 | SNV | C | C:T | 1 | | 0 |
| chr2 | 44524035 | 44524035 | SNV | T | G:T | 1 | | 0 |
| chr2 | 44535965 | 44535965 | SNV | G | G:T | 1 | | 0 |
| chr2 | 44530267 | 44530267 | SNV | G | G:T | 3 | | 0 |
| chr2 | 44526548 | 44526548 | SNV | C | C:G | 1 | | 0 |
| chr2 | 44542224 | 44542224 | SNV | G | G:T | 1 | | 0 |
| chr2 | 44516997 | 44516997 | SNV | T | G:T | 1 | | 0 |
| chr2 | 44508552 | 44508552 | SNV | C\|P | C:G\|P:A | 1 | | 0 |
| chr2 | 44527132 | 44527132 | SNV | C\|T | A:C\|K:T | 1 | | 0 |
| chr2 | 44525153 | 44525153 | SNV | T | C:T | 1 | | 0 |
| chr2 | 44524092 | 44524092 | SNV | C | C:T | 1 | | 0 |
| chr2 | 44531211 | 44531211 | SNV | A | A:G | 1 | | 0 |
| chr2 | 44513352 | 44513352 | SNV | T | C:T | 2 | | 0 |
| chr2 | 44529098 | 44529098 | SNV | G | A:G | 1 | | 0 |
| chr2 | 44531547 | 44531547 | SNV | G | C:G | 1 | | 0 |
| chr2 | 44544639 | 44544639 | SNV | C | A:C | 1 | | 0 |
| chr2 | 44504405 | 44504405 | SNV | C | A:C | 1 | | 0 |
| chr2 | 44513397 | 44513397 | SNV | C | C:T | 1 | | 0 |
| chr2 | 44527743 | 44527743 | SNV | G | G:T | 1 | | 0 |
| chr2 | 44528288 | 44528288 | SNV | C | C:T | 1 | | 0 |
| chr2 | 44509259 | 44509259 | SNV | C | A:C | 1 | | 0 |
| chr2 | 44519297 | 44519297 | SNV | G | C:G | 1 | | 0 |
| chr2 | 44525908 | 44525908 | SNV | C | C:T | 1 | | 0 |
| chr2 | 44528032 | 44528032 | SNV | C | C:T | 1 | | 0 |
| chr2 | 44505604 | 44505604 | SNV | T | C:T | 1 | | 0 |
| chr2 | 44514373 | 44514373 | SNV | C | C:T | 1 | | 0 |
| chr2 | 44533230 | 44533230 | SNV | G | C:G | 1 | | 0 |
| chr2 | 44522781 | 44522781 | SNV | T | C:T | 1 | | 0 |
| chr2 | 44540926 | 44540926 | SNV | C | C:T | 1 | | 0 |
| chr2 | 44516347 | 44516347 | SNV | T | C:T | 1 | | 0 |
| chr2 | 44532056 | 44532056 | SNV | G | G:T | 2 | | 0 |
| chr2 | 44510082 | 44510082 | SNV | C | C:T | 1 | | 0 |
| chr2 | 44546905 | 44546905 | SNV | C | C:T | 1 | | 0 |
| chr2 | 44521251 | 44521251 | SNV | C | A:C | 1 | | 0 |
| chr2 | 44508601 | 44508601 | SNV | G\|R | A:G\|Q:R | 2 | | 0 |
| chr2 | 44543271 | 44543271 | SNV | C | C:T | 1 | | 0 |
| chr2 | 44505088 | 44505088 | SNV | T | C:T | 1 | | 0 |
| chr2 | 44527730 | 44527730 | SNV | T | C:T | 3 | | 0 |
| chr2 | 44531414 | 44531414 | SNV | C\|S | C:T\|S:S | 2 | | 0 |
| chr2 | 44532284 | 44532284 | insertion | - | -:C | 4 | | 0 |
| chr2 | 44514206 | 44514206 | SNV | C | C:T | 1 | | 0 |
| chr2 | 44510169 | 44510169 | SNV | C | A:C | 1 | | 0 |
| chr2 | 44504588 | 44504588 | SNV | T | G:T | 8 | | 0 |
| chr2 | 44535106 | 44535106 | SNV | C | C:G | 2 | | 0 |
| chr2 | 44515787 | 44515787 | SNV | A | A:G | 1 | | 0 |
| chr2 | 44545390 | 44545390 | SNV | C | C:G | 1 | | 0 |
| chr2 | 44513106 | 44513106 | SNV | A | A:C | 2 | | 0 |
| chr2 | 44541699 | 44541699 | SNV | G | A:G | 1 | | 0 |
| chr2 | 44547092 | 44547092 | SNV | T | A:T | 1 | | 0 |
| chr2 | 44515442 | 44515442 | SNV | G | G:T | 1 | | 0 |
| chr2 | 44513989 | 44513989 | SNV | C | C:T | 1 | | 0 |
| chr2 | 44528761 | 44528761 | SNV | C | C:G | 1 | | 0 |
| chr2 | 44536016 | 44536016 | SNV | T | C:T | 1 | | 0 |
| chr2 | 44539970 | 44539970 | SNV | A | A:C | 2 | | 0 |
| chr2 | 44521487 | 44521487 | SNV | T | C:T | 2 | | 0 |
| chr2 | 44541327 | 44541327 | SNV | T | C:T | 3 | | 0 |
| chr2 | 44543394 | 44543394 | SNV | C | C:T | 1 | | 0 |
| chr2 | 44547673 | 44547673 | SNV | C\|N | C:T\|N:N | 1 | | 0 |
| chr2 | 44536300 | 44536300 | SNV | T | G:T | 1 | | 0 |
| chr2 | 44520530 | 44520530 | SNV | C | A:C | 1 | | 0 |
| chr2 | 44512835 | 44512835 | SNV | C | C:G | 2 | | 0 |
| chr2 | 44537841 | 44537841 | SNV | T | A:T | 1 | | 0 |
| chr2 | 44528661 | 44528661 | SNV | C | C:T | 1 | | 0 |
| chr2 | 44509355 | 44509355 | SNV | C | C:G | 1 | | 0 |
| chr2 | 44522908 | 44522908 | SNV | A | A:C | 1 | | 0 |
| chr2 | 44525008 | 44525008 | SNV | A | A:G | 1 | | 0 |
| chr2 | 44511541 | 44511541 | SNV | T | C:T | 1 | | 0 |
| chr2 | 44506939 | 44506939 | SNV | C | A:C | 1 | | 0 |
| chr2 | 44530736 | 44530736 | SNV | A | A:T | 1 | | 0 |
| chr2 | 44508585 | 44508585 | SNV | C\|Q | C:T\|Q:* | 1 | | 0 |
| chr2 | 44524862 | 44524862 | SNV | G | A:G | 1 | | 0 |
| chr2 | 44527855 | 44527855 | SNV | A | A:T | 1 | | 0 |

**Supplementary Table 1. Variants of SLC3A1 in the 1KG database**

| Chr | Start Position | End Position | Variant type | Ref Genotype \| Ref Proteotype | Variant Genotype 1\| Variant Proteotype 1 | Count | Variant Genotype 2\| Variant Proteotype 2 | Count |
|---|---|---|---|---|---|---|---|---|
| **chr2** | 44538246 | 44538246 | SNV | T | G:T | 7 | | 0 |
| **chr2** | 44522582 | 44522582 | SNV | G | C:G | 3 | | 0 |
| **chr2** | 44539086 | 44539086 | SNV | C | C:T | 1 | | 0 |
| **chr2** | 44544659 | 44544659 | SNV | C | C:T | 1 | | 0 |
| **chr2** | 44526890 | 44526890 | SNV | T | A:T | 1 | | 0 |
| **chr2** | 44507747 | 44507747 | SNV | A | A:G | 1 | | 0 |
| **chr2** | 44524330 | 44524330 | SNV | C | C:T | 1 | | 0 |
| **chr2** | 44534008 | 44534008 | SNV | T | C:T | 1 | | 0 |
| **chr2** | 44521009 | 44521009 | SNV | C | C:G | 1 | | 0 |
| **chr2** | 44543990 | 44543990 | SNV | C | C:T | 1 | | 0 |
| **chr2** | 44512484 | 44512484 | SNV | T | C:T | 1 | | 0 |
| **chr2** | 44529961 | 44529961 | SNV | A | A:C | 6 | | 0 |
| **chr2** | 44533284 | 44533284 | SNV | A | A:T | 1 | | 0 |
| **chr2** | 44529373 | 44529373 | SNV | C | C:T | 2 | | 0 |
| **chr2** | 44543783 | 44543783 | SNV | A | A:G | 1 | | 0 |
| **chr2** | 44518106 | 44518106 | SNV | T | G:T | 1 | | 0 |
| **chr2** | 44503135 | 44503135 | SNV | G | G:T | 1 | | 0 |
| **chr2** | 44529827 | 44529827 | SNV | C | A:C | 1 | | 0 |
| **chr2** | 44505712 | 44505712 | SNV | C | C:T | 1 | | 0 |
| **chr2** | 44527119 | 44527119 | SNV | C\|R | C:T\|R:W | 1 | | 0 |
| **chr2** | 44508259 | 44508259 | SNV | C | C:G | 1 | | 0 |
| **chr2** | 44509734 | 44509734 | SNV | A | A:C | 1 | | 0 |
| **chr2** | 44532152 | 44532152 | SNV | T | C:T | 1 | | 0 |
| **chr2** | 44542362 | 44542362 | SNV | G | G:T | 3 | | 0 |
| **chr2** | 44524051 | 44524051 | SNV | C | C:T | 1 | | 0 |
| **chr2** | 44505448 | 44505448 | SNV | G | A:G | 1 | | 0 |
| **chr2** | 44533344 | 44533344 | SNV | T | C:T | 2 | | 0 |
| **chr2** | 44519340 | 44519340 | SNV | C | C:T | 2 | | 0 |
| **chr2** | 44506291 | 44506291 | SNV | T | C:T | 1 | | 0 |
| **chr2** | 44545466 | 44545466 | SNV | G | A:G | 1 | | 0 |
| **chr2** | 44533895 | 44533895 | SNV | C | C:T | 1 | | 0 |
| **chr2** | 44530724 | 44530724 | SNV | T | A:T | 2 | | 0 |
| **chr2** | 44516457 | 44516457 | SNV | C | C:T | 1 | | 0 |
| **chr2** | 44530966 | 44530966 | SNV | G | A:G | 1 | | 0 |
| **chr2** | 44525113 | 44525113 | SNV | C | C:T | 1 | | 0 |
| **chr2** | 44537753 | 44537753 | SNV | A | A:G | 1 | | 0 |
| **chr2** | 44524739 | 44524739 | SNV | C | C:T | 1 | | 0 |
| **chr2** | 44538441 | 44538441 | SNV | C | A:C | 1 | | 0 |
| **chr2** | 44533226 | 44533228 | deletion | TGT | -:TGT | 4 | | 0 |
| **chr2** | 44534623 | 44534623 | SNV | C | C:T | 1 | | 0 |
| **chr2** | 44507968 | 44507968 | SNV | G\|E | A:G\|K:E | 2 | | 0 |
| **chr2** | 44536251 | 44536251 | SNV | G | A:G | 1 | | 0 |
| **chr2** | 44531804 | 44531804 | SNV | G | A:G | 1 | | 0 |
| **chr2** | 44536397 | 44536397 | SNV | C | C:T | 1 | | 0 |
| **chr2** | 44539746 | 44539746 | SNV | C\|R | C:T\|R:W | 1 | | 0 |
| **chr2** | 44526579 | 44526579 | SNV | G | C:G | 1 | | 0 |
| **chr2** | 44514016 | 44514016 | SNV | G | A:G | 1 | | 0 |
| **chr2** | 44538063 | 44538063 | SNV | A | A:G | 1 | | 0 |
| **chr2** | 44543173 | 44543173 | SNV | A | A:G | 1 | | 0 |
| **chr2** | 44531496 | 44531496 | SNV | A | A:G | 1 | | 0 |
| **chr2** | 44539110 | 44539110 | SNV | G | C:G | 1 | | 0 |
| **chr2** | 44506513 | 44506513 | SNV | C | C:T | 2 | | 0 |
| **chr2** | 44547694 | 44547694 | SNV | C\|R | A:C\|R:R | 1 | | 0 |
| **chr2** | 44520345 | 44520345 | SNV | G | G:T | 1 | | 0 |
| **chr2** | 44526618 | 44526618 | SNV | C | A:C | 2 | | 0 |
| **chr2** | 44519574 | 44519574 | SNV | G | A:G | 1 | | 0 |
| **chr2** | 44543391 | 44543391 | SNV | T | C:T | 1 | | 0 |
| **chr2** | 44523462 | 44523462 | SNV | A | A:C | 3 | | 0 |
| **chr2** | 44513917 | 44513917 | SNV | C | C:T | 1 | | 0 |
| **chr2** | 44545557 | 44545557 | SNV | C | A:C | 1 | | 0 |
| **chr2** | 44532504 | 44532504 | SNV | T | C:T | 1 | | 0 |
| **chr2** | 44509983 | 44509983 | SNV | G | A:G | 1 | | 0 |
| **chr2** | 44502979 | 44502979 | SNV | C\|A | C:T\|A:V | 2 | | 0 |
| **chr2** | 44507992 | 44507992 | SNV | A\|M | A:G\|M:V | 1 | | 0 |
| **chr2** | 44538339 | 44538339 | SNV | C | C:T | 1 | | 0 |
| **chr2** | 44504996 | 44504996 | SNV | A | A:C | 1 | | 0 |
| **chr2** | 44527532 | 44527532 | SNV | A | A:C | 2 | | 0 |
| **chr2** | 44540730 | 44540730 | SNV | G | A:G | 1 | | 0 |
| **chr2** | 44528454 | 44528454 | SNV | T | G:T | 1 | | 0 |
| **chr2** | 44514743 | 44514743 | SNV | C | C:T | 2 | | 0 |
| **chr2** | 44530166 | 44530166 | SNV | T | C:T | 1 | | 0 |
| **chr2** | 44509913 | 44509913 | SNV | A | A:T | 1 | | 0 |
| **chr2** | 44522637 | 44522637 | SNV | A | A:G | 1 | | 0 |
| **chr2** | 44533919 | 44533919 | SNV | C | C:T | 2 | | 0 |
| **chr2** | 44547488 | 44547488 | SNV | G\|D | A:G\|N:D | 2 | | 0 |
| **chr2** | 44518308 | 44518308 | SNV | C | C:G | 2 | | 0 |
| **chr2** | 44534956 | 44534956 | SNV | A | A:G | 2 | | 0 |
| **chr2** | 44522886 | 44522886 | SNV | T | A:T | 2 | | 0 |
| **chr2** | 44526785 | 44526785 | SNV | C | A:C | 1 | | 0 |
| **chr2** | 44503833 | 44503833 | SNV | G | A:G | 1 | | 0 |
| **chr2** | 44519250 | 44519250 | SNV | T | C:T | 1 | | 0 |
| **chr2** | 44521477 | 44521477 | SNV | C | G:G | 1 | C:G | 2 |
| **chr2** | 44521213 | 44521213 | SNV | A | A:G | 1 | | 0 |
| **chr2** | 44506469 | 44506469 | SNV | A | A:G | 1 | | 0 |
| **chr2** | 44533391 | 44533391 | SNV | C | A:C | 4 | | 0 |
| **chr2** | 44506692 | 44506692 | SNV | C | C:T | 1 | | 0 |
| **chr2** | 44528680 | 44528680 | SNV | C | A:C | 1 | | 0 |
| **chr2** | 44533825 | 44533825 | SNV | T | C:T | 1 | | 0 |
| **chr2** | 44538248 | 44538248 | SNV | C | C:G | 1 | | 0 |
| **chr2** | 44547560 | 44547560 | SNV | C\|L | A:C\|I:L | 1 | | 0 |
| **chr2** | 44545851 | 44545851 | SNV | C | C:T | 1 | | 0 |
| **chr2** | 44535585 | 44535585 | SNV | G | C:G | 1 | | 0 |
| **chr2** | 44543698 | 44543698 | SNV | C | C:T | 3 | | 0 |
| **chr2** | 44533493 | 44533493 | SNV | C | C:T | 1 | | 0 |

**Supplementary Table 1. Variants of SLC3A1 in the 1KG database**

| Chr | Start Position | End Position | Variant type | Ref Genotype \| Ref Proteotype | Variant Genotype 1\| Variant Proteotype 1 | Count | Variant Genotype 2\| Variant Proteotype 2 | Count |
|---|---|---|---|---|---|---|---|---|
| **chr2** | 44542787 | 44542787 | SNV | C | C:T | 1 | | 0 |
| **chr2** | 44510617 | 44510617 | SNV | G | A:G | 1 | | 0 |
| **chr2** | 44511631 | 44511631 | SNV | C | C:T | 1 | | 0 |
| **chr2** | 44540400 | 44540400 | SNV | C | C:T | 1 | | 0 |
| **chr2** | 44515819 | 44515819 | SNV | A | A:G | 1 | | 0 |
| **chr2** | 44547762 | 44547762 | SNV | T\|L | C:T\|P:L | 1 | | 0 |
| **chr2** | 44541558 | 44541558 | SNV | T | C:T | 1 | | 0 |
| **chr2** | 44503057 | 44503057 | SNV | C\|P | C:T\|P:L | 1 | | 0 |
| **chr2** | 44531198 | 44531198 | SNV | G | C:G | 1 | | 0 |
| **chr2** | 44528224 | 44528224 | SNV | G\|R | A:G\|Q:R | 1 | | 0 |
| **chr2** | 44546510 | 44546510 | SNV | T | C:T | 1 | | 0 |
| **chr2** | 44503469 | 44503469 | SNV | G | A:G | 1 | | 0 |
| **chr2** | 44506261 | 44506261 | SNV | C | C:T | 1 | | 0 |
| **chr2** | 44534654 | 44534654 | SNV | C | C:T | 1 | | 0 |
| **chr2** | 44505234 | 44505234 | SNV | C | C:T | 2 | | 0 |
| **chr2** | 44518685 | 44518685 | SNV | T | G:T | 1 | | 0 |
| **chr2** | 44505528 | 44505528 | SNV | A | A:G | 2 | | 0 |
| **chr2** | 44534771 | 44534771 | SNV | G | G:T | 1 | | 0 |
| **chr2** | 44519299 | 44519299 | SNV | G | A:G | 1 | | 0 |
| **chr2** | 44517786 | 44517786 | SNV | G | A:G | 1 | | 0 |
| **chr2** | 44532027 | 44532027 | SNV | C | C:T | 1 | | 0 |
| **chr2** | 44540459 | 44540459 | SNV | G | C:G | 2 | | 0 |
| **chr2** | 44530442 | 44530442 | SNV | G | A:G | 1 | | 0 |
| **chr2** | 44531141 | 44531141 | SNV | T | C:T | 1 | | 0 |
| **chr2** | 44539757 | 44539757 | SNV | G\|S | A:G\|S:S | 1 | | 0 |
| **chr2** | 44540074 | 44540074 | SNV | T | C:T | 1 | | 0 |
| **chr2** | 44544935 | 44544935 | SNV | A | A:T | 1 | | 0 |
| **chr2** | 44540618 | 44540618 | SNV | G | G:T | 1 | | 0 |
| **chr2** | 44504076 | 44504076 | SNV | A | A:G | 2 | | 0 |
| **chr2** | 44504757 | 44504757 | SNV | T | C:T | 1 | | 0 |
| **chr2** | 44526287 | 44526287 | SNV | C | C:T | 2 | | 0 |
| **chr2** | 44528519 | 44528519 | SNV | T | G:T | 1 | | 0 |
| **chr2** | 44531294 | 44531294 | SNV | T\|T | C:T\|T:T | 4 | | 0 |
| **chr2** | 44510041 | 44510041 | SNV | C | C:T | 1 | | 0 |
| **chr2** | 44531085 | 44531085 | SNV | T | C:T | 1 | | 0 |
| **chr2** | 44515269 | 44515269 | SNV | C | C:T | 1 | | 0 |
| **chr2** | 44514677 | 44514677 | SNV | A | A:T | 1 | | 0 |
| **chr2** | 44524895 | 44524895 | SNV | C | C:T | 1 | | 0 |
| **chr2** | 44504874 | 44504874 | SNV | T | G:T | 2 | | 0 |
| **chr2** | 44542683 | 44542683 | SNV | T | G:T | 1 | | 0 |
| **chr2** | 44528326 | 44528326 | SNV | C | A:C | 1 | | 0 |
| **chr2** | 44539443 | 44539443 | SNV | T | C:T | 4 | | 0 |
| **chr2** | 44505548 | 44505548 | SNV | C | C:T | 1 | | 0 |
| **chr2** | 44507188 | 44507188 | SNV | C | A:C | 1 | | 0 |
| **chr2** | 44509979 | 44509979 | SNV | T | A:T | 1 | | 0 |
| **chr2** | 44517062 | 44517062 | SNV | G | A:G | 2 | | 0 |
| **chr2** | 44517699 | 44517699 | SNV | G | C:G | 1 | | 0 |
| **chr2** | 44534940 | 44534940 | SNV | G | A:G | 1 | | 0 |
| **chr2** | 44526121 | 44526121 | SNV | T | G:T | 2 | | 0 |
| **chr2** | 44544359 | 44544359 | SNV | T | G:T | 1 | | 0 |
| **chr2** | 44543237 | 44543237 | SNV | C | C:T | 1 | | 0 |
| **chr2** | 44524061 | 44524061 | SNV | C | C:T | 5 | | 0 |
| **chr2** | 44509019 | 44509019 | SNV | T | A:T | 1 | | 0 |
| **chr2** | 44543622 | 44543622 | SNV | G | A:G | 1 | | 0 |
| **chr2** | 44544496 | 44544496 | SNV | G | A:G | 1 | | 0 |
| **chr2** | 44517472 | 44517472 | SNV | C | C:T | 1 | | 0 |
| **chr2** | 44543268 | 44543268 | SNV | A | A:C | 1 | | 0 |
| **chr2** | 44502747 | 44502747 | SNV | G\|V | C:G\|L:V | 1 | | 0 |
| **chr2** | 44544466 | 44544466 | SNV | T | G:T | 1 | | 0 |
| **chr2** | 44542518 | 44542518 | SNV | T | C:T | 1 | | 0 |
| **chr2** | 44533349 | 44533349 | SNV | G | A:G | 1 | | 0 |
| **chr2** | 44511229 | 44511229 | SNV | T | A:T | 1 | | 0 |
| **chr2** | 44531316 | 44531316 | SNV | G\|D | G:T\|D:Y | 1 | | 0 |
| **chr2** | 44528215 | 44528215 | SNV | G\|R | A:G\|H:R | 2 | | 0 |
| **chr2** | 44511270 | 44511270 | SNV | C | A:C | 3 | | 0 |
| **chr2** | 44547693 | 44547693 | SNV | G\|R | A:G\|H:R | 1 | | 0 |
| **chr2** | 44527476 | 44527476 | SNV | A | A:T | 1 | | 0 |
| **chr2** | 44526846 | 44526846 | SNV | T | A:T | 1 | | 0 |
| **chr2** | 44531090 | 44531090 | SNV | G | G:T | 1 | | 0 |
| **chr2** | 44519815 | 44519815 | SNV | A | A:T | 1 | | 0 |
| **chr2** | 44523738 | 44523738 | insertion | - | -:A | 3 | | 0 |
| **chr2** | 44514411 | 44514411 | SNV | T | C:T | 2 | | 0 |
| **chr2** | 44542387 | 44542387 | SNV | T | C:T | 1 | | 0 |
| **chr2** | 44525220 | 44525220 | SNV | C | C:G | 1 | | 0 |
| **chr2** | 44530232 | 44530232 | SNV | C | C:G | 2 | | 0 |
| **chr2** | 44533347 | 44533347 | SNV | G | A:G | 1 | | 0 |
| **chr2** | 44507350 | 44507350 | SNV | G | A:G | 1 | | 0 |
| **chr2** | 44525604 | 44525604 | SNV | G | A:G | 1 | | 0 |
| **chr2** | 44520922 | 44520922 | SNV | C | C:T | 2 | | 0 |
| **chr2** | 44540417 | 44540417 | SNV | G | C:G | 1 | | 0 |
| **chr2** | 44515941 | 44515941 | SNV | A | A:G | 1 | | 0 |
| **chr2** | 44546868 | 44546868 | SNV | C | C:T | 1 | | 0 |
| **chr2** | 44546362 | 44546362 | SNV | C | C:T | 1 | | 0 |
| **chr2** | 44545265 | 44545265 | SNV | C | A:C | 2 | | 0 |
| **chr2** | 44512540 | 44512540 | SNV | C | C:T | 1 | | 0 |
| **chr2** | 44541014 | 44541014 | SNV | G\|S | C:G\|T:S | 1 | | 0 |
| **chr2** | 44530972 | 44530972 | SNV | T | C:T | 1 | | 0 |
