## supplementary table 2 for "Population Genetics Analysis of *SLC3A1* and *SLC7A9* Revealed the Etiology of Cystine Stone May Be More Than What Our Current Genetic Knowledge Can Explain"

**Supplementary Table 2. Variants of SLC7A9 in the 1KG database**

| Chr | Start Position | End Position | Variant type | Ref Genotype \| Ref Proteotype | Variant Genotype 1\| Variant Proteotype 1 | Count | Variant Genotype 2\| Variant Proteotype 2 | Count |
|---|---|---|---|---|---|---|---|---|
| **chr19** | 33321444 | 33321444 | SNV | G | A:G | 4 | | 0 |
| **chr19** | 33321447 | 33321447 | SNV | A | G:G | 87 | A:G | 702 |
| **chr19** | 33321546 | 33321546 | SNV | G\|P | A:G\|S:P | 15 | | 0 |
| **chr19** | 33321587 | 33321587 | SNV | G\|P | A:G\|L:P | 21 | | 0 |
| **chr19** | 33321716 | 33321716 | SNV | C | C:T | 2 | | 0 |
| **chr19** | 33321717 | 33321717 | SNV | A | G:G | 347 | A:G | 1050 |
| **chr19** | 33321775 | 33321775 | SNV | C | G:G | 24 | C:G | 338 |
| **chr19** | 33321821 | 33321821 | SNV | G | A:G | 5 | | 0 |
| **chr19** | 33321926 | 33321926 | SNV | G | A:G | 13 | | 0 |
| **chr19** | 33321997 | 33321998 | deletion | GT | -:GT | 7 | | 0 |
| **chr19** | 33321997 | 33321997 | SNV | G | C:C | 807 | C:G | 1127 |
| **chr19** | 33322086 | 33322086 | SNV | C | T:T | 1 | C:T | 54 |
| **chr19** | 33322094 | 33322094 | SNV | A | A:G | 50 | | 0 |
| **chr19** | 33322269 | 33322269 | SNV | C | C:T | 1063 | T:T | 1085 |
| **chr19** | 33322280 | 33322280 | SNV | C | C:T | 28 | | 0 |
| **chr19** | 33322341 | 33322341 | SNV | C | T:T | 22 | C:T | 327 |
| **chr19** | 33322359 | 33322359 | insertion | - | -:T | 343 | T:T | 28 |
| **chr19** | 33322414 | 33322414 | SNV | C | A:C | 898 | A:A | 168 |
| **chr19** | 33322431 | 33322431 | SNV | G | A:A | 24 | A:G | 365 |
| **chr19** | 33322484 | 33322484 | SNV | G | A:G | 30 | | 0 |
| **chr19** | 33322496 | 33322496 | SNV | A | A:G | 29 | | 0 |
| **chr19** | 33322678 | 33322678 | deletion | C | -:C | 56 | | 0 |
| **chr19** | 33322734 | 33322734 | SNV | G | A:G | 3 | | 0 |
| **chr19** | 33322766 | 33322766 | SNV | A | A:G | 1 | | 0 |
| **chr19** | 33322907 | 33322907 | SNV | C | C:T | 74 | T:T | 2 |
| **chr19** | 33322945 | 33322945 | SNV | A | G:G | 1139 | A:G | 1020 |
| **chr19** | 33323010 | 33323010 | SNV | C | C:T | 51 | | 0 |
| **chr19** | 33323030 | 33323032 | deletion | AAC | -:AAC | 55 | -:- | 1 |
| **chr19** | 33323042 | 33323042 | SNV | G | A:A | 1139 | A:G | 1020 |
| **chr19** | 33323067 | 33323067 | SNV | C | C:T | 4 | | 0 |
| **chr19** | 33323103 | 33323103 | SNV | G | A:G | 389 | A:A | 26 |
| **chr19** | 33323214 | 33323214 | SNV | C | C:T | 22 | | 0 |
| **chr19** | 33323215 | 33323215 | SNV | G | A:G | 29 | | 0 |
| **chr19** | 33323235 | 33323235 | SNV | G | A:G | 41 | | 0 |
| **chr19** | 33323290 | 33323290 | SNV | G | G:T | 55 | | 0 |
| **chr19** | 33323349 | 33323349 | SNV | T | C:T | 539 | C:C | 51 |
| **chr19** | 33323386 | 33323386 | SNV | G | A:G | 29 | | 0 |
| **chr19** | 33323437 | 33323437 | SNV | T | A:T | 70 | A:A | 2 |
| **chr19** | 33323511 | 33323511 | deletion | G | -:G | 981 | -:- | 280 |
| **chr19** | 33323535 | 33323535 | SNV | C | C:T | 4 | | 0 |
| **chr19** | 33323661 | 33323661 | insertion | - | A:A | 25 | -:A | 386 |
| **chr19** | 33323846 | 33323846 | SNV | C | C:T | 1 | | 0 |
| **chr19** | 33323980 | 33323980 | SNV | G | T:T | 1 | G:T | 54 |
| **chr19** | 33324089 | 33324089 | SNV | G\|V | A:A\|V:V | 1 | A:G\|V:V | 71 |
| **chr19** | 33324275 | 33324275 | SNV | T | C:C | 7 | C:T | 121 |
| **chr19** | 33324711 | 33324711 | deletion | A | -:A | 1361 | -:- | 557 |
| **chr19** | 33324793 | 33324793 | insertion | - | -:T | 16 | | 0 |
| **chr19** | 33325006 | 33325006 | SNV | C | C:T | 682 | T:T | 79 |
| **chr19** | 33325045 | 33325045 | SNV | C | C:T | 3 | | 0 |
| **chr19** | 33325397 | 33325397 | SNV | G | A:G | 8 | | 0 |
| **chr19** | 33325451 | 33325451 | SNV | G | G:T | 31 | | 0 |
| **chr19** | 33325484 | 33325484 | SNV | A | C:C | 870 | A:C | 1091 |
| **chr19** | 33325530 | 33325530 | insertion | - | A:A | 1 | -:A | 23 |
| **chr19** | 33325586 | 33325586 | SNV | A | A:C | 2 | | 0 |
| **chr19** | 33325595 | 33325595 | SNV | A | A:G | 12 | | 0 |
| **chr19** | 33325619 | 33325619 | SNV | G | A:G | 3 | | 0 |
| **chr19** | 33325682 | 33325682 | SNV | T | C:T | 1 | | 0 |
| **chr19** | 33325720 | 33325720 | SNV | A | A:G | 4 | | 0 |
| **chr19** | 33325746 | 33325746 | SNV | G | A:G | 3 | | 0 |
| **chr19** | 33325841 | 33325841 | SNV | C | T:T | 7 | C:T | 119 |
| **chr19** | 33325888 | 33325888 | SNV | G | G:T | 1 | | 0 |
| **chr19** | 33325943 | 33325943 | SNV | A | A:G | 1091 | G:G | 870 |
| **chr19** | 33326333 | 33326333 | SNV | A | A:G | 3 | | 0 |
| **chr19** | 33326462 | 33326462 | SNV | G | A:G | 1 | | 0 |
| **chr19** | 33326604 | 33326604 | SNV | G | A:G | 1 | | 0 |
| **chr19** | 33326668 | 33326668 | SNV | C | C:G | 29 | | 0 |
| **chr19** | 33326679 | 33326679 | SNV | G | C:G | 667 | C:C | 77 |
| **chr19** | 33326779 | 33326779 | SNV | T | C:T | 29 | | 0 |
| **chr19** | 33326818 | 33326819 | deletion | TG | -:- | 544 | -:TG | 1260 |
| **chr19** | 33326819 | 33326819 | SNV | G | A:G | 56 | A:A | 3 |
| **chr19** | 33326889 | 33326889 | SNV | C | C:T | 13 | | 0 |
| **chr19** | 33326981 | 33326981 | SNV | G | A:G | 6 | A:A | 1 |
| **chr19** | 33327012 | 33327012 | SNV | G | C:G | 36 | | 0 |
| **chr19** | 33327082 | 33327082 | SNV | A | A:G | 1129 | G:G | 810 |
| **chr19** | 33327196 | 33327196 | SNV | G | A:G | 1129 | A:A | 810 |
| **chr19** | 33327202 | 33327202 | SNV | G | A:G | 337 | A:A | 27 |
| **chr19** | 33327474 | 33327474 | SNV | C | T:T | 1 | C:T | 33 |
| **chr19** | 33327910 | 33327910 | SNV | C | C:T | 4 | | 0 |
| **chr19** | 33327961 | 33327961 | deletion | A | -:A | 327 | -:- | 22 |
| **chr19** | 33328221 | 33328221 | SNV | C | C:T | 2 | | 0 |
| **chr19** | 33328390 | 33328390 | SNV | T | C:C | 815 | C:T | 1122 |
| **chr19** | 33328397 | 33328422 | deletion | TCAAAAAAAAAAAAAAAAAAAAAAAA | -:- | 547 | -:TCAAAAAAAAAAAAAAAAAAAAAAAA | 850 |
| **chr19** | 33328534 | 33328537 | deletion | CAAA | -:CAAA | 5 | | 0 |
| **chr19** | 33328574 | 33328574 | SNV | T | C:T | 54 | | 0 |
| **chr19** | 33328583 | 33328583 | SNV | A | G:G | 77 | A:G | 668 |
| **chr19** | 33328606 | 33328606 | SNV | C | C:T | 37 | | 0 |
| **chr19** | 33328812 | 33328812 | SNV | T | C:T | 38 | C:C | 1 |
| **chr19** | 33328989 | 33328989 | SNV | C | A:C | 49 | | 0 |
| **chr19** | 33329087 | 33329087 | SNV | C | C:T | 2 | | 0 |
| **chr19** | 33329100 | 33329100 | SNV | C | C:T | 28 | | 0 |
| **chr19** | 33329144 | 33329144 | SNV | T | C:C | 22 | C:T | 327 |
| **chr19** | 33329192 | 33329192 | SNV | A | A:G | 29 | | 0 |
| **chr19** | 33329435 | 33329435 | insertion | - | -:TATAT | 378 | TATAT:TATAT | 20 |
| **chr19** | 33329449 | 33329449 | SNV | T | C:C | 26 | C:T | 371 |
| **chr19** | 33329451 | 33329451 | SNV | T | A:T | 1 | | 0 |
| **chr19** | 33329522 | 33329529 | deletion | TATATACT | -:TATATACT | 27 | | 0 |
| **chr19** | 33329572 | 33329572 | insertion | - | -:TAG | 3 | | 0 |
| **chr19** | 33329575 | 33329575 | SNV | G | T:T | 8 | G:T | 120 |
| **chr19** | 33329595 | 33329600 | deletion | TATATG | -:- | 77 | -:TATATG | 665 |
| **chr19** | 33329612 | 33329612 | insertion | - | TA:TA | 22 | -:TA | 379 |
| **chr19** | 33329639 | 33329640 | deletion | AC | -:AC | 29 | | 0 |
| **chr19** | 33329712 | 33329715 | deletion | TATC | -:TATC | 327 | -:- | 22 |
| **chr19** | 33329770 | 33329770 | SNV | A | G:G | 22 | A:G | 327 |
| **chr19** | 33330009 | 33330009 | SNV | G | A:G | 58 | | 0 |
| **chr19** | 33330079 | 33330079 | SNV | A | A:G | 22 | | 0 |
| **chr19** | 33330171 | 33330171 | SNV | A | A:G | 24 | | 0 |
| **chr19** | 33330209 | 33330209 | SNV | C | C:G | 29 | | 0 |
| **chr19** | 33330320 | 33330320 | SNV | G | A:G | 51 | | 0 |
| **chr19** | 33330462 | 33330462 | SNV | A | A:C | 29 | | 0 |
| **chr19** | 33330575 | 33330575 | SNV | C | C:T | 7 | | 0 |
| **chr19** | 33330666 | 33330666 | SNV | C | G:G | 1089 | C:G | 1058 |
| **chr19** | 33330917 | 33330917 | SNV | C | T:T | 787 | C:T | 1139 |
| **chr19** | 33330974 | 33330974 | SNV | T | C:T | 2 | | 0 |
| **chr19** | 33331102 | 33331102 | SNV | G | A:G | 1 | | 0 |
| **chr19** | 33331218 | 33331218 | SNV | A | G:G | 8 | A:G | 135 |
| **chr19** | 33331458 | 33331468 | deletion | TGGCCTCTCTG | -:TGGCCTCTCTG | 56 | | 0 |
| **chr19** | 33331529 | 33331529 | SNV | T | C:T | 33 | | 0 |
| **chr19** | 33331544 | 33331544 | SNV | A | G:G | 25 | A:G | 388 |
| **chr19** | 33331573 | 33331573 | SNV | C | C:G | 1 | | 0 |
| **chr19** | 33331598 | 33331598 | SNV | A | A:G | 18 | | 0 |
| **chr19** | 33331618 | 33331618 | SNV | T | C:T | 1 | | 0 |
| **chr19** | 33331722 | 33331722 | SNV | G | A:G | 2 | | 0 |
| **chr19** | 33331738 | 33331738 | SNV | G | A:G | 1 | | 0 |
| **chr19** | 33331910 | 33331910 | SNV | T | C:T | 37 | C:C | 1 |
| **chr19** | 33331913 | 33331913 | SNV | C | T:T | 516 | C:T | 1176 |
| **chr19** | 33332189 | 33332189 | SNV | A | A:G | 1 | | 0 |
| **chr19** | 33332312 | 33332312 | SNV | C | C:G | 334 | G:G | 22 |
| **chr19** | 33332516 | 33332516 | SNV | G | A:G | 377 | A:A | 22 |
| **chr19** | 33332642 | 33332642 | SNV | A | G:G | 22 | A:G | 376 |
| **chr19** | 33332737 | 33332737 | SNV | G | A:G | 5 | | 0 |
| **chr19** | 33332791 | 33332791 | SNV | C | T:T | 1 | C:T | 33 |
| **chr19** | 33332828 | 33332828 | SNV | C | G:G | 1082 | C:G | 1060 |

**Supplementary Table 2. Variants of SLC7A9 in the 1KG database**

| Chr | Start Position | End Position | Variant type | Ref Genotype \| Ref Proteotype | Variant Genotype 1\| Variant Proteotype 1 | Count | Variant Genotype 2\| Variant Proteotype 2 | Count |
|---|---|---|---|---|---|---|---|---|
| chr19 | 33332920 | 33332920 | SNV | A | A:C | 5 | | 0 |
| chr19 | 33332927 | 33332927 | SNV | G | A:G | 12 | | 0 |
| chr19 | 33332952 | 33332952 | SNV | C | C:T | 695 | T:T | 82 |
| chr19 | 33333096 | 33333096 | SNV | T\|K | C:T\|R:K | 1 | | 0 |
| chr19 | 33333155 | 33333155 | SNV | G\|A | A:G\|A:A | 911 | A:A\|A:A | 166 |
| chr19 | 33333432 | 33333432 | SNV | A | A:C | 13 | | 0 |
| chr19 | 33333480 | 33333480 | SNV | T | A:T | 1 | | 0 |
| chr19 | 33333680 | 33333680 | SNV | G | A:G | 60 | A:A | 2 |
| chr19 | 33333744 | 33333744 | SNV | C | C:T | 1 | | 0 |
| chr19 | 33333745 | 33333745 | SNV | G | A:G | 10 | | 0 |
| chr19 | 33334025 | 33334025 | SNV | C | C:T | 328 | T:T | 22 |
| chr19 | 33334161 | 33334161 | SNV | A | G:G | 1088 | A:G | 1057 |
| chr19 | 33334207 | 33334207 | deletion | A | -:A | 3 | | 0 |
| chr19 | 33334255 | 33334255 | SNV | C | C:T | 23 | | 0 |
| chr19 | 33334311 | 33334311 | SNV | C | C:T | 3 | | 0 |
| chr19 | 33334362 | 33334362 | SNV | G | A:G | 30 | | 0 |
| chr19 | 33334366 | 33334366 | insertion | - | A:A | 7 | -:A | 136 |
| chr19 | 33334394 | 33334395 | deletion | CA | -:- | 2 | -:CA | 90 |
| chr19 | 33334415 | 33334415 | SNV | T | C:T | 1061 | C:C | 1080 |
| chr19 | 33334441 | 33334441 | SNV | G | A:G | 4 | | 0 |
| chr19 | 33334546 | 33334546 | SNV | A | A:G | 1 | | 0 |
| chr19 | 33334565 | 33334565 | SNV | A | C:C | 1 | A:C | 30 |
| chr19 | 33334641 | 33334641 | SNV | A | A:G | 2 | | 0 |
| chr19 | 33334693 | 33334693 | SNV | G | A:G | 241 | A:A | 24 |
| chr19 | 33334776 | 33334776 | SNV | G\|P | A:G\|P:P | 13 | | 0 |
| chr19 | 33334874 | 33334874 | SNV | C | C:T | 6 | | 0 |
| chr19 | 33334882 | 33334882 | SNV | C | T:T | 1 | C:T | 27 |
| chr19 | 33334934 | 33334934 | SNV | G | A:G | 1 | | 0 |
| chr19 | 33334978 | 33334978 | SNV | G | A:G | 788 | A:A | 176 |
| chr19 | 33335085 | 33335085 | SNV | G | A:G | 20 | | 0 |
| chr19 | 33335943 | 33335943 | SNV | G | A:A | 809 | A:G | 1125 |
| chr19 | 33335984 | 33335984 | SNV | C | C:T | 185 | T:T | 9 |
| chr19 | 33335985 | 33335985 | SNV | G | A:G | 2 | | 0 |
| chr19 | 33336011 | 33336011 | SNV | A | T:T | 1 | A:T | 33 |
| chr19 | 33336073 | 33336073 | SNV | A | A:G | 8 | | 0 |
| chr19 | 33336080 | 33336083 | deletion | AAAT | -:AAAT | 1125 | -:- | 809 |
| chr19 | 33336213 | 33336213 | SNV | C | T:T | 176 | C:T | 789 |
| chr19 | 33336230 | 33336230 | SNV | G | A:G | 328 | A:A | 22 |
| chr19 | 33336262 | 33336262 | SNV | G | A:A | 1 | A:G | 9 |
| chr19 | 33336269 | 33336269 | SNV | C | T:T | 810 | C:T | 1124 |
| chr19 | 33336270 | 33336270 | SNV | G | A:G | 14 | | 0 |
| chr19 | 33336330 | 33336330 | SNV | A | A:G | 5 | | 0 |
| chr19 | 33336337 | 33336337 | SNV | G | A:G | 1 | | 0 |
| chr19 | 33336651 | 33336651 | SNV | T | C:C | 1 | C:T | 12 |
| chr19 | 33336667 | 33336667 | SNV | A | A:G | 7 | | 0 |
| chr19 | 33336711 | 33336711 | SNV | C | C:T | 612 | T:T | 59 |
| chr19 | 33336774 | 33336774 | SNV | A | A:G | 1018 | G:G | 1134 |
| chr19 | 33336859 | 33336859 | SNV | G | A:G | 74 | A:A | 1 |
| chr19 | 33337075 | 33337075 | SNV | C | C:T | 49 | | 0 |
| chr19 | 33337081 | 33337081 | SNV | A | A:G | 1 | | 0 |
| chr19 | 33337085 | 33337085 | SNV | T | C:T | 2 | | 0 |
| chr19 | 33337100 | 33337100 | SNV | G | A:G | 16 | | 0 |
| chr19 | 33337111 | 33337111 | SNV | G | A:G | 379 | A:A | 22 |
| chr19 | 33337199 | 33337199 | deletion | C | -:- | 807 | -:C | 1127 |
| chr19 | 33337248 | 33337248 | SNV | C | A:C | 22 | | 0 |
| chr19 | 33337280 | 33337280 | SNV | C | C:T | 14 | | 0 |
| chr19 | 33337303 | 33337303 | SNV | C | C:T | 787 | T:T | 176 |
| chr19 | 33337310 | 33337310 | SNV | A | A:G | 1062 | G:G | 1079 |
| chr19 | 33337379 | 33337379 | SNV | G | A:G | 20 | A:A | 1 |
| chr19 | 33337423 | 33337423 | SNV | A | G:G | 1079 | A:G | 1062 |
| chr19 | 33337534 | 33337534 | SNV | T | C:T | 17 | | 0 |
| chr19 | 33337608 | 33337608 | SNV | G | C:C | 1 | C:G | 28 |
| chr19 | 33337669 | 33337669 | SNV | A | C:C | 22 | A:C | 377 |
| chr19 | 33337686 | 33337686 | SNV | G | C:C | 3 | C:G | 52 |
| chr19 | 33337775 | 33337775 | SNV | G | A:G | 1 | | 0 |
| chr19 | 33337861 | 33337861 | SNV | G | A:G | 377 | A:A | 22 |
| chr19 | 33337924 | 33337924 | SNV | C | C:T | 2 | | 0 |
| chr19 | 33338073 | 33338073 | SNV | T | C:T | 376 | C:C | 22 |
| chr19 | 33338083 | 33338083 | SNV | C | C:T | 49 | | 0 |
| chr19 | 33338089 | 33338089 | SNV | A | A:G | 376 | G:G | 22 |
| chr19 | 33338105 | 33338105 | SNV | G | A:A | 1 | A:G | 44 |
| chr19 | 33338118 | 33338118 | SNV | A | G:G | 22 | A:G | 375 |
| chr19 | 33338119 | 33338119 | SNV | A | A:C | 375 | C:C | 22 |
| chr19 | 33338207 | 33338207 | SNV | A | A:G | 1061 | G:G | 1080 |
| chr19 | 33338281 | 33338281 | SNV | A | A:G | 1 | | 0 |
| chr19 | 33338286 | 33338286 | SNV | G | A:G | 7 | | 0 |
| chr19 | 33338350 | 33338350 | insertion | - | -:GATGAAATG | 237 | GATGAAATG:GATGAAATG | 21 |
| chr19 | 33338372 | 33338372 | SNV | C | C:T | 50 | | 0 |
| chr19 | 33338373 | 33338373 | SNV | G | A:G | 28 | | 0 |
| chr19 | 33338566 | 33338566 | SNV | T | A:T | 11 | | 0 |
| chr19 | 33338573 | 33338573 | SNV | A | A:T | 2 | | 0 |
| chr19 | 33338605 | 33338605 | SNV | C | T:T | 82 | C:T | 694 |
| chr19 | 33338721 | 33338721 | SNV | C | C:T | 1125 | T:T | 809 |
| chr19 | 33338723 | 33338723 | SNV | G | A:A | 22 | A:G | 328 |
| chr19 | 33338784 | 33338784 | SNV | C | C:T | 58 | | 0 |
| chr19 | 33338790 | 33338790 | SNV | G | A:G | 10 | | 0 |
| chr19 | 33338827 | 33338827 | SNV | G | A:A | 1080 | A:G | 1061 |
| chr19 | 33338839 | 33338839 | SNV | G | A:G | 377 | A:A | 22 |
| chr19 | 33339247 | 33339247 | SNV | G | G:T | 1 | | 0 |
| chr19 | 33339321 | 33339321 | SNV | C | A:A | 176 | A:C | 789 |
| chr19 | 33339360 | 33339360 | deletion | A | -:- | 165 | -:A | 908 |
| chr19 | 33339427 | 33339427 | SNV | C | C:T | 191 | T:T | 17 |
| chr19 | 33339547 | 33339547 | SNV | C | C:G | 2 | | 0 |
| chr19 | 33339737 | 33339737 | SNV | T | C:T | 6 | | 0 |
| chr19 | 33339871 | 33339871 | deletion | A | -:- | 22 | -:A | 328 |
| chr19 | 33340192 | 33340192 | SNV | A | A:G | 329 | G:G | 20 |
| chr19 | 33340611 | 33340611 | SNV | A | A:C | 378 | C:C | 20 |
| chr19 | 33341116 | 33341116 | SNV | G | C:C | 166 | C:G | 901 |
| chr19 | 33341184 | 33341184 | SNV | T | A:A | 1005 | A:T | 1081 |
| chr19 | 33341485 | 33341485 | SNV | A | A:G | 57 | | 0 |
| chr19 | 33341621 | 33341621 | SNV | A | A:C | 109 | C:C | 8 |
| chr19 | 33341852 | 33341852 | SNV | G | A:G | 5 | | 0 |
| chr19 | 33342084 | 33342084 | SNV | T | C:T | 330 | C:C | 25 |
| chr19 | 33342169 | 33342169 | SNV | G | A:G | 2 | | 0 |
| chr19 | 33342403 | 33342403 | SNV | C | C:T | 1232 | T:T | 548 |
| chr19 | 33342551 | 33342551 | SNV | A | A:G | 7 | | 0 |
| chr19 | 33342584 | 33342584 | deletion | C | -:C | 4 | | 0 |
| chr19 | 33342628 | 33342628 | SNV | G | A:A | 28 | A:G | 340 |
| chr19 | 33342719 | 33342719 | SNV | C | C:G | 4 | | 0 |
| chr19 | 33342833 | 33342833 | SNV | A | A:C | 22 | | 0 |
| chr19 | 33342842 | 33342842 | SNV | C | C:T | 94 | T:T | 9 |
| chr19 | 33342844 | 33342844 | SNV | G | A:G | 705 | A:A | 82 |
| chr19 | 33342916 | 33342916 | SNV | G | A:G | 332 | A:A | 22 |
| chr19 | 33342950 | 33342950 | SNV | A | A:T | 2 | | 0 |
| chr19 | 33342977 | 33342977 | SNV | A | A:T | 789 | T:T | 176 |
| chr19 | 33343178 | 33343178 | SNV | C | A:C | 15 | | 0 |
| chr19 | 33343405 | 33343405 | SNV | C | C:T | 12 | | 0 |
| chr19 | 33343599 | 33343599 | SNV | G | A:G | 2 | | 0 |
| chr19 | 33343616 | 33343616 | SNV | A | A:T | 377 | T:T | 22 |
| chr19 | 33343684 | 33343684 | SNV | G | C:G | 19 | C:C | 2 |
| chr19 | 33343722 | 33343722 | SNV | C | C:G | 1094 | G:G | 1035 |
| chr19 | 33343851 | 33343851 | SNV | G | A:G | 789 | A:A | 176 |
| chr19 | 33343955 | 33343955 | SNV | G | G:T | 1127 | T:T | 809 |
| chr19 | 33343968 | 33343968 | SNV | C | C:T | 28 | | 0 |
| chr19 | 33343977 | 33343977 | SNV | C | C:T | 15 | | 0 |
| chr19 | 33344068 | 33344068 | SNV | C | C:T | 46 | | 0 |
| chr19 | 33344221 | 33344221 | SNV | A | A:G | 1 | | 0 |
| chr19 | 33344417 | 33344417 | deletion | T | -:- | 779 | -:T | 1135 |
| chr19 | 33344545 | 33344545 | SNV | A | A:C | 2 | | 0 |
| chr19 | 33344573 | 33344573 | SNV | A | A:G | 28 | | 0 |
| chr19 | 33344823 | 33344826 | deletion | AAAA | -:AAAA | 1052 | -:- | 1105 |

**Supplementary Table 2. Variants of SLC7A9 in the 1KG database**

| Chr | Start Position | End Position | Variant type | Ref Genotype \| Ref Proteotype | Variant Genotype 1\| Variant Proteotype 1 | Count | Variant Genotype 2\| Variant Proteotype 2 | Count |
|---|---|---|---|---|---|---|---|---|
| **chr19** | 33344949 | 33344949 | SNV | T | C:T | 1 | | 0 |
| **chr19** | 33344989 | 33344989 | SNV | G | C:G | 27 | | 0 |
| **chr19** | 33345494 | 33345496 | deletion | TTG | -:TTG | 9 | | 0 |
| **chr19** | 33345563 | 33345563 | SNV | C | C:T | 12 | | 0 |
| **chr19** | 33345588 | 33345588 | SNV | G | A:G | 9 | | 0 |
| **chr19** | 33345796 | 33345796 | SNV | G | T:T | 1 | G:T | 11 |
| **chr19** | 33345805 | 33345805 | SNV | T | G:T | 1 | | 0 |
| **chr19** | 33345858 | 33345858 | insertion | - | TA:TA | 1 | -:TA | 62 |
| **chr19** | 33345945 | 33345945 | SNV | G | A:A | 797 | A:G | 1143 |
| **chr19** | 33346027 | 33346027 | SNV | G | A:G | 1 | | 0 |
| **chr19** | 33346046 | 33346046 | SNV | C | C:T | 477 | T:T | 30 |
| **chr19** | 33346135 | 33346135 | SNV | G | A:G | 1144 | A:A | 792 |
| **chr19** | 33346201 | 33346201 | SNV | T | C:T | 6 | | 0 |
| **chr19** | 33346204 | 33346204 | SNV | G | A:A | 1077 | A:G | 1077 |
| **chr19** | 33346220 | 33346220 | SNV | G | A:G | 63 | A:A | 1 |
| **chr19** | 33346331 | 33346331 | SNV | C | T:T | 6 | C:T | 164 |
| **chr19** | 33346341 | 33346341 | SNV | C | C:T | 1202 | T:T | 640 |
| **chr19** | 33346344 | 33346344 | SNV | G | A:G | 21 | | 0 |
| **chr19** | 33346354 | 33346354 | SNV | C | A:C | 6 | | 0 |
| **chr19** | 33346372 | 33346372 | SNV | A | A:G | 135 | G:G | 1 |
| **chr19** | 33346402 | 33346402 | SNV | A | G:G | 1 | A:G | 104 |
| **chr19** | 33346412 | 33346412 | SNV | T | C:C | 38 | C:T | 518 |
| **chr19** | 33346416 | 33346416 | SNV | A | A:T | 1017 | T:T | 293 |
| **chr19** | 33346421 | 33346421 | SNV | C | C:T | 3 | | 0 |
| **chr19** | 33346422 | 33346422 | SNV | G | A:G | 2 | | 0 |
| **chr19** | 33346430 | 33346430 | SNV | C | T:T | 2 | C:T | 89 |
| **chr19** | 33346433 | 33346433 | SNV | A | A:G | 415 | G:G | 17 |
| **chr19** | 33346480 | 33346480 | SNV | C | C:T | 16 | | 0 |
| **chr19** | 33346481 | 33346481 | SNV | A | A:G | 18 | | 0 |
| **chr19** | 33346525 | 33346525 | SNV | G | A:G | 14 | | 0 |
| **chr19** | 33346526 | 33346526 | insertion | - | GA:GA | 1 | -:GA | 95 |
| **chr19** | 33346544 | 33346544 | SNV | C | C:G | 1 | | 0 |
| **chr19** | 33346586 | 33346586 | SNV | T | C:T | 38 | C:C | 1 |
| **chr19** | 33346607 | 33346607 | insertion | - | -:A | 447 | A:A | 20 |
| **chr19** | 33346686 | 33346686 | SNV | C | C:T | 1 | | 0 |
| **chr19** | 33346770 | 33346770 | SNV | C | C:T | 517 | T:T | 44 |
| **chr19** | 33346799 | 33346799 | SNV | G | C:C | 37 | C:G | 526 |
| **chr19** | 33346836 | 33346836 | SNV | G | A:G | 67 | A:A | 2 |
| **chr19** | 33346883 | 33346883 | SNV | T | C:T | 470 | C:C | 30 |
| **chr19** | 33347193 | 33347193 | SNV | T | C:T | 415 | C:C | 26 |
| **chr19** | 33347246 | 33347246 | SNV | G | A:A | 21 | A:G | 318 |
| **chr19** | 33347277 | 33347277 | SNV | A | T:T | 25 | A:T | 417 |
| **chr19** | 33347291 | 33347291 | SNV | A | A:G | 1 | | 0 |
| **chr19** | 33347327 | 33347327 | SNV | C | C:T | 38 | | 0 |
| **chr19** | 33347352 | 33347352 | SNV | G | A:G | 13 | | 0 |
| **chr19** | 33347353 | 33347353 | SNV | G | C:G | 3 | | 0 |
| **chr19** | 33347390 | 33347390 | SNV | A | A:G | 22 | | 0 |
| **chr19** | 33347391 | 33347391 | SNV | T | C:T | 22 | | 0 |
| **chr19** | 33347403 | 33347403 | SNV | G | G:T | 326 | T:T | 21 |
| **chr19** | 33347439 | 33347439 | SNV | G | A:G | 33 | A:A | 1 |
| **chr19** | 33347461 | 33347463 | deletion | AAT | -:AAT | 9 | | 0 |
| **chr19** | 33347776 | 33347776 | SNV | C | C:T | 22 | | 0 |
| **chr19** | 33347781 | 33347781 | SNV | G | G:T | 20 | | 0 |
| **chr19** | 33347840 | 33347840 | SNV | C | C:T | 2 | | 0 |
| **chr19** | 33348190 | 33348190 | SNV | G | A:A | 26 | A:G | 430 |
| **chr19** | 33348191 | 33348191 | SNV | A | T:T | 2 | A:T | 63 |
| **chr19** | 33348230 | 33348230 | SNV | G | G:T | 25 | | 0 |
| **chr19** | 33348269 | 33348269 | SNV | G | C:G | 1 | | 0 |
| **chr19** | 33348390 | 33348390 | deletion | A | -:A | 927 | -:- | 190 |
| **chr19** | 33348418 | 33348418 | SNV | G | A:A | 48 | A:G | 549 |
| **chr19** | 33348615 | 33348615 | SNV | A | A:G | 1 | | 0 |
| **chr19** | 33348638 | 33348638 | SNV | G | A:G | 3 | | 0 |
| **chr19** | 33349179 | 33349179 | SNV | G | A:G | 401 | A:A | 21 |
| **chr19** | 33349274 | 33349274 | SNV | G | C:G | 2 | | 0 |
| **chr19** | 33349295 | 33349295 | SNV | T | C:C | 39 | C:T | 548 |
| **chr19** | 33349351 | 33349351 | SNV | C\|A | C:T\|A:A | 34 | | 0 |
| **chr19** | 33349381 | 33349381 | SNV | G\|I | A:G\|I:I | 1 | | 0 |
| **chr19** | 33349488 | 33349488 | SNV | T | C:C | 1530 | C:T | 785 |
| **chr19** | 33349613 | 33349613 | SNV | C | C:T | 1 | | 0 |
| **chr19** | 33349674 | 33349674 | SNV | C | A:A | 1 | A:C | 33 |
| **chr19** | 33349676 | 33349676 | SNV | T | A:T | 714 | A:A | 81 |
| **chr19** | 33349694 | 33349694 | SNV | A | A:G | 17 | | 0 |
| **chr19** | 33349715 | 33349715 | SNV | A | C:C | 2 | A:C | 83 |
| **chr19** | 33349751 | 33349751 | SNV | A | A:G | 300 | G:G | 21 |
| **chr19** | 33349764 | 33349764 | SNV | G | A:G | 715 | A:A | 77 |
| **chr19** | 33349805 | 33349805 | SNV | G | A:G | 64 | | 0 |
| **chr19** | 33349930 | 33349930 | SNV | C | C:T | 1022 | T:T | 212 |
| **chr19** | 33349967 | 33349967 | SNV | A | A:C | 78 | C:C | 1 |
| **chr19** | 33349992 | 33349992 | SNV | C | C:T | 3 | | 0 |
| **chr19** | 33350060 | 33350060 | SNV | T | C:C | 1170 | C:T | 1063 |
| **chr19** | 33350071 | 33350071 | SNV | T | C:T | 8 | | 0 |
| **chr19** | 33350101 | 33350101 | SNV | G | A:A | 176 | A:G | 959 |
| **chr19** | 33350106 | 33350106 | SNV | A | A:G | 7 | G:G | 1 |
| **chr19** | 33350249 | 33350249 | insertion | - | -:T | 26 | | 0 |
| **chr19** | 33350263 | 33350263 | SNV | C | G:G | 6 | C:G | 143 |
| **chr19** | 33350305 | 33350305 | SNV | T | C:C | 2 | C:T | 61 |
| **chr19** | 33350315 | 33350315 | SNV | G | A:G | 26 | | 0 |
| **chr19** | 33350479 | 33350479 | SNV | A | A:G | 102 | G:G | 33 |
| **chr19** | 33350610 | 33350610 | SNV | G | A:G | 2 | | 0 |
| **chr19** | 33350622 | 33350622 | SNV | T | C:T | 1 | | 0 |
| **chr19** | 33351065 | 33351065 | SNV | A | A:G | 20 | | 0 |
| **chr19** | 33351136 | 33351136 | SNV | C | C:T | 47 | T:T | 1 |
| **chr19** | 33351695 | 33351695 | SNV | T | C:T | 1161 | C:C | 312 |
| **chr19** | 33351913 | 33351913 | SNV | G | A:G | 15 | | 0 |
| **chr19** | 33351972 | 33351972 | SNV | C | C:T | 20 | | 0 |
| **chr19** | 33352041 | 33352041 | SNV | C | C:G | 28 | | 0 |
| **chr19** | 33352186 | 33352186 | SNV | C | C:T | 25 | | 0 |
| **chr19** | 33352190 | 33352190 | SNV | A | A:C | 4 | | 0 |
| **chr19** | 33352287 | 33352287 | SNV | C | C:T | 20 | | 0 |
| **chr19** | 33352449 | 33352449 | SNV | G | A:G | 17 | | 0 |
| **chr19** | 33352456 | 33352456 | SNV | G | A:G | 38 | | 0 |
| **chr19** | 33352484 | 33352484 | SNV | G | A:G | 120 | A:A | 9 |
| **chr19** | 33352654 | 33352654 | SNV | A | A:G | 1059 | G:G | 214 |
| **chr19** | 33352720 | 33352720 | insertion | - | -:AAAAAAAC | 6 | | 0 |
| **chr19** | 33352770 | 33352770 | SNV | G | A:G | 804 | A:A | 106 |
| **chr19** | 33352840 | 33352840 | SNV | G | G:T | 17 | | 0 |
| **chr19** | 33352871 | 33352871 | SNV | C | C:T | 2 | | 0 |
| **chr19** | 33352961 | 33352961 | SNV | G | A:G | 4 | | 0 |
| **chr19** | 33352969 | 33352969 | SNV | G | A:G | 1 | | 0 |
| **chr19** | 33352980 | 33352980 | SNV | A | A:G | 19 | | 0 |
| **chr19** | 33352997 | 33352997 | SNV | G | A:G | 1 | | 0 |
| **chr19** | 33353041 | 33353041 | SNV | G\|L | A:G\|L:L | 1013 | A:A\|L:L | 190 |
| **chr19** | 33353061 | 33353061 | SNV | G\|L | G:T\|L:M | 1013 | T:T\|M:M | 190 |
| **chr19** | 33353215 | 33353215 | SNV | G | A:G | 125 | A:A | 10 |
| **chr19** | 33353343 | 33353343 | SNV | A | G:G | 193 | A:G | 1017 |
| **chr19** | 33353357 | 33353357 | SNV | C | C:T | 320 | T:T | 21 |
| **chr19** | 33353427 | 33353427 | SNV | C\|A | C:T\|A:T | 5 | | 0 |
| **chr19** | 33353464 | 33353464 | SNV | G\|S | A:G\|S:S | 1016 | A:A\|S:S | 193 |
| **chr19** | 33353580 | 33353580 | SNV | T | A:T | 33 | A:A | 1 |
| **chr19** | 33353581 | 33353581 | SNV | T | A:T | 24 | | 0 |
| **chr19** | 33353582 | 33353582 | SNV | T | A:T | 2 | | 0 |
| **chr19** | 33353716 | 33353716 | SNV | G | A:G | 2 | | 0 |
| **chr19** | 33353854 | 33353854 | SNV | T | C:C | 225 | C:T | 1067 |
| **chr19** | 33353889 | 33353889 | SNV | G | G:T | 24 | | 0 |
| **chr19** | 33354189 | 33354189 | SNV | C | C:T | 316 | T:T | 21 |
| **chr19** | 33354255 | 33354255 | SNV | A | A:T | 1 | | 0 |
| **chr19** | 33354280 | 33354280 | SNV | A | A:G | 1067 | G:G | 233 |
| **chr19** | 33354401 | 33354401 | SNV | G | A:G | 315 | A:A | 21 |
| **chr19** | 33354404 | 33354404 | SNV | G | A:G | 320 | A:A | 21 |
| **chr19** | 33354576 | 33354576 | insertion | - | -:CC | 11 | | 0 |
| **chr19** | 33354676 | 33354676 | SNV | G | A:A | 1 | A:G | 76 |

**Supplementary Table 2. Variants of SLC7A9 in the 1KG database**

| Chr | Start Position | End Position | Variant type | Ref Genotype \| Ref Proteotype | Variant Genotype 1\| Variant Proteotype 1 | Count | Variant Genotype 2\| Variant Proteotype 2 | Count |
|---|---|---|---|---|---|---|---|---|
| **chr19** | 33354820 | 33354820 | SNV | C | C:T | 60 | | 0 |
| **chr19** | 33354826 | 33354826 | insertion | - | -:C | 23 | | 0 |
| **chr19** | 33354832 | 33354832 | SNV | A | G:G | 1 | A:G | 23 |
| **chr19** | 33354992 | 33354992 | SNV | A | A:G | 363 | G:G | 21 |
| **chr19** | 33355055 | 33355055 | SNV | A\|V | G:G\|A:A | 232 | A:G\|V:A | 1069 |
| **chr19** | 33355069 | 33355069 | SNV | A\|C | G:G\|C:C | 21 | A:G\|C:C | 320 |
| **chr19** | 33355081 | 33355081 | SNV | G\|S | A:A\|S:S | 117 | A:G\|S:S | 825 |
| **chr19** | 33355167 | 33355167 | SNV | C\|G | C:T\|G:R | 3 | | 0 |
| **chr19** | 33355228 | 33355228 | SNV | C\|A | C:T\|A:A | 17 | | 0 |
| **chr19** | 33355295 | 33355295 | SNV | G | A:G | 8 | A:A | 1 |
| **chr19** | 33355305 | 33355305 | SNV | A | A:T | 4 | | 0 |
| **chr19** | 33355326 | 33355326 | SNV | T | A:T | 13 | | 0 |
| **chr19** | 33355356 | 33355356 | SNV | C | C:T | 321 | T:T | 20 |
| **chr19** | 33355495 | 33355495 | SNV | C | A:C | 1 | | 0 |
| **chr19** | 33355513 | 33355513 | SNV | A | A:C | 1033 | C:C | 202 |
| **chr19** | 33355542 | 33355542 | SNV | C\|A | G:G\|A:A | 1 | C:G\|A:A | 34 |
| **chr19** | 33355772 | 33355772 | SNV | C | T:T | 1 | C:T | 37 |
| **chr19** | 33355781 | 33355781 | SNV | C | C:T | 2 | | 0 |
| **chr19** | 33355782 | 33355782 | SNV | C | C:G | 12 | | 0 |
| **chr19** | 33355816 | 33355816 | SNV | G | A:G | 13 | | 0 |
| **chr19** | 33355892 | 33355892 | SNV | G | A:A | 2 | A:G | 113 |
| **chr19** | 33355893 | 33355893 | SNV | C | C:T | 3 | | 0 |
| **chr19** | 33355951 | 33355951 | SNV | A | G:G | 185 | A:G | 1002 |
| **chr19** | 33356018 | 33356018 | SNV | C | C:G | 2 | | 0 |
| **chr19** | 33356076 | 33356076 | SNV | G | A:G | 20 | | 0 |
| **chr19** | 33356082 | 33356082 | SNV | G | A:G | 4 | | 0 |
| **chr19** | 33356088 | 33356088 | SNV | C | C:T | 89 | | 0 |
| **chr19** | 33356145 | 33356145 | SNV | T | C:T | 60 | | 0 |
| **chr19** | 33356246 | 33356246 | SNV | G | C:C | 7 | C:G | 103 |
| **chr19** | 33356287 | 33356287 | SNV | G | C:G | 42 | | 0 |
| **chr19** | 33356384 | 33356384 | SNV | T | G:T | 14 | | 0 |
| **chr19** | 33356522 | 33356522 | SNV | G | C:G | 27 | | 0 |
| **chr19** | 33356706 | 33356706 | SNV | G | A:G | 1 | | 0 |
| **chr19** | 33356857 | 33356857 | insertion | - | CT:CT | 1251 | -:CT | 1003 |
| **chr19** | 33356879 | 33356879 | SNV | A | A:C | 41 | | 0 |
| **chr19** | 33356891 | 33356891 | SNV | T | C:T | 1169 | C:C | 476 |
| **chr19** | 33356919 | 33356921 | deletion | AAG | -:AAG | 6 | | 0 |
| **chr19** | 33356962 | 33356962 | SNV | C | C:T | 405 | T:T | 30 |
| **chr19** | 33357016 | 33357016 | SNV | A | A:C | 162 | C:C | 14 |
| **chr19** | 33357039 | 33357039 | SNV | C | T:T | 1 | C:T | 34 |
| **chr19** | 33357058 | 33357058 | SNV | G | A:A | 40 | A:G | 243 |
| **chr19** | 33357099 | 33357099 | SNV | G | A:G | 315 | A:A | 21 |
| **chr19** | 33357176 | 33357176 | SNV | G | A:G | 493 | A:A | 46 |
| **chr19** | 33357187 | 33357187 | SNV | C | C:T | 508 | T:T | 59 |
| **chr19** | 33357297 | 33357297 | SNV | C | C:T | 1 | | 0 |
| **chr19** | 33357318 | 33357318 | SNV | A | G:G | 1243 | A:G | 1008 |
| **chr19** | 33357340 | 33357340 | SNV | G | A:G | 3 | | 0 |
| **chr19** | 33357433 | 33357433 | SNV | G | C:C | 704 | C:G | 1235 |
| **chr19** | 33357677 | 33357677 | SNV | T | C:C | 105 | C:T | 607 |
| **chr19** | 33357705 | 33357705 | SNV | G | A:A | 32 | A:G | 110 |
| **chr19** | 33357779 | 33357779 | SNV | G | A:G | 20 | | 0 |
| **chr19** | 33357844 | 33357844 | SNV | G | C:C | 552 | C:G | 1181 |
| **chr19** | 33358007 | 33358007 | SNV | G | A:G | 62 | A:A | 1 |
| **chr19** | 33358012 | 33358012 | SNV | T | G:T | 1013 | G:G | 206 |
| **chr19** | 33358075 | 33358075 | SNV | A | A:G | 24 | | 0 |
| **chr19** | 33358079 | 33358079 | SNV | G | A:A | 7 | A:G | 91 |
| **chr19** | 33358231 | 33358231 | SNV | G | A:G | 13 | | 0 |
| **chr19** | 33358262 | 33358262 | SNV | A | A:G | 607 | G:G | 105 |
| **chr19** | 33358355 | 33358355 | SNV | T | C:T | 1236 | C:C | 703 |
| **chr19** | 33358428 | 33358428 | SNV | A | A:T | 3 | | 0 |
| **chr19** | 33358438 | 33358438 | SNV | A | A:T | 7 | | 0 |
| **chr19** | 33358442 | 33358442 | SNV | A | A:T | 159 | T:T | 5 |
| **chr19** | 33358527 | 33358527 | SNV | A | A:C | 611 | C:C | 105 |
| **chr19** | 33358629 | 33358629 | SNV | T | C:T | 28 | | 0 |
| **chr19** | 33358687 | 33358687 | SNV | C | A:C | 20 | | 0 |
| **chr19** | 33358712 | 33358712 | SNV | G | A:G | 74 | A:A | 2 |
| **chr19** | 33358716 | 33358716 | SNV | T | A:T | 20 | | 0 |
| **chr19** | 33358887 | 33358887 | SNV | C | C:G | 28 | | 0 |
| **chr19** | 33358963 | 33358963 | SNV | C | G:G | 97 | C:G | 588 |
| **chr19** | 33358998 | 33358998 | SNV | A | A:G | 1236 | G:G | 703 |
| **chr19** | 33359006 | 33359006 | SNV | G | A:G | 11 | | 0 |
| **chr19** | 33359008 | 33359008 | SNV | G | G:T | 11 | | 0 |
| **chr19** | 33359047 | 33359047 | SNV | G | A:G | 20 | | 0 |
| **chr19** | 33359061 | 33359061 | SNV | G | A:A | 107 | A:G | 607 |
| **chr19** | 33359078 | 33359078 | SNV | A | A:G | 20 | | 0 |
| **chr19** | 33359079 | 33359079 | SNV | T | G:T | 1177 | G:G | 556 |
| **chr19** | 33359240 | 33359240 | SNV | A | A:G | 1026 | G:G | 221 |
| **chr19** | 33359479 | 33359479 | SNV | G | A:G | 637 | A:A | 106 |
| **chr19** | 33359679 | 33359679 | SNV | G | A:G | 24 | | 0 |
| **chr19** | 33359723 | 33359723 | SNV | A | A:C | 13 | | 0 |
| **chr19** | 33359778 | 33359778 | SNV | T | G:T | 1 | | 0 |
| **chr19** | 33359791 | 33359791 | SNV | A | A:T | 701 | T:T | 70 |
| **chr19** | 33359832 | 33359832 | SNV | G | A:G | 927 | A:A | 186 |
| **chr19** | 33359943 | 33359943 | SNV | G | A:A | 110 | A:G | 617 |
| **chr19** | 33359960 | 33359960 | deletion | A | -:- | 387 | -:A | 1352 |
| **chr19** | 33359984 | 33359984 | SNV | T | C:C | 1 | C:T | 36 |
| **chr19** | 33360053 | 33360053 | SNV | C | A:A | 7 | A:C | 180 |
| **chr19** | 33360083 | 33360083 | SNV | C | G:G | 32 | C:G | 450 |
| **chr19** | 33360121 | 33360122 | deletion | TC | -:TC | 11 | | 0 |
| **chr19** | 33360131 | 33360131 | SNV | A | C:C | 5 | A:C | 89 |
| **chr19** | 33360214 | 33360214 | SNV | T | C:T | 1 | | 0 |
| **chr19** | 33360306 | 33360306 | insertion | - | -:T | 17 | | 0 |
| **chr19** | 33360341 | 33360341 | SNV | C | C:T | 41 | | 0 |
| **chr19** | 33360342 | 33360342 | SNV | G | A:A | 21 | A:G | 314 |
| **chr19** | 33360369 | 33360369 | SNV | C | A:C | 1225 | A:A | 560 |
| **chr19** | 33360409 | 33360409 | SNV | C | C:T | 28 | | 0 |
| **chr19** | 33360575 | 33360575 | SNV | C | G:G | 9 | C:G | 111 |
| **chr19** | 33360652 | 33360652 | SNV | A | T:T | 763 | A:T | 1177 |
| **chr19** | 33346332 | 33346332 | SNV | G | A:G | 3 | | 0 |
| **chr19** | 33346603 | 33346603 | SNV | G | A:G | 1 | | 0 |
| **chr19** | 33332649 | 33332649 | SNV | C | C:G | 10 | | 0 |
| **chr19** | 33354259 | 33354259 | SNV | G | A:G | 1 | | 0 |
| **chr19** | 33333116 | 33333116 | SNV | G\|I | A:G\|I:I | 3 | | 0 |
| **chr19** | 33346564 | 33346564 | SNV | T | C:T | 5 | | 0 |
| **chr19** | 33326797 | 33326797 | SNV | G | C:G | 11 | | 0 |
| **chr19** | 33333179 | 33333179 | SNV | C\|S | C:T\|S:S | 50 | | 0 |
| **chr19** | 33324286 | 33324289 | deletion | AAAA | -:AAAA | 5 | | 0 |
| **chr19** | 33338812 | 33338812 | SNV | T | G:T | 1 | | 0 |
| **chr19** | 33359317 | 33359317 | SNV | A | A:G | 2 | | 0 |
| **chr19** | 33336876 | 33336876 | insertion | - | -:TAG | 10 | | 0 |
| **chr19** | 33326009 | 33326009 | SNV | T | G:T | 14 | G:G | 1 |
| **chr19** | 33323739 | 33323739 | SNV | C | C:T | 3 | | 0 |
| **chr19** | 33324959 | 33324962 | deletion | AGGC | -:AGGC | 46 | | 0 |
| **chr19** | 33356989 | 33356989 | SNV | C | A:A | 1 | A:C | 14 |
| **chr19** | 33321608 | 33321608 | SNV | A | A:G | 6 | | 0 |
| **chr19** | 33353508 | 33353508 | SNV | C | C:T | 1 | | 0 |
| **chr19** | 33342947 | 33342947 | SNV | G | A:G | 1 | | 0 |
| **chr19** | 33345245 | 33345245 | SNV | G | A:G | 1 | | 0 |
| **chr19** | 33355594 | 33355594 | SNV | G\|T | A:G\|M:T | 1 | | 0 |
| **chr19** | 33356002 | 33356002 | SNV | G | G:T | 2 | | 0 |
| **chr19** | 33355747 | 33355747 | SNV | C | C:T | 7 | | 0 |
| **chr19** | 33355066 | 33355066 | SNV | C\|A | C:T\|A:A | 1 | | 0 |
| **chr19** | 33351922 | 33351922 | SNV | G | A:G | 7 | | 0 |
| **chr19** | 33323334 | 33323334 | SNV | A | A:G | 1 | | 0 |
| **chr19** | 33330181 | 33330181 | SNV | C | C:T | 1 | | 0 |
| **chr19** | 33358056 | 33358056 | SNV | T | C:T | 8 | | 0 |
| **chr19** | 33353488 | 33353488 | SNV | G\|F | A:G\|F:F | 2 | | 0 |
| **chr19** | 33326465 | 33326465 | SNV | A | A:C | 12 | | 0 |
| **chr19** | 33334350 | 33334350 | SNV | G | A:G | 1 | | 0 |
| **chr19** | 33337919 | 33337919 | SNV | A | A:G | 5 | | 0 |
| **chr19** | 33337606 | 33337606 | SNV | G | A:G | 3 | | 0 |

**Supplementary Table 2. Variants of SLC7A9 in the 1KG database**

| Chr | Start Position | End Position | Variant type | Ref Genotype \| Ref Proteotype | Variant Genotype 1\| Variant Proteotype 1 | Count | Variant Genotype 2\| Variant Proteotype 2 | Count |
|---|---|---|---|---|---|---|---|---|
| chr19 | 33349753 | 33349753 | SNV | G | G:T | 1 | | 0 |
| chr19 | 33336001 | 33336001 | SNV | C | A:C | 1 | | 0 |
| chr19 | 33357148 | 33357148 | SNV | G | G:T | 2 | | 0 |
| chr19 | 33344106 | 33344106 | SNV | C | T:T | 2 | C:T | 31 |
| chr19 | 33329644 | 33329644 | SNV | C | C:T | 8 | | 0 |
| chr19 | 33336975 | 33336975 | SNV | G | A:G | 3 | | 0 |
| chr19 | 33354535 | 33354535 | SNV | G | A:G | 1 | | 0 |
| chr19 | 33337233 | 33337233 | SNV | C | C:T | 12 | | 0 |
| chr19 | 33358017 | 33358017 | SNV | C | C:G | 1 | | 0 |
| chr19 | 33360320 | 33360320 | SNV | C | C:T | 2 | | 0 |
| chr19 | 33322185 | 33322185 | SNV | A | A:G | 3 | | 0 |
| chr19 | 33332857 | 33332857 | SNV | G | A:G | 4 | | 0 |
| chr19 | 33357257 | 33357257 | SNV | G | A:G | 1 | | 0 |
| chr19 | 33333614 | 33333614 | SNV | G | A:G | 8 | | 0 |
| chr19 | 33326771 | 33326771 | SNV | C | C:T | 1 | | 0 |
| chr19 | 33355773 | 33355773 | SNV | G | A:G | 14 | | 0 |
| chr19 | 33324453 | 33324453 | SNV | C | C:T | 10 | | 0 |
| chr19 | 33354215 | 33354215 | SNV | T | G:T | 5 | | 0 |
| chr19 | 33329186 | 33329186 | SNV | A | A:G | 4 | | 0 |
| chr19 | 33330143 | 33330143 | SNV | C | C:T | 5 | | 0 |
| chr19 | 33337234 | 33337234 | SNV | G | A:G | 1 | | 0 |
| chr19 | 33345745 | 33345745 | SNV | C | C:T | 1 | | 0 |
| chr19 | 33352872 | 33352872 | SNV | G | A:G | 2 | | 0 |
| chr19 | 33338335 | 33338335 | SNV | G | A:G | 2 | | 0 |
| chr19 | 33333327 | 33333327 | SNV | A | A:G | 1 | | 0 |
| chr19 | 33354154 | 33354154 | SNV | A | A:G | 4 | | 0 |
| chr19 | 33351478 | 33351478 | SNV | C | C:T | 1 | | 0 |
| chr19 | 33323091 | 33323091 | SNV | A | A:T | 3 | | 0 |
| chr19 | 33344887 | 33344887 | SNV | T | G:T | 2 | | 0 |
| chr19 | 33332334 | 33332334 | SNV | A | A:G | 1 | | 0 |
| chr19 | 33331398 | 33331398 | SNV | C | C:T | 4 | | 0 |
| chr19 | 33360644 | 33360644 | SNV | G | A:G | 1 | | 0 |
| chr19 | 33323017 | 33323017 | SNV | A | A:G | 8 | | 0 |
| chr19 | 33336672 | 33336672 | SNV | G | A:G | 12 | | 0 |
| chr19 | 33335049 | 33335049 | SNV | G | G:T | 1 | | 0 |
| chr19 | 33344769 | 33344769 | SNV | G | C:G | 49 | | 0 |
| chr19 | 33328972 | 33328972 | SNV | C | C:T | 2 | | 0 |
| chr19 | 33327041 | 33327041 | SNV | G | A:G | 13 | | 0 |
| chr19 | 33344259 | 33344259 | SNV | A | A:G | 1 | | 0 |
| chr19 | 33356446 | 33356446 | SNV | G | A:G | 5 | | 0 |
| chr19 | 33337182 | 33337182 | SNV | A | A:T | 3 | | 0 |
| chr19 | 33341957 | 33341957 | SNV | G | A:G | 1 | | 0 |
| chr19 | 33354896 | 33354896 | SNV | C | C:T | 3 | | 0 |
| chr19 | 33325926 | 33325926 | SNV | C | C:G | 7 | | 0 |
| chr19 | 33342499 | 33342499 | SNV | C | C:T | 3 | | 0 |
| chr19 | 33329610 | 33329610 | SNV | A | A:T | 1 | | 0 |
| chr19 | 33321586 | 33321586 | SNV | C\|P | C:T\|P:P | 6 | | 0 |
| chr19 | 33326291 | 33326291 | SNV | A | A:T | 2 | | 0 |
| chr19 | 33360294 | 33360294 | SNV | A | A:C | 1 | | 0 |
| chr19 | 33334875 | 33334875 | SNV | G | A:G | 1 | | 0 |
| chr19 | 33346464 | 33346464 | SNV | C | A:C | 2 | | 0 |
| chr19 | 33343032 | 33343032 | SNV | A | A:G | 1 | | 0 |
| chr19 | 33349559 | 33349559 | SNV | C | C:T | 5 | | 0 |
| chr19 | 33323489 | 33323489 | SNV | G | C:G | 12 | | 0 |
| chr19 | 33346855 | 33346855 | SNV | G | G:T | 3 | | 0 |
| chr19 | 33334723 | 33334723 | SNV | C | C:T | 1 | | 0 |
| chr19 | 33355719 | 33355719 | SNV | G | A:G | 2 | | 0 |
| chr19 | 33337364 | 33337364 | SNV | C | C:T | 9 | | 0 |
| chr19 | 33347186 | 33347186 | SNV | T | C:C | 1 | C:T | 18 |
| chr19 | 33350757 | 33350757 | SNV | G\|A | A:G\|V:A | 1 | | 0 |
| chr19 | 33337359 | 33337359 | SNV | G | A:G | 1 | | 0 |
| chr19 | 33347998 | 33347998 | SNV | G | G:T | 1 | | 0 |
| chr19 | 33335189 | 33335189 | SNV | T | A:T | 3 | | 0 |
| chr19 | 33350463 | 33350463 | SNV | G | G:T | 48 | | 0 |
| chr19 | 33337001 | 33337001 | SNV | G | A:G | 1 | | 0 |
| chr19 | 33346472 | 33346472 | SNV | A | A:G | 2 | | 0 |
| chr19 | 33345791 | 33345791 | SNV | G | A:G | 1 | | 0 |
| chr19 | 33359130 | 33359130 | SNV | C | C:T | 1 | | 0 |
| chr19 | 33331639 | 33331639 | SNV | C | C:T | 1 | | 0 |
| chr19 | 33322172 | 33322172 | SNV | T | C:T | 1 | | 0 |
| chr19 | 33336991 | 33336991 | SNV | C | C:T | 3 | | 0 |
| chr19 | 33345910 | 33345910 | SNV | T | A:T | 2 | | 0 |
| chr19 | 33344844 | 33344844 | SNV | C | C:T | 9 | | 0 |
| chr19 | 33324683 | 33324683 | SNV | A | A:G | 10 | | 0 |
| chr19 | 33324565 | 33324565 | SNV | G | A:G | 7 | | 0 |
| chr19 | 33353449 | 33353449 | SNV | G\|S | A:G\|S:S | 10 | | 0 |
| chr19 | 33356583 | 33356583 | SNV | G | T:T | 1 | G:T | 19 |
| chr19 | 33346438 | 33346438 | SNV | T | C:T | 13 | | 0 |
| chr19 | 33332313 | 33332313 | SNV | G | A:G | 5 | | 0 |
| chr19 | 33337531 | 33337531 | SNV | C | C:T | 1 | | 0 |
| chr19 | 33339720 | 33339720 | SNV | C | C:T | 10 | | 0 |
| chr19 | 33330996 | 33330996 | SNV | A | A:T | 1 | | 0 |
| chr19 | 33344277 | 33344277 | SNV | T | C:T | 3 | C:C | 1 |
| chr19 | 33360383 | 33360383 | SNV | G | G:T | 5 | | 0 |
| chr19 | 33357926 | 33357926 | SNV | C | A:C | 1 | | 0 |
| chr19 | 33331511 | 33331511 | SNV | A | A:T | 14 | | 0 |
| chr19 | 33350637 | 33350637 | SNV | G | A:G | 2 | | 0 |
| chr19 | 33347790 | 33347790 | SNV | G | A:G | 1 | | 0 |
| chr19 | 33356467 | 33356467 | SNV | G | A:G | 1 | | 0 |
| chr19 | 33336386 | 33336386 | SNV | C | C:T | 4 | | 0 |
| chr19 | 33328646 | 33328646 | SNV | A | A:G | 1 | | 0 |
| chr19 | 33326536 | 33326536 | SNV | C | A:C | 4 | | 0 |
| chr19 | 33342068 | 33342068 | SNV | A | A:G | 17 | | 0 |
| chr19 | 33358904 | 33358904 | SNV | C | C:T | 7 | | 0 |
| chr19 | 33346556 | 33346556 | SNV | C | C:G | 9 | | 0 |
| chr19 | 33340000 | 33340000 | SNV | A | A:C | 8 | | 0 |
| chr19 | 33360615 | 33360615 | SNV | C | C:T | 2 | | 0 |
| chr19 | 33322354 | 33322354 | SNV | G | A:G | 1 | | 0 |
| chr19 | 33343965 | 33343965 | SNV | G | A:G | 1 | | 0 |
| chr19 | 33348822 | 33348822 | SNV | G | C:G | 1 | | 0 |
| chr19 | 33324212 | 33324212 | SNV | G\|P | A:G\|P:P | 4 | | 0 |
| chr19 | 33335695 | 33335695 | SNV | C | A:C | 12 | | 0 |
| chr19 | 33331888 | 33331888 | SNV | T | C:T | 1 | | 0 |
| chr19 | 33335800 | 33335800 | SNV | A | A:G | 3 | | 0 |
| chr19 | 33346333 | 33346333 | SNV | C | C:T | 21 | | 0 |
| chr19 | 33335515 | 33335515 | SNV | C | C:T | 1 | | 0 |
| chr19 | 33332878 | 33332878 | SNV | T | G:T | 2 | | 0 |
| chr19 | 33354134 | 33354134 | deletion | C | -:C | 5 | | 0 |
| chr19 | 33324303 | 33324304 | deletion | AA | -:AA | 3 | | 0 |
| chr19 | 33353696 | 33353696 | SNV | G | C:G | 24 | | 0 |
| chr19 | 33325067 | 33325067 | SNV | A | A:C | 9 | | 0 |
| chr19 | 33350418 | 33350418 | SNV | T | C:T | 31 | | 0 |
| chr19 | 33343428 | 33343428 | SNV | C | C:T | 1 | | 0 |
| chr19 | 33345515 | 33345515 | SNV | C | C:T | 1 | | 0 |
| chr19 | 33324578 | 33324578 | SNV | C | C:T | 26 | T:T | 1 |
| chr19 | 33353228 | 33353228 | SNV | G | A:G | 1 | | 0 |
| chr19 | 33331571 | 33331571 | SNV | C | A:C | 4 | | 0 |
| chr19 | 33345773 | 33345773 | SNV | C | C:T | 4 | | 0 |
| chr19 | 33331136 | 33331136 | SNV | T | C:T | 6 | | 0 |
| chr19 | 33331663 | 33331663 | SNV | C | C:T | 2 | | 0 |
| chr19 | 33343850 | 33343850 | SNV | C | C:T | 5 | | 0 |
| chr19 | 33331142 | 33331142 | SNV | G | A:G | 30 | | 0 |
| chr19 | 33353576 | 33353576 | insertion | - | -:G | 5 | | 0 |
| chr19 | 33331016 | 33331016 | SNV | G | A:G | 1 | | 0 |
| chr19 | 33345564 | 33345564 | SNV | G | A:G | 2 | | 0 |
| chr19 | 33354104 | 33354104 | SNV | A | A:T | 5 | | 0 |
| chr19 | 33349322 | 33349322 | SNV | G | A:G | 5 | | 0 |
| chr19 | 33358821 | 33358821 | SNV | G | A:G | 6 | | 0 |
| chr19 | 33356009 | 33356009 | SNV | G | A:G | 6 | | 0 |
| chr19 | 33351229 | 33351229 | SNV | G | A:G | 15 | | 0 |
| chr19 | 33348038 | 33348038 | SNV | A | A:G | 6 | | 0 |

**Supplementary Table 2. Variants of SLC7A9 in the 1KG database**

| Chr | Start Position | End Position | Variant type | Ref Genotype \| Ref Proteotype | Variant Genotype 1\| Variant Proteotype 1 | Count | Variant Genotype 2\| Variant Proteotype 2 | Count |
|---|---|---|---|---|---|---|---|---|
| **chr19** | 33347249 | 33347249 | SNV | C | C:T | 8 | | 0 |
| **chr19** | 33338312 | 33338312 | SNV | C | C:T | 1 | | 0 |
| **chr19** | 33354426 | 33354426 | SNV | C | C:T | 2 | | 0 |
| **chr19** | 33336102 | 33336102 | SNV | T | A:T | 24 | | 0 |
| **chr19** | 33335612 | 33335612 | SNV | C | C:T | 1 | | 0 |
| **chr19** | 33353954 | 33353954 | SNV | G | A:G | 3 | | 0 |
| **chr19** | 33348699 | 33348699 | SNV | C | A:C | 8 | A:A | 1 |
| **chr19** | 33352510 | 33352510 | SNV | G | A:G | 2 | | 0 |
| **chr19** | 33337596 | 33337596 | SNV | C | C:G | 1 | | 0 |
| **chr19** | 33332958 | 33332958 | SNV | T | C:T | 1 | | 0 |
| **chr19** | 33335624 | 33335624 | SNV | C | C:T | 1 | | 0 |
| **chr19** | 33356081 | 33356081 | SNV | C | C:T | 10 | | 0 |
| **chr19** | 33343709 | 33343709 | SNV | G | A:G | 26 | A:A | 1 |
| **chr19** | 33337671 | 33337671 | SNV | C | A:C | 1 | | 0 |
| **chr19** | 33354681 | 33354681 | SNV | G | A:G | 5 | | 0 |
| **chr19** | 33349302 | 33349302 | SNV | C | C:T | 1 | | 0 |
| **chr19** | 33349623 | 33349623 | SNV | G | A:G | 1 | | 0 |
| **chr19** | 33346118 | 33346118 | SNV | C | C:T | 1 | | 0 |
| **chr19** | 33322286 | 33322286 | SNV | G | A:G | 1 | | 0 |
| **chr19** | 33348089 | 33348089 | SNV | G | G:T | 1 | | 0 |
| **chr19** | 33326854 | 33326854 | SNV | T | C:T | 2 | | 0 |
| **chr19** | 33360331 | 33360331 | SNV | C | C:T | 1 | | 0 |
| **chr19** | 33358625 | 33358625 | SNV | C | C:T | 9 | | 0 |
| **chr19** | 33344136 | 33344136 | SNV | G | C:G | 5 | | 0 |
| **chr19** | 33322349 | 33322349 | SNV | C | C:T | 1 | | 0 |
| **chr19** | 33328083 | 33328083 | insertion | - | -:T | 8 | | 0 |
| **chr19** | 33342997 | 33342997 | SNV | G | A:G | 1 | | 0 |
| **chr19** | 33353496 | 33353496 | SNV | G | A:G | 1 | | 0 |
| **chr19** | 33336918 | 33336918 | SNV | A | A:G | 8 | | 0 |
| **chr19** | 33357713 | 33357713 | SNV | T | C:T | 1 | | 0 |
| **chr19** | 33328525 | 33328525 | SNV | T | G:T | 3 | | 0 |
| **chr19** | 33331678 | 33331678 | SNV | C | C:T | 1 | | 0 |
| **chr19** | 33323044 | 33323044 | SNV | A | A:G | 2 | | 0 |
| **chr19** | 33339402 | 33339402 | SNV | A | A:G | 2 | | 0 |
| **chr19** | 33335385 | 33335385 | SNV | A | A:G | 8 | | 0 |
| **chr19** | 33344738 | 33344738 | SNV | G | A:G | 9 | | 0 |
| **chr19** | 33359462 | 33359462 | SNV | G | A:G | 1 | | 0 |
| **chr19** | 33356294 | 33356294 | SNV | C | C:T | 21 | | 0 |
| **chr19** | 33336101 | 33336101 | SNV | A | A:G | 2 | | 0 |
| **chr19** | 33345763 | 33345763 | SNV | G | G:T | 1 | | 0 |
| **chr19** | 33324004 | 33324004 | SNV | G | A:G | 1 | | 0 |
| **chr19** | 33358336 | 33358336 | SNV | T | C:T | 11 | | 0 |
| **chr19** | 33321544 | 33321544 | SNV | C\|P | C:T\|P:P | 1 | | 0 |
| **chr19** | 33345172 | 33345172 | SNV | G | A:G | 49 | | 0 |
| **chr19** | 33332863 | 33332863 | SNV | G | A:G | 3 | | 0 |
| **chr19** | 33327065 | 33327065 | SNV | G | A:G | 3 | | 0 |
| **chr19** | 33353578 | 33353578 | SNV | T | A:T | 8 | | 0 |
| **chr19** | 33325852 | 33325852 | SNV | A | A:C | 2 | | 0 |
| **chr19** | 33350045 | 33350045 | SNV | G | A:G | 1 | | 0 |
| **chr19** | 33332411 | 33332411 | SNV | C | C:T | 3 | | 0 |
| **chr19** | 33353445 | 33353445 | SNV | C\|V | C:T\|V:I | 3 | | 0 |
| **chr19** | 33347433 | 33347433 | SNV | A | A:G | 2 | | 0 |
| **chr19** | 33350372 | 33350372 | SNV | C | C:T | 5 | | 0 |
| **chr19** | 33352700 | 33352700 | SNV | G | A:G | 1 | | 0 |
| **chr19** | 33327331 | 33327331 | SNV | C | C:T | 4 | | 0 |
| **chr19** | 33348632 | 33348632 | SNV | G | C:G | 2 | | 0 |
| **chr19** | 33357866 | 33357866 | SNV | G | A:G | 2 | | 0 |
| **chr19** | 33335642 | 33335642 | SNV | C | C:T | 1 | | 0 |
| **chr19** | 33332647 | 33332647 | SNV | T | C:T | 10 | | 0 |
| **chr19** | 33341555 | 33341555 | SNV | G | G:T | 12 | | 0 |
| **chr19** | 33356390 | 33356390 | SNV | G | C:G | 1 | | 0 |
| **chr19** | 33326939 | 33326939 | SNV | C | C:T | 1 | | 0 |
| **chr19** | 33348012 | 33348012 | SNV | C | C:T | 4 | | 0 |
| **chr19** | 33327043 | 33327043 | SNV | T | C:T | 10 | | 0 |
| **chr19** | 33339219 | 33339219 | SNV | A | A:G | 12 | | 0 |
| **chr19** | 33353345 | 33353345 | SNV | G | A:G | 5 | | 0 |
| **chr19** | 33357057 | 33357057 | SNV | C | C:T | 3 | | 0 |
| **chr19** | 33354843 | 33354843 | SNV | G | A:G | 1 | | 0 |
| **chr19** | 33329006 | 33329006 | SNV | G | A:G | 2 | | 0 |
| **chr19** | 33359029 | 33359029 | SNV | C | A:A | 3 | A:C | 52 |
| **chr19** | 33344108 | 33344108 | SNV | C | C:T | 2 | | 0 |
| **chr19** | 33322458 | 33322458 | SNV | T | C:T | 1 | | 0 |
| **chr19** | 33356068 | 33356068 | SNV | T | C:T | 1 | | 0 |
| **chr19** | 33350791 | 33350791 | SNV | C\|V | C:T\|V:M | 5 | | 0 |
| **chr19** | 33353446 | 33353446 | SNV | G\|Y | A:G\|Y:Y | 1 | | 0 |
| **chr19** | 33359086 | 33359086 | SNV | G | A:G | 19 | A:A | 1 |
| **chr19** | 33358354 | 33358354 | SNV | T | C:T | 1 | | 0 |
| **chr19** | 33352340 | 33352340 | SNV | C | C:T | 2 | | 0 |
| **chr19** | 33345760 | 33345760 | SNV | C | C:T | 1 | | 0 |
| **chr19** | 33335933 | 33335933 | SNV | G | A:G | 10 | | 0 |
| **chr19** | 33354132 | 33354132 | SNV | C | C:T | 9 | T:T | 2 |
| **chr19** | 33357225 | 33357225 | SNV | A | A:G | 1 | | 0 |
| **chr19** | 33355111 | 33355111 | SNV | C\|T | C:T\|T:T | 5 | | 0 |
| **chr19** | 33349293 | 33349293 | SNV | T | C:T | 19 | C:C | 1 |
| **chr19** | 33356237 | 33356240 | deletion | TCTC | -:TCTC | 5 | | 0 |
| **chr19** | 33354258 | 33354258 | SNV | C | C:T | 1 | | 0 |
| **chr19** | 33337832 | 33337832 | SNV | G | G:T | 8 | | 0 |
| **chr19** | 33330599 | 33330599 | SNV | A | A:C | 1 | | 0 |
| **chr19** | 33353358 | 33353358 | SNV | G | A:G | 4 | | 0 |
| **chr19** | 33348328 | 33348328 | SNV | C | C:T | 49 | | 0 |
| **chr19** | 33353398 | 33353398 | SNV | G\|I | G:T\|I:I | 5 | | 0 |
| **chr19** | 33331791 | 33331791 | SNV | G | A:G | 1 | | 0 |
| **chr19** | 33324073 | 33324073 | SNV | A\|W | A:G\|W:R | 1 | | 0 |
| **chr19** | 33358313 | 33358313 | SNV | C | C:G | 1 | | 0 |
| **chr19** | 33351506 | 33351506 | SNV | T | C:C | 1 | C:T | 27 |
| **chr19** | 33348601 | 33348601 | SNV | G | A:G | 1 | | 0 |
| **chr19** | 33346557 | 33346557 | SNV | G | A:G | 1 | | 0 |
| **chr19** | 33338552 | 33338552 | SNV | G | G:T | 27 | T:T | 1 |
| **chr19** | 33328260 | 33328260 | SNV | G | G:T | 2 | | 0 |
| **chr19** | 33338956 | 33338956 | SNV | A | A:G | 1 | | 0 |
| **chr19** | 33352443 | 33352443 | SNV | C | C:T | 11 | | 0 |
| **chr19** | 33355789 | 33355789 | SNV | C | C:T | 11 | | 0 |
| **chr19** | 33336167 | 33336167 | SNV | G | A:G | 3 | | 0 |
| **chr19** | 33337137 | 33337137 | SNV | C | C:T | 1 | | 0 |
| **chr19** | 33345291 | 33345291 | SNV | C | C:T | 1 | | 0 |
| **chr19** | 33329030 | 33329030 | SNV | A | A:G | 3 | | 0 |
| **chr19** | 33345587 | 33345587 | SNV | C | C:T | 4 | | 0 |
| **chr19** | 33348051 | 33348051 | SNV | A | A:G | 1 | | 0 |
| **chr19** | 33333482 | 33333482 | SNV | G | A:G | 3 | | 0 |
| **chr19** | 33345947 | 33345947 | SNV | C | C:T | 1 | | 0 |
| **chr19** | 33337212 | 33337212 | SNV | G | A:G | 7 | | 0 |
| **chr19** | 33338489 | 33338489 | SNV | G | A:G | 2 | | 0 |
| **chr19** | 33339759 | 33339759 | SNV | T | C:T | 4 | | 0 |
| **chr19** | 33344218 | 33344218 | SNV | G | A:G | 1 | | 0 |
| **chr19** | 33338729 | 33338729 | SNV | C | C:G | 1 | | 0 |
| **chr19** | 33335388 | 33335388 | SNV | C | C:G | 4 | | 0 |
| **chr19** | 33347453 | 33347453 | SNV | C | C:T | 1 | | 0 |
| **chr19** | 33334746 | 33334746 | SNV | G | A:G | 1 | | 0 |
| **chr19** | 33359199 | 33359199 | insertion | - | -:CAGA | 13 | | 0 |
| **chr19** | 33327143 | 33327143 | SNV | T | G:T | 4 | | 0 |
| **chr19** | 33353479 | 33353479 | SNV | T\|T | C:T\|T:T | 4 | | 0 |
| **chr19** | 33346630 | 33346630 | SNV | A | A:C | 13 | | 0 |
| **chr19** | 33357782 | 33357782 | SNV | A | A:C | 2 | | 0 |
| **chr19** | 33332371 | 33332371 | SNV | C | A:C | 1 | | 0 |
| **chr19** | 33328461 | 33328461 | SNV | G | A:G | 4 | | 0 |
| **chr19** | 33334298 | 33334298 | SNV | G | G:T | 1 | | 0 |
| **chr19** | 33352991 | 33352991 | SNV | C | T:T | 1 | C:T | 27 |
| **chr19** | 33359836 | 33359836 | SNV | T | C:T | 2 | | 0 |
| **chr19** | 33351184 | 33351184 | SNV | C | C:T | 1 | | 0 |
| **chr19** | 33332373 | 33332373 | SNV | T | C:T | 1 | | 0 |
| **chr19** | 33332645 | 33332645 | SNV | T | C:T | 10 | | 0 |

**Supplementary Table 2. Variants of SLC7A9 in the 1KG database**

| Chr | Start Position | End Position | Variant type | Ref Genotype \| Ref Proteotype | Variant Genotype 1\| Variant Proteotype 1 | Count | Variant Genotype 2\| Variant Proteotype 2 | Count |
|---|---|---|---|---|---|---|---|---|
| **chr19** | 33353315 | 33353315 | SNV | T | C:T | 2 | | 0 |
| **chr19** | 33352926 | 33352926 | insertion | - | -:A | 36 | | 0 |
| **chr19** | 33354191 | 33354191 | SNV | G | A:G | 1 | | 0 |
| **chr19** | 33331164 | 33331164 | SNV | G | A:G | 7 | | 0 |
| **chr19** | 33337334 | 33337334 | SNV | G | C:G | 14 | C:C | 1 |
| **chr19** | 33349284 | 33349284 | SNV | C | C:T | 11 | | 0 |
| **chr19** | 33327096 | 33327096 | SNV | G | A:G | 1 | | 0 |
| **chr19** | 33358028 | 33358028 | SNV | C | C:G | 14 | G:G | 1 |
| **chr19** | 33358359 | 33358359 | SNV | G | A:G | 5 | | 0 |
| **chr19** | 33335651 | 33335651 | SNV | C | C:T | 1 | | 0 |
| **chr19** | 33347748 | 33347748 | SNV | A | A:G | 7 | | 0 |
| **chr19** | 33346477 | 33346477 | SNV | A | A:G | 5 | | 0 |
| **chr19** | 33333445 | 33333445 | SNV | G | A:G | 1 | | 0 |
| **chr19** | 33328591 | 33328591 | SNV | G | A:G | 8 | | 0 |
| **chr19** | 33352267 | 33352267 | SNV | A | A:G | 1 | | 0 |
| **chr19** | 33327881 | 33327881 | SNV | C | A:A | 1 | A:C | 22 |
| **chr19** | 33334753 | 33334753 | SNV | G | C:G | 1 | | 0 |
| **chr19** | 33323507 | 33323507 | SNV | A | A:G | 4 | | 0 |
| **chr19** | 33351785 | 33351785 | SNV | G | G:T | 4 | | 0 |
| **chr19** | 33354427 | 33354427 | SNV | G | A:G | 1 | | 0 |
| **chr19** | 33356968 | 33356968 | SNV | C | C:T | 2 | | 0 |
| **chr19** | 33330257 | 33330257 | SNV | G | A:G | 3 | | 0 |
| **chr19** | 33345026 | 33345026 | SNV | C | C:T | 10 | | 0 |
| **chr19** | 33349475 | 33349475 | SNV | T | C:T | 3 | | 0 |
| **chr19** | 33349696 | 33349696 | SNV | T | G:T | 7 | | 0 |
| **chr19** | 33354719 | 33354719 | SNV | G | A:G | 7 | | 0 |
| **chr19** | 33357670 | 33357670 | SNV | C | C:T | 1 | | 0 |
| **chr19** | 33347250 | 33347250 | SNV | G | A:G | 1 | | 0 |
| **chr19** | 33332909 | 33332909 | SNV | G | A:G | 6 | | 0 |
| **chr19** | 33329041 | 33329041 | SNV | G | G:T | 1 | | 0 |
| **chr19** | 33352653 | 33352653 | SNV | C | C:T | 12 | | 0 |
| **chr19** | 33333299 | 33333299 | SNV | A | A:G | 1 | | 0 |
| **chr19** | 33356430 | 33356430 | SNV | C | C:T | 5 | | 0 |
| **chr19** | 33343738 | 33343738 | SNV | T | C:T | 3 | | 0 |
| **chr19** | 33334294 | 33334294 | SNV | C | C:T | 2 | | 0 |
| **chr19** | 33344209 | 33344209 | SNV | G | A:G | 33 | | 0 |
| **chr19** | 33327942 | 33327942 | SNV | C | C:T | 6 | | 0 |
| **chr19** | 33345165 | 33345165 | SNV | C | C:T | 8 | | 0 |
| **chr19** | 33339903 | 33339903 | SNV | A | A:C | 9 | | 0 |
| **chr19** | 33355112 | 33355112 | SNV | G\|T | A:G\|M:T | 2 | | 0 |
| **chr19** | 33335196 | 33335196 | SNV | A | A:T | 2 | | 0 |
| **chr19** | 33321518 | 33321518 | SNV | G | A:G | 8 | | 0 |
| **chr19** | 33358443 | 33358443 | SNV | T | A:T | 1 | | 0 |
| **chr19** | 33354057 | 33354057 | SNV | G | A:G | 7 | | 0 |
| **chr19** | 33359464 | 33359464 | SNV | G | A:G | 1 | | 0 |
| **chr19** | 33325565 | 33325565 | SNV | A | A:C | 1 | | 0 |
| **chr19** | 33321991 | 33321991 | SNV | C | C:T | 1 | | 0 |
| **chr19** | 33326626 | 33326626 | SNV | C | A:C | 2 | | 0 |
| **chr19** | 33335534 | 33335534 | SNV | G | A:G | 7 | | 0 |
| **chr19** | 33346595 | 33346595 | SNV | A | A:G | 6 | | 0 |
| **chr19** | 33328658 | 33328658 | SNV | C | A:C | 1 | | 0 |
| **chr19** | 33345094 | 33345094 | SNV | T | C:T | 1 | | 0 |
| **chr19** | 33355270 | 33355270 | SNV | C | C:T | 1 | | 0 |
| **chr19** | 33359282 | 33359282 | SNV | A | A:C | 2 | | 0 |
| **chr19** | 33349463 | 33349463 | SNV | C | C:T | 1 | | 0 |
| **chr19** | 33353205 | 33353205 | SNV | T | G:T | 7 | | 0 |
| **chr19** | 33327838 | 33327838 | SNV | T | C:T | 1 | | 0 |
| **chr19** | 33332896 | 33332896 | SNV | A | A:G | 1 | | 0 |
| **chr19** | 33341734 | 33341734 | SNV | A | A:G | 1 | | 0 |
| **chr19** | 33329085 | 33329085 | SNV | G | A:G | 8 | | 0 |
| **chr19** | 33357392 | 33357392 | SNV | G | A:G | 5 | | 0 |
| **chr19** | 33349897 | 33349897 | SNV | G | A:G | 24 | A:A | 1 |
| **chr19** | 33345279 | 33345279 | SNV | C | C:T | 1 | | 0 |
| **chr19** | 33321810 | 33321810 | SNV | T | G:T | 2 | | 0 |
| **chr19** | 33357415 | 33357415 | SNV | C | C:T | 1 | | 0 |
| **chr19** | 33360159 | 33360159 | SNV | G | G:T | 8 | | 0 |
| **chr19** | 33356688 | 33356688 | SNV | C | C:G | 2 | | 0 |
| **chr19** | 33336850 | 33336850 | SNV | C | C:G | 1 | | 0 |
| **chr19** | 33330998 | 33331009 | deletion | CCCTTTCTACTT | -:CCCTTTCTACTT | 6 | | 0 |
| **chr19** | 33330383 | 33330383 | SNV | C | A:C | 1 | | 0 |
| **chr19** | 33355581 | 33355581 | SNV | C\|G | C:T\|G:G | 1 | | 0 |
| **chr19** | 33354605 | 33354605 | SNV | A | A:G | 6 | | 0 |
| **chr19** | 33351422 | 33351422 | SNV | G | G:T | 1 | | 0 |
| **chr19** | 33349148 | 33349148 | SNV | A | A:C | 12 | | 0 |
| **chr19** | 33341571 | 33341571 | SNV | T | C:T | 8 | | 0 |
| **chr19** | 33321545 | 33321545 | SNV | G\|P | A:G\|L:P | 1 | | 0 |
| **chr19** | 33350829 | 33350829 | SNV | G\|T | A:G\|M:T | 1 | | 0 |
| **chr19** | 33350730 | 33350730 | SNV | G | A:G | 1 | | 0 |
| **chr19** | 33353502 | 33353502 | SNV | T | C:T | 1 | | 0 |
| **chr19** | 33337705 | 33337705 | SNV | C | C:T | 11 | | 0 |
| **chr19** | 33326587 | 33326587 | SNV | A | A:C | 6 | | 0 |
| **chr19** | 33357142 | 33357142 | SNV | C | C:T | 21 | | 0 |
| **chr19** | 33331742 | 33331742 | SNV | A | A:G | 1 | | 0 |
| **chr19** | 33335994 | 33335994 | SNV | C | C:T | 2 | | 0 |
| **chr19** | 33347675 | 33347675 | SNV | G | A:G | 1 | | 0 |
| **chr19** | 33335742 | 33335742 | SNV | C | C:T | 1 | | 0 |
| **chr19** | 33330349 | 33330349 | SNV | C | C:T | 3 | | 0 |
| **chr19** | 33355848 | 33355848 | SNV | G | A:G | 2 | | 0 |
| **chr19** | 33336333 | 33336333 | SNV | G | A:G | 2 | | 0 |
| **chr19** | 33328662 | 33328662 | SNV | A | A:G | 1 | | 0 |
| **chr19** | 33331612 | 33331612 | SNV | G | A:G | 1 | | 0 |
| **chr19** | 33333131 | 33333131 | SNV | C\|T | C:T\|T:T | 1 | | 0 |
| **chr19** | 33360595 | 33360595 | SNV | C | C:T | 1 | | 0 |
| **chr19** | 33323536 | 33323536 | SNV | G | A:G | 1 | | 0 |
| **chr19** | 33324794 | 33324794 | SNV | T | C:T | 1 | | 0 |
| **chr19** | 33346343 | 33346343 | SNV | C | C:T | 1 | | 0 |
| **chr19** | 33338966 | 33338966 | SNV | C | C:T | 3 | | 0 |
| **chr19** | 33326486 | 33326486 | SNV | T | A:T | 2 | | 0 |
| **chr19** | 33356137 | 33356137 | SNV | C | C:T | 1 | | 0 |
| **chr19** | 33336342 | 33336342 | SNV | A | A:G | 3 | | 0 |
| **chr19** | 33348842 | 33348842 | SNV | G | G:T | 1 | | 0 |
| **chr19** | 33347886 | 33347886 | SNV | G | A:G | 1 | | 0 |
| **chr19** | 33330475 | 33330475 | SNV | A | A:C | 1 | | 0 |
| **chr19** | 33353570 | 33353570 | SNV | T | A:T | 1 | | 0 |
| **chr19** | 33332251 | 33332251 | SNV | G | A:G | 1 | | 0 |
| **chr19** | 33346678 | 33346678 | SNV | G | C:G | 2 | | 0 |
| **chr19** | 33322279 | 33322279 | SNV | G | A:G | 3 | | 0 |
| **chr19** | 33344109 | 33344109 | SNV | G | A:G | 1 | | 0 |
| **chr19** | 33360616 | 33360616 | SNV | G | A:G | 1 | | 0 |
| **chr19** | 33359396 | 33359396 | SNV | C\|S | C:T\|S:S | 1 | | 0 |
| **chr19** | 33332780 | 33332780 | SNV | T | G:T | 2 | | 0 |
| **chr19** | 33353428 | 33353428 | SNV | G\|T | A:G\|T:T | 1 | | 0 |
| **chr19** | 33338497 | 33338497 | SNV | A | A:C | 1 | | 0 |
| **chr19** | 33359499 | 33359499 | SNV | A | A:G | 1 | | 0 |
| **chr19** | 33342043 | 33342043 | SNV | C | C:G | 1 | | 0 |
| **chr19** | 33359810 | 33359812 | deletion | AAG | -:AAG | 5 | | 0 |
| **chr19** | 33331485 | 33331485 | SNV | T | C:T | 2 | | 0 |
| **chr19** | 33322771 | 33322771 | SNV | G | A:G | 1 | | 0 |
| **chr19** | 33332854 | 33332854 | SNV | C | C:T | 1 | | 0 |
| **chr19** | 33345343 | 33345343 | SNV | T | C:T | 1 | | 0 |
| **chr19** | 33333276 | 33333276 | SNV | A | A:G | 1 | | 0 |
| **chr19** | 33327531 | 33327531 | SNV | T | G:T | 2 | | 0 |
| **chr19** | 33346398 | 33346398 | SNV | G | G:T | 1 | | 0 |
| **chr19** | 33348032 | 33348032 | SNV | G | A:G | 1 | | 0 |
| **chr19** | 33353526 | 33353526 | SNV | T | C:T | 1 | | 0 |
| **chr19** | 33349122 | 33349122 | SNV | G | A:G | 1 | | 0 |
| **chr19** | 33330872 | 33330872 | SNV | C | C:T | 1 | | 0 |
| **chr19** | 33349448 | 33349448 | SNV | G\|T | A:G\|I:T | 1 | | 0 |
| **chr19** | 33335405 | 33335405 | SNV | T | A:T | 1 | | 0 |
| **chr19** | 33348161 | 33348161 | SNV | G | A:G | 1 | | 0 |
| **chr19** | 33341947 | 33341947 | SNV | T | G:T | 1 | | 0 |

**Supplementary Table 2. Variants of SLC7A9 in the 1KG database**

| Chr | Start Position | End Position | Variant type | Ref Genotype \| Ref Proteotype | Variant Genotype 1\| Variant Proteotype 1 | Count | Variant Genotype 2\| Variant Proteotype 2 | Count |
|---|---|---|---|---|---|---|---|---|
| **chr19** | 33349611 | 33349611 | SNV | C | C:T | 1 | | 0 |
| **chr19** | 33354551 | 33354551 | SNV | G | A:G | 1 | | 0 |
| **chr19** | 33337358 | 33337358 | SNV | C | C:T | 2 | | 0 |
| **chr19** | 33344111 | 33344111 | SNV | C | C:T | 1 | | 0 |
| **chr19** | 33334531 | 33334531 | SNV | C | C:T | 1 | | 0 |
| **chr19** | 33354927 | 33354927 | SNV | G | A:G | 1 | | 0 |
| **chr19** | 33333257 | 33333257 | SNV | A | A:G | 1 | | 0 |
| **chr19** | 33330821 | 33330821 | SNV | A | A:T | 1 | | 0 |
| **chr19** | 33328496 | 33328496 | SNV | T | A:T | 1 | | 0 |
| **chr19** | 33324428 | 33324428 | SNV | G | G:T | 2 | | 0 |
| **chr19** | 33335995 | 33335995 | SNV | G | A:G | 4 | | 0 |
| **chr19** | 33348235 | 33348235 | SNV | A | A:C | 1 | | 0 |
| **chr19** | 33359466 | 33359466 | SNV | C | A:C | 1 | | 0 |
| **chr19** | 33353459 | 33353459 | SNV | C\|R | C:T\|R:Q | 1 | | 0 |
| **chr19** | 33343571 | 33343571 | SNV | G | A:G | 1 | | 0 |
| **chr19** | 33355504 | 33355504 | SNV | G | A:G | 2 | | 0 |
| **chr19** | 33360145 | 33360145 | SNV | A | A:T | 2 | | 0 |
| **chr19** | 33357704 | 33357704 | SNV | C | C:T | 1 | | 0 |
| **chr19** | 33332216 | 33332216 | SNV | A | A:G | 1 | | 0 |
| **chr19** | 33356799 | 33356799 | SNV | C | C:T | 1 | | 0 |
| **chr19** | 33323623 | 33323623 | SNV | G | A:G | 1 | | 0 |
| **chr19** | 33349688 | 33349688 | SNV | T | G:T | 1 | | 0 |
| **chr19** | 33355517 | 33355517 | SNV | G | G:T | 1 | | 0 |
| **chr19** | 33321952 | 33321952 | SNV | C | C:G | 1 | | 0 |
| **chr19** | 33348673 | 33348673 | SNV | T | C:T | 4 | | 0 |
| **chr19** | 33336858 | 33336858 | SNV | C | C:T | 1 | | 0 |
| **chr19** | 33333725 | 33333725 | SNV | G | G:T | 1 | | 0 |
| **chr19** | 33323595 | 33323595 | SNV | G | A:G | 1 | | 0 |
| **chr19** | 33354834 | 33354834 | SNV | G | A:G | 1 | | 0 |
| **chr19** | 33332843 | 33332843 | SNV | G | A:G | 1 | | 0 |
| **chr19** | 33334483 | 33334483 | SNV | T | C:T | 1 | | 0 |
| **chr19** | 33334224 | 33334224 | SNV | G | A:G | 2 | | 0 |
| **chr19** | 33325069 | 33325069 | SNV | T | C:T | 1 | | 0 |
| **chr19** | 33329350 | 33329350 | SNV | T | C:T | 1 | | 0 |
| **chr19** | 33337365 | 33337365 | SNV | G | A:G | 5 | | 0 |
| **chr19** | 33331061 | 33331061 | SNV | C | C:T | 2 | | 0 |
| **chr19** | 33327791 | 33327800 | deletion | TCTGATAAGA | -:TCTGATAAGA | 3 | | 0 |
| **chr19** | 33351228 | 33351228 | SNV | C | C:T | 1 | | 0 |
| **chr19** | 33338557 | 33338557 | SNV | G | G:T | 1 | | 0 |
| **chr19** | 33327286 | 33327286 | SNV | T | C:T | 1 | | 0 |
| **chr19** | 33334883 | 33334883 | SNV | G | A:G | 5 | | 0 |
| **chr19** | 33354004 | 33354004 | SNV | T | C:T | 1 | | 0 |
| **chr19** | 33337244 | 33337244 | SNV | C | C:T | 1 | | 0 |
| **chr19** | 33357093 | 33357093 | SNV | C | C:T | 1 | | 0 |
| **chr19** | 33326701 | 33326701 | SNV | T | G:T | 1 | | 0 |
| **chr19** | 33354110 | 33354110 | SNV | C | C:T | 2 | | 0 |
| **chr19** | 33326421 | 33326422 | deletion | TG | -:TG | 4 | | 0 |
| **chr19** | 33345639 | 33345639 | SNV | A | A:G | 1 | | 0 |
| **chr19** | 33335969 | 33335969 | SNV | C | C:G | 2 | | 0 |
| **chr19** | 33330330 | 33330330 | SNV | G | A:G | 2 | | 0 |
| **chr19** | 33334007 | 33334007 | SNV | C | C:T | 2 | | 0 |
| **chr19** | 33326128 | 33326128 | SNV | T | A:T | 1 | | 0 |
| **chr19** | 33350587 | 33350587 | SNV | G | A:G | 1 | | 0 |
| **chr19** | 33350341 | 33350341 | SNV | C | A:C | 1 | | 0 |
| **chr19** | 33326361 | 33326361 | SNV | G | G:T | 1 | | 0 |
| **chr19** | 33351671 | 33351671 | SNV | C | A:C | 1 | | 0 |
| **chr19** | 33359408 | 33359408 | SNV | C\|E | C:T\|E:E | 1 | | 0 |
| **chr19** | 33342858 | 33342858 | SNV | A | A:G | 3 | | 0 |
| **chr19** | 33342733 | 33342733 | SNV | A | A:T | 1 | | 0 |
| **chr19** | 33346387 | 33346387 | SNV | C | C:T | 1 | | 0 |
| **chr19** | 33335846 | 33335846 | SNV | G | A:G | 3 | | 0 |
| **chr19** | 33357957 | 33357957 | SNV | G | A:G | 1 | | 0 |
| **chr19** | 33345496 | 33345496 | SNV | G | G:T | 3 | | 0 |
| **chr19** | 33330403 | 33330403 | SNV | G | C:G | 1 | | 0 |
| **chr19** | 33358647 | 33358647 | SNV | T | C:T | 1 | | 0 |
| **chr19** | 33358250 | 33358250 | SNV | G | G:T | 1 | | 0 |
| **chr19** | 33324055 | 33324055 | SNV | T\|K | C:T\|E:K | 1 | | 0 |
| **chr19** | 33328562 | 33328562 | SNV | G | G:T | 2 | | 0 |
| **chr19** | 33349352 | 33349352 | SNV | G\|A | A:G\|V:A | 1 | | 0 |
| **chr19** | 33327663 | 33327663 | SNV | C | C:T | 1 | | 0 |
| **chr19** | 33350633 | 33350633 | SNV | G | A:G | 1 | | 0 |
| **chr19** | 33339671 | 33339671 | SNV | A | A:G | 1 | | 0 |
| **chr19** | 33345271 | 33345271 | SNV | G | A:G | 1 | | 0 |
| **chr19** | 33354944 | 33354944 | SNV | C | C:T | 1 | | 0 |
| **chr19** | 33334393 | 33334393 | SNV | G | C:G | 1 | | 0 |
| **chr19** | 33331072 | 33331072 | SNV | T | C:T | 1 | | 0 |
| **chr19** | 33330213 | 33330213 | SNV | C | C:G | 4 | | 0 |
| **chr19** | 33335711 | 33335711 | SNV | C | C:T | 1 | | 0 |
| **chr19** | 33351889 | 33351889 | SNV | G | A:G | 2 | | 0 |
| **chr19** | 33359275 | 33359275 | SNV | G | A:G | 1 | | 0 |
| **chr19** | 33343392 | 33343392 | SNV | T | C:T | 4 | | 0 |
| **chr19** | 33349141 | 33349141 | SNV | C | C:T | 2 | | 0 |
| **chr19** | 33352525 | 33352525 | SNV | T | C:T | 1 | | 0 |
| **chr19** | 33354109 | 33354109 | SNV | C | C:T | 1 | | 0 |
| **chr19** | 33355341 | 33355341 | SNV | G | A:G | 2 | | 0 |
| **chr19** | 33355151 | 33355151 | SNV | T\|Y | C:T\|C:Y | 1 | | 0 |
| **chr19** | 33330638 | 33330638 | SNV | G | A:G | 1 | | 0 |
| **chr19** | 33353785 | 33353785 | SNV | C | C:T | 1 | | 0 |
| **chr19** | 33353324 | 33353324 | SNV | G | A:G | 2 | | 0 |
| **chr19** | 33324344 | 33324344 | SNV | A | A:G | 1 | | 0 |
| **chr19** | 33324664 | 33324664 | SNV | T | C:T | 1 | | 0 |
| **chr19** | 33359740 | 33359740 | SNV | T | C:T | 1 | | 0 |
| **chr19** | 33325949 | 33325949 | SNV | G | G:T | 1 | | 0 |
| **chr19** | 33323106 | 33323106 | SNV | A | A:T | 1 | | 0 |
| **chr19** | 33338441 | 33338441 | SNV | G | A:G | 1 | | 0 |
| **chr19** | 33358711 | 33358711 | SNV | C | A:C | 1 | | 0 |
| **chr19** | 33331480 | 33331480 | SNV | C | C:T | 1 | | 0 |
| **chr19** | 33339167 | 33339167 | SNV | G | C:G | 1 | | 0 |
| **chr19** | 33324684 | 33324684 | SNV | C | C:T | 1 | | 0 |
| **chr19** | 33323557 | 33323557 | SNV | C | C:T | 2 | | 0 |
| **chr19** | 33358914 | 33358914 | SNV | A | A:G | 2 | | 0 |
| **chr19** | 33343553 | 33343553 | SNV | T | C:T | 1 | | 0 |
| **chr19** | 33359306 | 33359306 | SNV | G | A:G | 1 | | 0 |
| **chr19** | 33349318 | 33349318 | SNV | C | A:C | 1 | | 0 |
| **chr19** | 33346428 | 33346428 | SNV | A | A:G | 1 | | 0 |
| **chr19** | 33337195 | 33337195 | SNV | G | A:G | 1 | | 0 |
| **chr19** | 33345921 | 33345921 | SNV | G | A:G | 1 | | 0 |
| **chr19** | 33334180 | 33334180 | SNV | T | C:T | 1 | | 0 |
| **chr19** | 33357316 | 33357316 | SNV | G | A:G | 1 | | 0 |
| **chr19** | 33357981 | 33357981 | SNV | A | A:C | 4 | | 0 |
| **chr19** | 33357845 | 33357845 | SNV | C | C:T | 3 | | 0 |
| **chr19** | 33353001 | 33353001 | SNV | C | C:T | 1 | | 0 |
| **chr19** | 33331664 | 33331664 | SNV | C | C:G | 1 | | 0 |
| **chr19** | 33348917 | 33348917 | SNV | C | A:C | 1 | | 0 |
| **chr19** | 33355540 | 33355540 | SNV | G\|T | A:G\|M:T | 1 | | 0 |
| **chr19** | 33323728 | 33323728 | SNV | T | C:T | 1 | | 0 |
| **chr19** | 33346469 | 33346469 | SNV | G | A:G | 1 | | 0 |
| **chr19** | 33324211 | 33324211 | SNV | C\|V | C:T\|V:I | 1 | | 0 |
| **chr19** | 33347706 | 33347706 | SNV | C | C:T | 1 | | 0 |
| **chr19** | 33338418 | 33338418 | SNV | G | A:G | 1 | | 0 |
| **chr19** | 33321454 | 33321454 | SNV | A | A:G | 1 | | 0 |
| **chr19** | 33356201 | 33356201 | SNV | A | A:G | 1 | | 0 |
| **chr19** | 33330615 | 33330615 | SNV | C | A:C | 1 | | 0 |
| **chr19** | 33356492 | 33356492 | SNV | A | A:G | 1 | | 0 |
| **chr19** | 33325653 | 33325653 | SNV | T | C:T | 1 | | 0 |
| **chr19** | 33339504 | 33339504 | SNV | C | C:G | 1 | | 0 |
| **chr19** | 33355075 | 33355075 | SNV | A\|Y | A:G\|Y:Y | 1 | | 0 |
| **chr19** | 33350636 | 33350636 | SNV | C | C:T | 1 | | 0 |
| **chr19** | 33355125 | 33355125 | SNV | C\|V | C:T\|V:I | 1 | | 0 |
| **chr19** | 33348005 | 33348005 | SNV | G | A:G | 1 | | 0 |
| **chr19** | 33345558 | 33345558 | SNV | G | A:G | 1 | | 0 |

**Supplementary Table 2. Variants of SLC7A9 in the 1KG database**

| Chr | Start Position | End Position | Variant type | Ref Genotype \| Ref Proteotype | Variant Genotype 1\| Variant Proteotype 1 | Count | Variant Genotype 2\| Variant Proteotype 2 | Count |
|---|---|---|---|---|---|---|---|---|
| **chr19** | 33351045 | 33351045 | SNV | A | A:G | 1 | | 0 |
| **chr19** | 33323171 | 33323171 | SNV | G | A:G | 1 | | 0 |
| **chr19** | 33323924 | 33323926 | deletion | TCC | -:TCC | 3 | | 0 |
| **chr19** | 33356274 | 33356274 | SNV | A | A:C | 2 | | 0 |
| **chr19** | 33323011 | 33323011 | SNV | G | A:G | 1 | | 0 |
| **chr19** | 33346049 | 33346049 | SNV | T | C:T | 2 | | 0 |
| **chr19** | 33358641 | 33358641 | SNV | A | A:G | 2 | | 0 |
| **chr19** | 33351487 | 33351487 | SNV | T | A:T | 1 | | 0 |
| **chr19** | 33336685 | 33336685 | SNV | G | A:G | 1 | | 0 |
| **chr19** | 33356752 | 33356752 | SNV | C | C:T | 1 | | 0 |
| **chr19** | 33327179 | 33327179 | SNV | T | C:T | 1 | | 0 |
| **chr19** | 33336166 | 33336166 | SNV | C | C:T | 1 | | 0 |
| **chr19** | 33326268 | 33326268 | SNV | A | A:C | 1 | | 0 |
| **chr19** | 33323383 | 33323383 | SNV | C | C:T | 2 | | 0 |
| **chr19** | 33335055 | 33335055 | SNV | C | C:T | 1 | | 0 |
| **chr19** | 33324332 | 33324332 | SNV | C | C:G | 2 | | 0 |
| **chr19** | 33334502 | 33334502 | SNV | A | A:G | 1 | | 0 |
| **chr19** | 33359942 | 33359942 | SNV | C | C:T | 1 | | 0 |
| **chr19** | 33355587 | 33355587 | SNV | A\|A | A:T\|A:A | 1 | | 0 |
| **chr19** | 33343346 | 33343346 | SNV | G | A:G | 1 | | 0 |
| **chr19** | 33356313 | 33356313 | SNV | A | A:G | 1 | | 0 |
| **chr19** | 33358721 | 33358721 | SNV | C | C:T | 1 | | 0 |
| **chr19** | 33351196 | 33351196 | SNV | C | C:T | 1 | | 0 |
| **chr19** | 33352270 | 33352270 | SNV | A | A:C | 1 | | 0 |
| **chr19** | 33343914 | 33343914 | SNV | G | C:G | 6 | | 0 |
| **chr19** | 33325757 | 33325757 | SNV | C | A:C | 1 | | 0 |
| **chr19** | 33345908 | 33345908 | SNV | T | A:T | 2 | | 0 |
| **chr19** | 33333679 | 33333679 | SNV | C | C:T | 1 | | 0 |
| **chr19** | 33338226 | 33338226 | SNV | C | C:T | 4 | | 0 |
| **chr19** | 33335652 | 33335652 | SNV | G | A:G | 1 | | 0 |
| **chr19** | 33359873 | 33359873 | SNV | C | C:T | 1 | | 0 |
| **chr19** | 33333359 | 33333359 | SNV | T | G:T | 1 | | 0 |
| **chr19** | 33332042 | 33332042 | SNV | G | C:G | 1 | | 0 |
| **chr19** | 33321680 | 33321680 | SNV | T | G:T | 1 | | 0 |
| **chr19** | 33353409 | 33353409 | SNV | C\|V | C:T\|V:M | 1 | | 0 |
| **chr19** | 33359588 | 33359588 | SNV | G | A:G | 1 | | 0 |
| **chr19** | 33336437 | 33336437 | SNV | G | C:G | 1 | | 0 |
| **chr19** | 33334652 | 33334652 | SNV | A | A:G | 1 | | 0 |
| **chr19** | 33323219 | 33323219 | SNV | G | A:G | 2 | | 0 |
| **chr19** | 33322249 | 33322249 | SNV | G | A:G | 1 | | 0 |
| **chr19** | 33355842 | 33355842 | SNV | G | A:G | 1 | | 0 |
| **chr19** | 33353953 | 33353953 | SNV | C | C:T | 1 | | 0 |
| **chr19** | 33322884 | 33322884 | SNV | A | A:G | 1 | | 0 |
| **chr19** | 33324116 | 33324116 | SNV | G\|S | A:G\|S:S | 1 | | 0 |
| **chr19** | 33346420 | 33346420 | SNV | A | A:G | 1 | | 0 |
| **chr19** | 33322557 | 33322557 | SNV | C | C:T | 1 | | 0 |
| **chr19** | 33343601 | 33343601 | SNV | A | A:G | 1 | | 0 |
| **chr19** | 33337958 | 33337958 | SNV | A | A:G | 1 | | 0 |
| **chr19** | 33326392 | 33326392 | SNV | T | C:T | 1 | | 0 |
| **chr19** | 33354148 | 33354148 | SNV | G | G:T | 1 | | 0 |
| **chr19** | 33328256 | 33328256 | SNV | C | C:T | 1 | | 0 |
| **chr19** | 33330849 | 33330849 | SNV | T | C:T | 1 | | 0 |
| **chr19** | 33335191 | 33335191 | SNV | T | C:T | 1 | | 0 |
| **chr19** | 33324586 | 33324586 | SNV | C | C:T | 1 | | 0 |
| **chr19** | 33324619 | 33324619 | SNV | C | C:T | 1 | | 0 |
| **chr19** | 33346376 | 33346376 | SNV | C | C:T | 1 | | 0 |
| **chr19** | 33322674 | 33322674 | SNV | T | G:T | 1 | | 0 |
| **chr19** | 33347319 | 33347319 | SNV | C | C:T | 8 | | 0 |
| **chr19** | 33350171 | 33350171 | SNV | G | A:G | 1 | | 0 |
| **chr19** | 33349127 | 33349127 | SNV | C | C:T | 1 | | 0 |
| **chr19** | 33325468 | 33325468 | SNV | G | A:G | 1 | | 0 |
| **chr19** | 33338567 | 33338567 | SNV | A | A:G | 3 | | 0 |
| **chr19** | 33355539 | 33355539 | SNV | C\|T | C:T\|T:T | 1 | | 0 |
| **chr19** | 33338144 | 33338144 | SNV | G | G:T | 1 | | 0 |
| **chr19** | 33355389 | 33355389 | SNV | G | A:G | 1 | | 0 |
| **chr19** | 33332503 | 33332503 | SNV | A | A:G | 4 | | 0 |
| **chr19** | 33351270 | 33351270 | SNV | C | C:T | 4 | | 0 |
| **chr19** | 33348253 | 33348253 | SNV | G | A:G | 1 | | 0 |
| **chr19** | 33360620 | 33360620 | SNV | G | A:G | 1 | | 0 |
| **chr19** | 33346401 | 33346401 | SNV | C | C:T | 1 | | 0 |
| **chr19** | 33353113 | 33353113 | SNV | C\|K | A:C\|N:K | 1 | | 0 |
| **chr19** | 33360433 | 33360433 | SNV | G | A:G | 5 | | 0 |
| **chr19** | 33323660 | 33323660 | SNV | G | G:T | 1 | | 0 |
| **chr19** | 33335329 | 33335329 | SNV | G | A:G | 1 | | 0 |
| **chr19** | 33333102 | 33333102 | SNV | G\|T | C:G\|R:T | 1 | | 0 |
| **chr19** | 33335561 | 33335561 | SNV | C | A:C | 1 | | 0 |
| **chr19** | 33345552 | 33345552 | SNV | T | C:T | 1 | | 0 |
| **chr19** | 33357190 | 33357190 | SNV | C | C:G | 1 | | 0 |
| **chr19** | 33339319 | 33339319 | SNV | A | A:G | 1 | | 0 |
| **chr19** | 33322115 | 33322115 | SNV | A | A:G | 3 | | 0 |
| **chr19** | 33351475 | 33351475 | SNV | G | G:T | 1 | | 0 |
| **chr19** | 33348144 | 33348144 | SNV | G | C:G | 1 | | 0 |
| **chr19** | 33348621 | 33348621 | SNV | C | C:T | 1 | | 0 |
| **chr19** | 33325735 | 33325735 | SNV | C | C:G | 1 | | 0 |
| **chr19** | 33344467 | 33344467 | SNV | A | A:G | 1 | | 0 |
| **chr19** | 33360109 | 33360109 | SNV | A | A:G | 2 | | 0 |
| **chr19** | 33344947 | 33344947 | SNV | T | C:T | 3 | | 0 |
| **chr19** | 33339801 | 33339801 | SNV | C | C:T | 5 | | 0 |
| **chr19** | 33335678 | 33335678 | SNV | G | A:G | 2 | | 0 |
| **chr19** | 33326690 | 33326690 | SNV | T | G:T | 1 | | 0 |
| **chr19** | 33346501 | 33346501 | SNV | A | A:G | 1 | | 0 |
| **chr19** | 33327516 | 33327516 | SNV | T | A:T | 1 | | 0 |
| **chr19** | 33349560 | 33349560 | SNV | G | A:G | 1 | | 0 |
| **chr19** | 33357387 | 33357387 | SNV | A | A:G | 1 | | 0 |
| **chr19** | 33321517 | 33321517 | SNV | C | C:T | 1 | | 0 |
| **chr19** | 33332349 | 33332349 | SNV | G | A:G | 1 | | 0 |
| **chr19** | 33354497 | 33354497 | SNV | C | C:G | 1 | | 0 |
| **chr19** | 33351681 | 33351696 | deletion | TTCCAGGTGAATTTTT | -:TTCCAGGTGAATTTTT | 5 | | 0 |
| **chr19** | 33349489 | 33349489 | SNV | G | C:G | 1 | | 0 |
| **chr19** | 33325960 | 33325960 | SNV | C | C:T | 1 | | 0 |
| **chr19** | 33349473 | 33349473 | SNV | C | C:T | 2 | | 0 |
| **chr19** | 33328261 | 33328261 | SNV | T | C:T | 1 | | 0 |
| **chr19** | 33332610 | 33332610 | SNV | A | A:C | 1 | | 0 |
| **chr19** | 33322285 | 33322285 | SNV | C | C:T | 1 | | 0 |
| **chr19** | 33350522 | 33350522 | SNV | C | C:T | 1 | | 0 |
| **chr19** | 33354391 | 33354391 | SNV | A | A:G | 1 | | 0 |
| **chr19** | 33355623 | 33355623 | SNV | G\|F | A:G\|F:F | 2 | | 0 |
| **chr19** | 33357391 | 33357391 | SNV | C | C:T | 1 | | 0 |
| **chr19** | 33332578 | 33332578 | SNV | C | C:G | 1 | | 0 |
| **chr19** | 33322795 | 33322795 | SNV | C | C:T | 1 | | 0 |
| **chr19** | 33355067 | 33355067 | SNV | G\|A | A:G\|V:A | 1 | | 0 |
| **chr19** | 33350015 | 33350015 | SNV | T | C:T | 3 | | 0 |
| **chr19** | 33357291 | 33357291 | SNV | A | A:G | 1 | | 0 |
| **chr19** | 33357861 | 33357861 | SNV | C | C:G | 1 | | 0 |
| **chr19** | 33323122 | 33323122 | SNV | G | G:T | 1 | | 0 |
| **chr19** | 33358117 | 33358117 | SNV | G | A:G | 1 | | 0 |
| **chr19** | 33331687 | 33331687 | SNV | G | G:T | 2 | | 0 |
| **chr19** | 33323117 | 33323117 | SNV | C | C:T | 1 | | 0 |
| **chr19** | 33325885 | 33325885 | SNV | A | A:G | 1 | | 0 |
| **chr19** | 33346432 | 33346432 | SNV | C | C:T | 1 | | 0 |
| **chr19** | 33348079 | 33348079 | SNV | G | A:G | 1 | | 0 |
| **chr19** | 33331088 | 33331088 | SNV | T | C:T | 1 | | 0 |
| **chr19** | 33345942 | 33345942 | SNV | A | A:C | 1 | | 0 |
| **chr19** | 33324224 | 33324224 | SNV | G\|P | A:G\|P:P | 1 | | 0 |
| **chr19** | 33346629 | 33346629 | SNV | C | C:T | 2 | | 0 |
| **chr19** | 33360654 | 33360654 | SNV | G | A:G | 1 | | 0 |
| **chr19** | 33340109 | 33340109 | SNV | A | A:G | 1 | | 0 |
| **chr19** | 33346888 | 33346888 | SNV | A | A:G | 1 | | 0 |
| **chr19** | 33343417 | 33343417 | SNV | C | C:T | 1 | | 0 |
| **chr19** | 33350586 | 33350586 | SNV | C | A:C | 1 | | 0 |
| **chr19** | 33354898 | 33354898 | SNV | T | C:T | 1 | | 0 |

**Supplementary Table 2. Variants of SLC7A9 in the 1KG database**

| Chr | Start Position | End Position | Variant type | Ref Genotype \| Ref Proteotype | Variant Genotype 1\| Variant Proteotype 1 | Count | Variant Genotype 2\| Variant Proteotype 2 | Count |
|---|---|---|---|---|---|---|---|---|
| **chr19** | 33321419 | 33321419 | SNV | T | C:T | 1 | | 0 |
| **chr19** | 33350314 | 33350314 | SNV | C | C:T | 1 | | 0 |
| **chr19** | 33322495 | 33322495 | SNV | G | A:G | 1 | | 0 |
| **chr19** | 33356594 | 33356594 | SNV | C | C:T | 1 | | 0 |
| **chr19** | 33339351 | 33339351 | SNV | C | A:C | 1 | | 0 |
| **chr19** | 33352338 | 33352338 | SNV | C | C:G | 1 | | 0 |
| **chr19** | 33345787 | 33345787 | SNV | C | C:T | 1 | | 0 |
| **chr19** | 33349839 | 33349839 | SNV | C | C:T | 1 | | 0 |
| **chr19** | 33346925 | 33346925 | SNV | G | C:G | 1 | | 0 |
| **chr19** | 33357442 | 33357442 | SNV | C | C:T | 1 | | 0 |
| **chr19** | 33330648 | 33330648 | SNV | T | C:T | 1 | | 0 |
| **chr19** | 33346018 | 33346018 | SNV | G | A:G | 1 | | 0 |
| **chr19** | 33323575 | 33323575 | SNV | T | C:T | 1 | | 0 |
| **chr19** | 33334023 | 33334023 | SNV | T | G:T | 1 | | 0 |
| **chr19** | 33333643 | 33333643 | SNV | T | G:T | 1 | | 0 |
| **chr19** | 33338705 | 33338705 | SNV | A | A:G | 5 | | 0 |
| **chr19** | 33354425 | 33354425 | SNV | C | A:C | 2 | | 0 |
| **chr19** | 33348686 | 33348686 | SNV | A | A:C | 1 | | 0 |
| **chr19** | 33349034 | 33349034 | SNV | C | C:T | 2 | | 0 |
| **chr19** | 33336600 | 33336600 | SNV | C | C:T | 1 | | 0 |
| **chr19** | 33333137 | 33333137 | SNV | G\|G | A:G\|G:G | 2 | | 0 |
| **chr19** | 33336149 | 33336149 | SNV | C | C:T | 1 | | 0 |
| **chr19** | 33335203 | 33335203 | SNV | A | A:T | 1 | | 0 |
| **chr19** | 33351912 | 33351912 | SNV | C | C:T | 2 | | 0 |
| **chr19** | 33351827 | 33351827 | SNV | C | A:C | 1 | | 0 |
| **chr19** | 33323621 | 33323621 | SNV | C | C:T | 3 | | 0 |
| **chr19** | 33353328 | 33353328 | SNV | G | A:G | 3 | | 0 |
| **chr19** | 33335520 | 33335520 | SNV | G | A:G | 1 | | 0 |
| **chr19** | 33332027 | 33332027 | SNV | G | G:T | 1 | | 0 |
| **chr19** | 33327770 | 33327770 | SNV | T | C:T | 1 | | 0 |
| **chr19** | 33333613 | 33333613 | SNV | A | A:G | 2 | | 0 |
| **chr19** | 33332608 | 33332608 | SNV | T | A:T | 1 | | 0 |
| **chr19** | 33328269 | 33328269 | SNV | C | C:T | 2 | | 0 |
| **chr19** | 33334374 | 33334374 | SNV | A | A:C | 1 | | 0 |
| **chr19** | 33355393 | 33355393 | SNV | A | A:G | 1 | | 0 |
| **chr19** | 33355156 | 33355156 | SNV | G\|P | A:G\|P:P | 1 | | 0 |
| **chr19** | 33357940 | 33357940 | SNV | C | C:G | 1 | | 0 |
| **chr19** | 33359516 | 33359516 | SNV | C | C:T | 1 | | 0 |
| **chr19** | 33350420 | 33350420 | SNV | C | C:T | 1 | | 0 |
| **chr19** | 33331520 | 33331520 | SNV | A | A:G | 2 | | 0 |
| **chr19** | 33350316 | 33350316 | SNV | T | C:T | 1 | | 0 |
| **chr19** | 33355622 | 33355622 | SNV | C\|V | C:T\|V:I | 1 | | 0 |
| **chr19** | 33329152 | 33329152 | SNV | T | G:T | 1 | | 0 |
| **chr19** | 33339235 | 33339235 | SNV | G | A:G | 1 | | 0 |
| **chr19** | 33327214 | 33327214 | SNV | C | A:C | 1 | | 0 |
| **chr19** | 33351579 | 33351579 | SNV | C | A:C | 1 | | 0 |
| **chr19** | 33336308 | 33336308 | SNV | G | G:T | 1 | | 0 |
| **chr19** | 33334137 | 33334137 | SNV | C | C:G | 1 | | 0 |
| **chr19** | 33325575 | 33325575 | SNV | A | A:T | 1 | | 0 |
| **chr19** | 33358198 | 33358198 | SNV | G | A:G | 1 | | 0 |
| **chr19** | 33339038 | 33339038 | SNV | C | C:T | 2 | | 0 |
| **chr19** | 33358292 | 33358292 | SNV | G | A:G | 2 | | 0 |
| **chr19** | 33324076 | 33324076 | SNV | C\|G | C:T\|G:R | 1 | | 0 |
| **chr19** | 33323545 | 33323545 | SNV | C | C:T | 2 | | 0 |
| **chr19** | 33326730 | 33326730 | SNV | A | A:G | 1 | | 0 |
| **chr19** | 33334872 | 33334872 | SNV | C | C:G | 1 | | 0 |
| **chr19** | 33346258 | 33346258 | SNV | T | C:T | 1 | | 0 |
| **chr19** | 33322449 | 33322449 | SNV | C | C:T | 2 | | 0 |
| **chr19** | 33345049 | 33345049 | SNV | G | G:T | 1 | | 0 |
| **chr19** | 33323985 | 33323985 | SNV | C | C:G | 1 | | 0 |
| **chr19** | 33354018 | 33354018 | SNV | T | G:T | 1 | | 0 |
| **chr19** | 33357355 | 33357355 | SNV | G | G:T | 1 | | 0 |
| **chr19** | 33324001 | 33324001 | SNV | G | A:G | 1 | | 0 |
| **chr19** | 33347175 | 33347175 | SNV | A | A:G | 2 | | 0 |
| **chr19** | 33323436 | 33323436 | SNV | G | C:G | 2 | | 0 |
| **chr19** | 33334619 | 33334619 | SNV | C | C:G | 1 | | 0 |
| **chr19** | 33332965 | 33332965 | SNV | G | A:G | 1 | | 0 |
| **chr19** | 33355343 | 33355343 | SNV | G | A:G | 1 | | 0 |
| **chr19** | 33355008 | 33355008 | SNV | C\|A | C:T\|A:T | 3 | | 0 |
| **chr19** | 33339729 | 33339729 | SNV | T | C:T | 1 | | 0 |
| **chr19** | 33333084 | 33333084 | SNV | C\|R | C:G\|R:T | 1 | | 0 |
| **chr19** | 33346397 | 33346397 | SNV | A | A:G | 1 | | 0 |
| **chr19** | 33333247 | 33333247 | SNV | G | A:G | 1 | | 0 |
| **chr19** | 33322320 | 33322320 | SNV | G | A:G | 1 | | 0 |
| **chr19** | 33337110 | 33337110 | SNV | C | C:T | 3 | | 0 |
| **chr19** | 33321869 | 33321869 | SNV | A | A:C | 1 | | 0 |
| **chr19** | 33322344 | 33322344 | SNV | C | C:T | 1 | | 0 |
| **chr19** | 33333565 | 33333565 | SNV | C | C:T | 1 | | 0 |
| **chr19** | 33347778 | 33347778 | SNV | G | G:T | 3 | | 0 |
| **chr19** | 33349142 | 33349142 | SNV | G | A:G | 2 | | 0 |
| **chr19** | 33334913 | 33334913 | SNV | G | A:G | 1 | | 0 |
| **chr19** | 33330262 | 33330262 | SNV | C | C:T | 1 | | 0 |
| **chr19** | 33356971 | 33356971 | SNV | A | A:G | 1 | | 0 |
| **chr19** | 33351825 | 33351825 | SNV | G | A:G | 1 | | 0 |
| **chr19** | 33359415 | 33359415 | SNV | C\|R | C:T\|R:Q | 1 | | 0 |
| **chr19** | 33322430 | 33322430 | SNV | C | C:T | 3 | | 0 |
| **chr19** | 33335264 | 33335264 | SNV | A | A:G | 1 | | 0 |
| **chr19** | 33325736 | 33325736 | SNV | G | A:G | 1 | | 0 |
| **chr19** | 33349431 | 33349431 | SNV | G\|L | C:G\|V:L | 1 | | 0 |
| **chr19** | 33335478 | 33335478 | SNV | C | C:G | 2 | | 0 |
| **chr19** | 33322562 | 33322562 | SNV | C | C:G | 1 | | 0 |
| **chr19** | 33354235 | 33354235 | SNV | G | A:G | 1 | | 0 |
| **chr19** | 33356208 | 33356208 | SNV | T | G:T | 1 | | 0 |
| **chr19** | 33327502 | 33327502 | SNV | G | G:T | 3 | | 0 |
